# Heart Stress, Frailty and Mortality Risk in two prospective cohorts

**DOI:** 10.64898/2026.01.25.26344776

**Authors:** Yaqi Huang, Meng Hao, Shuai Jiang, Xiangnan Li, Yulong Tang, Zixin Hu, Xiaofeng Wang, Li Han, Yi Li, Hui Zhang

**Author notes:** Corresponding author: Hui Zhang, School of Global Health, Chinese Center for Tropical Diseases Research, Shanghai Jiao Tong University School of Medicine, Shanghai, 200025, China.; Yi Li, Human Phenome Institute, Zhangjiang Fudan International Innovation Centre, Fudan University, Shanghai, 201203, China. These authors contributed equally (Yaqi Huang, Meng Hao).

## Abstract

**Importance:** Frailty is a multisystem syndrome that reflects age-related physiological decline, underscoring the need for more biologically informed risk stratification within frailty assessments. Frailty and heart stress (HS) are individually associated with increased mortality risk, but their combined effects remain practically unexplored.

**Objective:** To evaluate whether the combined exposure to frailty and HS is associated with an increased risk of mortality.

**Design, Setting, and Participants:** This prospective cohort study used data from the US National Health and Nutrition Examination Survey (NHANES) and the Health and Retirement Study (HRS). Participants with complete data on frailty and HS were included. Analyses was performed between May 2025 and October 2025.

**Exposure:** Frailty was assessed using three frailty indices (FI) based on self-reported items (FI-Self-report), blood biomarkers (FI-Lab), and their combination (FI-Combined). HS was defined by age-adjusted elevation in N-terminal pro-B-type natriuretic peptide (NT-proBNP) levels. Participants were estimate into four groups according to baseline frailty and HS status.

**Main Outcomes and Measures:** The primary outcome was all-cause mortality. Cox proportional hazard models were employed to calculate the hazard ratios (HRs) and 95% confidence intervals (CIs).

**Results:** A total of 12,252 participants from NHANES (mean age 49.91 years, 52.18% female), and 9,488 participants from HRS (mean age 69.16 years, 58.97% female) were included. Compared with those having neither frailty nor HS, participants with frailty and/or HS showed significantly elevated mortality risk in both cohorts, with HRs ranging from 1.81 to 5.54. The highest mortality risk was observed in participant with both frailty and HS, the HRs were 3.58 (95% CI: 3.20-4.01) for FI Self Report, 3.43 (95% CI: 3.04-3.86) for FI Lab, and 4.15 (95% CI: 3.70-4.67) for FI Combined in NHANES; the corresponding HRs were 5.02 (95% CI: 4.38-5.76), 4.73 (95% CI: 4.13-5.41), and 5.54 (95% CI: 4.84-6.35) in HRS, respectively.

**Conclusions and Relevance:** Co-occurrence of frailty and HS is common, and jointly associated with increased mortality risk in the general population. These findings support integrating HS into frailty assessments to improve mortality risk stratification and guide targeted interventions.

**Key Points:** **Question:** Is the combination of frailty and heart stress (HS) associated with increased mortality risk?

**Findings:** In this prospective cohort study including 12,252 participants from the US National Health and Nutrition Examination Survey (NHANES) and 9,488 participants from the Health and Retirement Study (HRS), participants with frailty and/or HS exhibited higher risk of all-cause mortality. The greatest mortality risk was found among participant with both frailty and HS.

**Meaning:** These findings indicate that co-occurrence of frailty and HS is associated with increased mortality risk, supporting integration of HS into frailty assessment for risk stratification and intervention.

## Introduction

Frailty is a multidimensional syndrome characterized by reduced physiologic reserve across multiple organ systems, resulting in increased susceptibility to stressors^1^. Its prevalence increases with age and affects millions of older adults worldwide^2^. Frailty is also associated with increased risks of numerous adverse outcomes, including mortality, cardiovascular disease, dementia^3–5^. Therefore, given these substantial burdens of frailty, there is an urgent need for more individualized frailty management that integrates multifaceted factors^1^. At present, some effective approaches have been advised for frail individuals, such as physical activity and nutrition^6^. However, due to the multidimensional and multisystem nature of frailty, a more integrated approach that incorporates age-related biological markers into frailty assessments is essential to improve risk stratification and inform targeted preventive strategies.

N-terminal pro-B-type natriuretic peptide (NT-proBNP), a hallmark cardiac marker of elevated ventricular wall stress and volume overload^7,8^, has been established as an independent predictor of all-cause and cardiovascular mortality^9^. Although both NT-proBNP and frailty reflect age-related physiological decline, they capture distinct risk dimensions^1,10^. NT-proBNP quantifies underlying heart disease burden, whereas frailty represents multisystem physiological dysregulation^11,12^. Notably, NT-proBNP is predominantly released from the ventricular myocardium and cleared by the kidney, thereby reflecting two key organ systems implicated in frailty development^13^, suggesting a close interaction between elevated NT-proBNP and frailty. This association was further supported by shared pathophysiological foundations, including chronic inflammation, immune system dysfunction, and neuroendocrine imbalance^14^. Recently, the concept of heart stress (HS), characterized by elevated NT-proBNP levels, has been proposed for blood pressure management in primary prevention^15,16^. Considering the intricate link between frailty and heart dysfunction, integrating HS into frailty assessment may present significant value for refining risk stratification and optimizing prevention strategies. However, whether this integration meaningfully improves risk prediction remains unclear, and there is still a lack of large-scale evidence on the joint effect of frailty and HS on mortality in the general population.

To address this knowledge gap, we first introduced the concept of HS into frailty assessments, and then examined: (1) the co-occurrence of frailty and HS, and (2) their individual and joint associations with mortality risk.

## Methods

### Study Population

The NHANES is a nationwide survey designed to assess the health and nutritional status of the U.S. population using a stratified, multistage probability sampling design^17^. For the present analysis, data from the 1999-2004 survey cycles were utilized. We excluded participants aged under 20 years and those with missing data on NT-proBNP, survival status, and FI information (**eFigure 1A**). This final analytic cohort consisted of 5,721 participants for FI-Self-report, 12,252 for FI-Lab, and 5,721 for FI-Combined. The HRS is a nationally representative longitudinal study of U.S. adults designed to examine national-level social and policy changes that may affect individuals^18^. The analysis included participants from the 2016 wave. Participants aged 50 years or older with complete data on NT-proBNP, survival status, FI information were included (**eFigure 1B**), and the final sample sizes for the three FI assessments were 9,488, 9,242, and 9,342, respectively.

**Figure 1.**
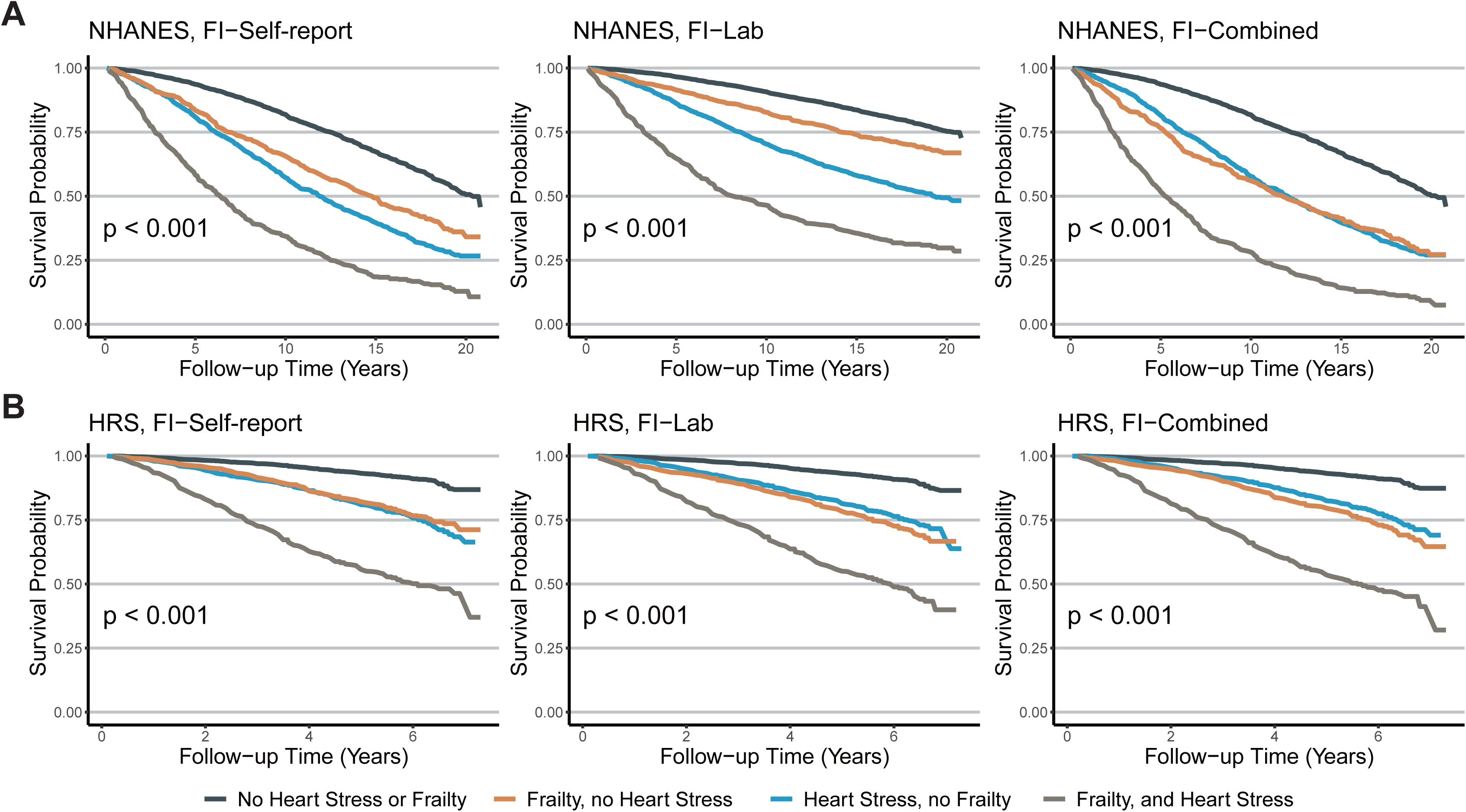
All-cause mortality stratified by frailty and HS. Kaplan-Meier plots demonstrate survival probabilities according to baseline frailty and HS status across 4 groups in the NHANES (A) and the HRS (B) cohorts. HS: heart stress. NHANES: National Health and Nutrition Examination Survey. HRS: Health and Retirement Study

### Frailty Assessment

Frailty was assessed using the frailty index (FI), following the standardized procedures established by Rockwood et.al^19–23^. For this study, we constructed three distinct FIs based on self-reported health deficits (FI-Self-report), blood biomarkers (FI-Lab), and their combinations (FI-Combined). FI scores were calculated as the ratio of the number of deficits present to the total number of deficits assessed, yielding a score from 0 to 1 (e.g., 10/40 deficits=0.25). To avoid conceptual overlap with HS, self-reported heart disease was excluded. Consequently, the FI-Self-report comprised 34 items for NHANES and 40 for HRS. Both cohorts shared 32 common items for FI-Lab, resulting in the FI-Combined of 66 items for NHANES and 72 for HRS. FI scores were calculated only for participants with less than 20% missing data across the included deficits. Finally, participants were categorized into frail and non-frail groups using a FI cutoff of 0.25. A detailed list of all FI items is provided in **eTable 1-3**.

### Heart Stress Definition

Given that NT-proBNP concentrations increase with age, HS was defined as age-adjusted NT-proBNP cutoffs to enhance specificity in older adults^16,24^. In accordance with recommendations from the Heart Failure Association of the European Society of Cardiology, the cutoffs were set at 75 pg/mL for individuals aged <50 years, 150 pg/mL for those aged 50-74 years and 300 pg/mL for those aged ≥75 years^15,24^. In NHAENS, NT-proBNP was measured in serum on the Roche Cobas e601 autoanalyzer at the University of Maryland School of Medicine, Baltimore, Maryland. The lower and upper limits of detection were 5 pg/ml and 35000 pg/ml, respectively, and the variation coefficient of NT-proBNP was between 2.7 and 3.1%^25^. In HRS, NT-proBNP was tested in serum on the Roche e411 analyzer utilizing a sandwich immunoassay method. The coefficient of variation was 3.3% at 140 pg/mL and 2.8% at 4707 pg/mL^26^.

### Outcomes

The primary outcome of this study was all-cause mortality, and the secondary outcome was cause-specific mortality, including cardiovascular disease (CVD), heart disease, cerebrovascular disease, cancer and other causes. In NHANES, mortality data were extracted from the National Death Index (NDI) database. The follow-up time extended from the date of participation until the date of death or December 31, 2019 (the last update date of the NDI database). The HRS employed various approaches to track participants over time, including gathering information from spouses and proxy respondents to determine the vital status at each wave. Participants who withdrew from the survey while still alive were retained in the analysis, and any available mortality information was incorporated.

### Covariates

The covariates comprised demographic characteristics and lifestyle factors. The demographic characteristics included age, gender, race/ethnicity, education and marital status. In NHANES, race/ethnicity was categorized as White (non-Hispanic White), Black (non-Hispanic Black), and Others (Mexican American, Other Hispanic, and Other Race). Education was categorized as less than high school, high school, and college or above. For marital status, in NHANES, participants reporting being married or living with partner were classified as married/living with partner, whereas those reporting being widowed, divorced, separated, or never married were classified as others. In the HRS, participants reporting being married, married with spouse absent, or partnered were categorized as married/living with partner, while those reporting being separated, divorced, separated/divorced, widowed, or never married were categorized as others. Lifestyle factors included body mass index (BMI), smoking status and vigorous physical activity. BMI was categorized as <25 kg/m² (normal or underweight), 25.0-29.9 kg/m² (overweight), and ≥30.0 kg/m² (obese). Smoking status (ever vs never) and vigorous physical activity (yes vs no) were captured through self-reported data of participants.

### Statistical Analysis

Participants were divided into four groups according to baseline frailty and HS status: no frailty or HS, HS and no frailty, frailty and no HS, both frailty and HS. Continuous variables were presented as mean (SD) and categorical variables as frequency (%). Cox proportional hazards regression models were used to evaluate the associations of frailty and HS with all-cause and cause-specific mortality using three models. Model 1 was the crude model; Model 2 was adjusted for age, gender, and race/ethnicity; and Model 3 was additionally adjusted for education, BMI, smoking status, physical activity, and marital status. Participants with no frailty or HS were served as the reference group. Moreover, subgroup and stratified analyses were performed based on age group, gender, BMI category, race/ethnicity, smoking status, vigorous physical activity, marital status and educational status using the fully adjusted Model 3. The P values for interactions were evaluated by including the cross-product terms for each assessed factor. Additionally, survival probabilities across the four groups were visualized using Kaplan-Meier curves, with log-rank tests applied for comparisons. Restricted cubic spline regressions with 3 knots were used to analyze the association between FI (as a continuous variable) and mortality risk, after adjusting for covariates (Model 3).

Sensitivity analyses were performed to assess the robustness of the main findings: (1) HS was re-defined by applying a fixed cutoff of 125 pg/mL for NT-proBNP, in accordance with prevailing heart failure guidelines and consensus statements^27^. (2) To minimize potential reverse causality, individuals who died within 2 years after baseline were excluded (NHANES: n=247 for FI-Self-report, 261 for FI-Lab, and 249 for FI-Combined; HRS: n=373 for FI-Self-report, 355 for FI-Lab, and 364 for FI-Combined). All analyses were performed using R statistical software, version 4.4.1. All P values were 2-sided and a value <0.05 was considered statistically significant.

## Results

### Baseline Characteristics of study population

The characteristics of the study populations are shown in **Table 1**. In NHANES, the analyses included 5,721 participants for the FI-Self-report (mean age 64.75 years, 50.95% female), 12,252 for the FI-Lab (mean age 49.91 years, 52.18% female), and 5,721 for the FI-Combined (mean age 64.73 years, 50.88% female). In HRS, corresponding sample sizes were 9,488, 9,242, and 9,342 participants, with a mean age of approximately 69 years and 59% female across all three FI definitions. Overall, 23.47%-33.49% participants had HS across both cohorts. After further stratification by frailty status, the majority of participants was classified as having no HS or frailty (54.68%-70.25%), followed by HS and no frailty. Furthermore, NHANES documented 3,109, 3,519, 3,114 deaths over a median follow-up of 15.40, 16.90, 15.30 years for the FI-Self-report, FI-Lab, and FI-Combined analyses, respectively. In HRS, 1,699, 1,637, 1,666 deaths were documented over a median follow-up of 5.9 years.

**Table 1.** Characteristics of study population at baseline.

|  | **NHANES** |  |  | **HRS** |  |  |
| --- | --- | --- | --- | --- | --- | --- |
| Sample, N | FI-Self-report<br>(N=5721) | FI-Lab<br>(N=12252) | FI-Combined<br>(N=5721) | FI-Self-report<br>(N=9488) | FI-Lab<br>(N=9242) | FI-Combined<br>(N=9342) |
| Age, years, M (SD) | 64.75 (15.12) | 49.91 (19.08) | 64.73 (15.15) | 69.16 (9.80) | 69.16 (9.77) | 69.17 (9.79) |
| Age Groups, N (%) |  |  |  |  |  |  |
| 20-49 | 916 (16.01) | 6358 (51.89) | 925 (16.17) | 0 (0.00) | 0 (0.00) | 0 (0.00) |
| 50-64 | 1496 (26.15) | 2585 (21.10) | 1491 (26.06) | 3667 (38.65) | 3563 (38.55) | 3605 (38.59) |
| 65 years or older | 3309 (57.84) | 3309 (27.01) | 3305 (57.77) | 5821 (61.35) | 5679 (61.45) | 5737 (61.41) |
| Gender, N (%) |  |  |  |  |  |  |
| Male | 2806 (49.05) | 5859 (47.82) | 2810 (49.12) | 3893 (41.03) | 3797 (41.08) | 3839 (41.09) |
| Female | 2915 (50.95) | 6393 (52.18) | 2911 (50.88) | 5595 (58.97) | 5445 (58.92) | 5503 (58.91) |
| BMI, kg/m<sup>2</sup>, M (SD) | 28.67 (5.98) | 28.36 (6.10) | 28.67 (6.00) | 28.97 (6.20) | 28.96 (6.18) | 28.97 (6.20) |
| BMI categories, N (%) |  |  |  |  |  |  |
| Normal or underweight | 1561 (27.29) | 3765 (30.73) | 1560 (27.27) | 2465 (25.98) | 2400 (25.97) | 2421 (25.92) |
| Overweight | 2039 (35.64) | 4301 (35.10) | 2033 (35.54) | 3437 (36.22) | 3361 (36.37) | 3394 (36.33) |
| Obese | 1872 (32.72) | 3847 (31.40) | 1874 (32.76) | 3480 (36.68) | 3379 (36.56) | 3424 (36.65) |
| Race/Ethnicity, N (%) |  |  |  |  |  |  |
| White/Caucasian | 3240 (56.63) | 6309 (51.49) | 3242 (56.67) | 6995 (73.72) | 6841 (74.02) | 6897 (73.83) |
| Black/African American | 954 (16.68) | 2180 (17.79) | 956 (16.71) | 1655 (17.44) | 1589 (17.19) | 1623 (17.37) |
| Others | 1527 (26.69) | 3763 (30.71) | 1523 (26.62) | 807 (8.51) | 782 (8.46) | 792 (8.48) |
| Education, N (%) |  |  |  |  |  |  |
| Less than high school | 2274 (39.75) | 3930 (32.08) | 2277 (39.80) | 1623 (17.11) | 1575 (17.04) | 1594 (17.06) |
| High school | 1376 (24.05) | 2926 (23.88) | 1377 (24.07) | 3119 (32.87) | 3052 (33.02) | 3083 (33.00) |
| College or above | 2056 (35.94) | 5373 (43.85) | 2052 (35.87) | 4743 (49.99) | 4613 (49.91) | 4662 (49.90) |
| Smoking status, N (%) |  |  |  |  |  |  |
| Ever | 3126 (54.64) | 5999 (48.96) | 3131 (54.73) | 5241 (55.24) | 5095 (55.13) | 5159 (55.22) |
| Never | 2586 (45.20) | 6234 (50.88) | 2581 (45.11) | 4199 (44.26) | 4099 (44.35) | 4135 (44.26) |
| Vigorous physical activity, N (%) |  |  |  |  |  |  |
| Yes | 908 (15.87) | 3354 (27.38) | 904 (15.80) | 4253 (44.83) | 4162 (45.03) | 4190 (44.85) |
| No | 4809 (84.06) | 8892 (72.58) | 4813 (84.13) | 5191 (54.71) | 5036 (54.49) | 5108 (54.68) |
| Marital status, N (%) |  |  |  |  |  |  |
| Married/Living with partner | 3300 (57.68) | 7414 (60.51) | 3289 (57.49) | 6040 (63.66) | 5895 (63.78) | 5947 (63.66) |
| Others | 2254 (39.40) | 4439 (36.23) | 2265 (39.59) | 3436 (36.21) | 3335 (36.09) | 3383 (36.21) |
| NT-proBNP, pg/mL, M (SD) | 366.70 (1473.70) | 199.55 (1034.08) | 366.91 (1472.74) | 357.44 (1618.45) | 351.64 (1610.73) | 353.36 (1612.49) |
| Frailty index, M (SD) | 0.17 (0.12) | 0.15 (0.08) | 0.17 (0.08) | 0.17 (0.15) | 0.18 (0.10) | 0.17 (0.10) |
| No Heart Stress or Frailty, N (%) | 3128 (54.68) | 8607 (70.25) | 3387 (59.20) | 5575 (58.76) | 5736 (62.06%) | 5690 (60.91%) |
| Heart Stress, no Frailty, N (%) | 1352 (23.63) | 2350 (19.18) | 1440 (25.17) | 1913 (20.16) | 1924 (20.82%) | 1892 (20.25%) |
| Frailty, no Heart Stress, N (%) | 685 (11.97) | 770 (6.28) | 418 (7.31) | 1086 (11.45) | 755 (8.17%) | 869 (9.30%) |
| Frailty, and Heart Stress, N (%) | 556 (9.72) | 525 (4.29) | 476 (8.32) | 914 (9.63) | 827 (8.95%) | 891 (9.54%) |

Missing in NHANES for the FI-self, FI-lab, and FI-combined groups, respectively: No. of BMI categories 249 (4.35%), 339 (2.77%), 254 (4.44%); education 15 (0.26%), 23 (0.19%), 15 (0.26%); smoking status 9 (0.16%), 19 (0.16%), 9 (0.16%); vigorous physical activity 4 (0.07%), 6 (0.05%), 4 (0.07%); marital status 167 (2.92%), 399 (3.26%), 167 (2.92%). Missing in HRS for the FI-self, FI-lab, and FI-combined groups, respectively: No. of BMI categories 106 (1.12%), 102 (1.10%), 103 (1.10%); race 31 (0.33%), 30 (0.32%), 30 (0.32%); education 3 (0.03%), 2 (0.02%), 3 (0.03%); smoking status 48 (0.51%), 48 (0.52%), 48 (0.51%); vigorous physical activity 44 (0.46%), 44 (0.48%), 44 (0.47%); marital status 12 (0.13%), 12 (0.13%), 12 (0.13%).
BMI: body mass index; M: mean; SD: standard deviation.

### Associations of Frailty and HS with All-cause Mortality Risk

Compared with the no frailty or HS group, participants with both frailty and HS demonstrated the highest risk of all-cause mortality in the two cohorts. This pattern was consistently observed across diverse models and FI constructions (**Table 2; eTable 4**). Specifically, in fully adjusted Cox models, the HRs were 3.58 (95% CI: 3.20-4.01) in NHANES and 5.02 (95% CI: 4.38-5.76) in HRS when frailty was defined by the FI-Self-report; 3.43 (95% CI: 3.04-3.86) and 4.73 (95% CI: 4.13-5.41) for the FI-Lab; and 4.15 (95% CI: 3.70-4.67) and 5.54 (95% CI: 4.84-6.35), for the FI-Combined. Furthermore, individuals with HS and no frailty, or frailty and no HS showed attenuated but still elevated associations with all-cause mortality risk. For example, the corresponding HRs were 1.88 (95% CI: 1.73-2.05), 2.06 (95% CI: 1.84-2.31) in NHANES; and 1.99 (95% CI: 1.75-2.27), 2.29 (95% CI:1.95-2.68) in HRS for FI-Self-report groups respectively. Additionally, Kaplan-Meier survival plots indicated that individuals in both frailty and HS groups had the greatest cumulative all-cause mortality among the four groups (**Figure 1**, log-rank P <0.001). In restricted cubic spline regressions, we observed approximately linear or superlinear association between FI increased risk of all-cause mortality in participants with/without HS (**Figure 2**).

**Figure 2.**
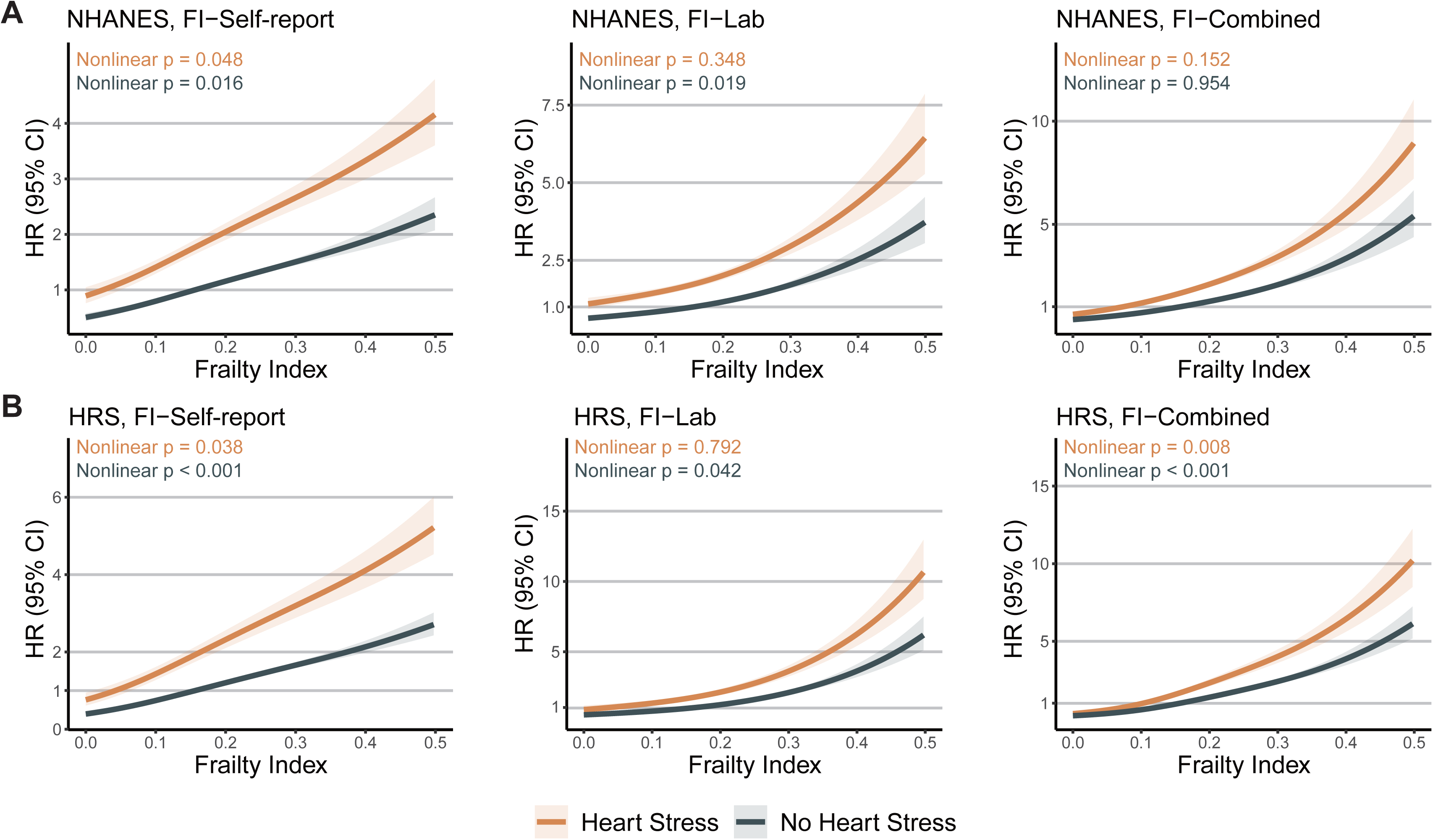
Association between baseline frailty and all-cause mortality stratified by HS status. Restricted cubic spline analyses show estimated hazard ratios for all-cause mortality across continuous levels of FI in the NHANES (A) and the HRS (B) cohorts. Models were adjusted for age, gender, race/ethnicity, education, body mass index, smoking status, physical activity, and marital status. HS: heart stress. FI: frailty index. NHANES: National Health and Nutrition Examination Survey. HRS: Health and Retirement Study

**Table 2.** Association of frailty and heart stress with risk of all-cause mortality in NHANES and HRS.

|  |  | FI-Self-report |  |  | FI-Lab |  |  | FI-Combined |  |  |
| --- | --- | --- | --- | --- | --- | --- | --- | --- | --- | --- |
|  |  | Event/N (%) | HR<br>(95%CI) | P-value | Event/N (%) | HR 95%CI | P-value | Event/N (%) | HR (95%CI) | P-value |
| **NHANES** |  |  |  |  |  |  |  |  |  |  |
| No Frailty | No HS | 1310/3128 (41.88) |  |  | 1813/8607<br>(21.06) |  |  | 1434/3387<br>(42.34) |  |  |
|  | HS | 925/1352 (68.42) | 1.88<br>(1.73-2.05) | 8.40E-47 | 1109/2350<br>(47.19) | 1.81<br>(1.67-1.96) | 1.51E-50 | 977/1440<br>(67.85) | 1.82<br>(1.67-1.98) | 6.96E-45 |
| Frailty | No HS | 404/685 (58.98) | 2.06<br>(1.84-2.31) | 3.34E-35 | 235/770<br>(30.52) | 1.91<br>(1.67-2.19) | 1.78E-20 | 279/418<br>(66.75) | 2.64<br>(2.31-3.01) | 3.79E-47 |
|  | HS | 470/556 (84.53) | 3.58<br>(3.20-4.01) | 1.61E-10<br>7 | 362/525<br>(68.95) | 3.43<br>(3.04-3.86) | 2.26E-92 | 424/476<br>(89.08) | 4.15<br>(3.70-4.67) | 1.47E-126 |
| **HRS** |  |  |  |  |  |  |  |  |  |  |
| No Frailty | No HS | 511/5575 (9.17) | Ref |  | 534/5736<br>(9.31) | Ref |  | 517/5690<br>(9.09) | Ref |  |
|  | HS | 478/1913 (24.99) | 1.99<br>(1.75-2.27) | 6.06E-26 | 465/1924<br>(24.17) | 1.86<br>(1.64-2.11) | 3.03E-21 | 441/1892<br>(23.31) | 1.85<br>(1.62-2.10) | 3.96E-20 |
| Frailty | No HS | 252/1086 (23.20) | 2.29<br>(1.95-2.68) | 6.02E-24 | 209/755<br>(27.68) | 2.44<br>(2.07-2.88) | 3.05E-26 | 238/869<br>(27.39) | 2.64<br>(2.24-3.11) | 1.76E-31 |
|  | HS | 458/914 (50.11) | 5.02<br>(4.38-5.76) | 1.21E-116 | 429/827<br>(51.87) | 4.73<br>(4.13-5.41) | 1.87E-112 | 470/891<br>(52.75) | 5.54<br>(4.84-6.35) | 6.86E-136 |

Model was fully adjusted for age, gender, race/ethnicity, education, body mass index, smoking status, physical activity, and marital status.
HS: heart stress; HR, hazard ratio; CI, confidence interval; NHANES, US National Health and Nutrition Examination Survey; HRS, Health and Retirement Study.

### Associations of Frailty and HS with Cause-specific Mortality Risk

Owing to the unavailability of cause-specific mortality data in HRS, the analysis of joint associations between frailty, HS, and cause-specific mortality risk was conducted only in the NHANES. Similar association patterns were noted for cause-specific mortality, with participants in both frailty and HS group showing the highest risks. In detail, for CVD mortality, the HRs were 5.91 (95% CI: 4.85-7.20), 4.50 (95% CI: 3.67-5.52) and 6.29 (95% CI: 5.15-7.69) for the FI-Self-report, FI-Lab and FI-Combined, respectively. For cancer mortality, the corresponding HRs were 3.64 (95% CI: 2.72-4.85), 3.71 (95% CI: 2.73-5.04) and 4.18 (95% CI:3.05-5.73) respectively. Elevated risks were also observed for mortality from other causes, with HRs of 3.40 (95% CI: 2.87–4.02), 3.81 (95% CI: 3.21–4.51), and 4.53 (95% CI: 3.82–5.38), respectively (**eTable 5**). Similar patterns were observed for heart disease and cerebrovascular disease mortality (**eTable 5**).

In addition, individuals with frailty or HS alone exhibited weaker but still significantly increased risks of cause-specific mortality. For example, under the FI-Lab definition, the corresponding HRs for CVD mortality were 2.51 (95% CI: 2.19-2.87) for HS and no frailty and 1.85 (95% CI: 1.39-2.47) for frailty and no HS, while the corresponding HRs for cancer mortality were 1.77 (95% CI: 1.49-2.11) and 2.31 (95% CI: 1.75-3.05), respectively. Furthermore, participants with both frailty and HS exhibited the highest cumulative incidence of cause-specific mortality compared to the other three groups (**eFigure 2**, log-rank P<0.001). Restricted cubic spline analyses further revealed a positive dose–response relationship between FI and cause-specific mortality (**eFigure 3**).

### Stratified Analyses in subgroups

Stratified Analyses were conducted in cohorts according to age group, gender, BMI category, race/ethnicity, smoking status, vigorous physical activity, marital status and educational status. Similarly, individuals with both frailty and HS had the highest risks of all-cause, CVD and cancer mortality in every stratum and under different assessments of frailty (**Figure 3; eTable 6-10**). Although statistically significant interactions were detected in some subgroups (P for interaction < 0.05), the stratified direction and magnitude of the associations remained generally consistent with the overall results. These findings indicated the joint effect of frailty and HS on mortality is robust independent of sociodemographic and behavioral factors. Moreover, some variation in effect size was observed across strata; for instance, more pronounced risks were observed among participants aged 50-64 years in the HRS cohort.

**Figure 3.**
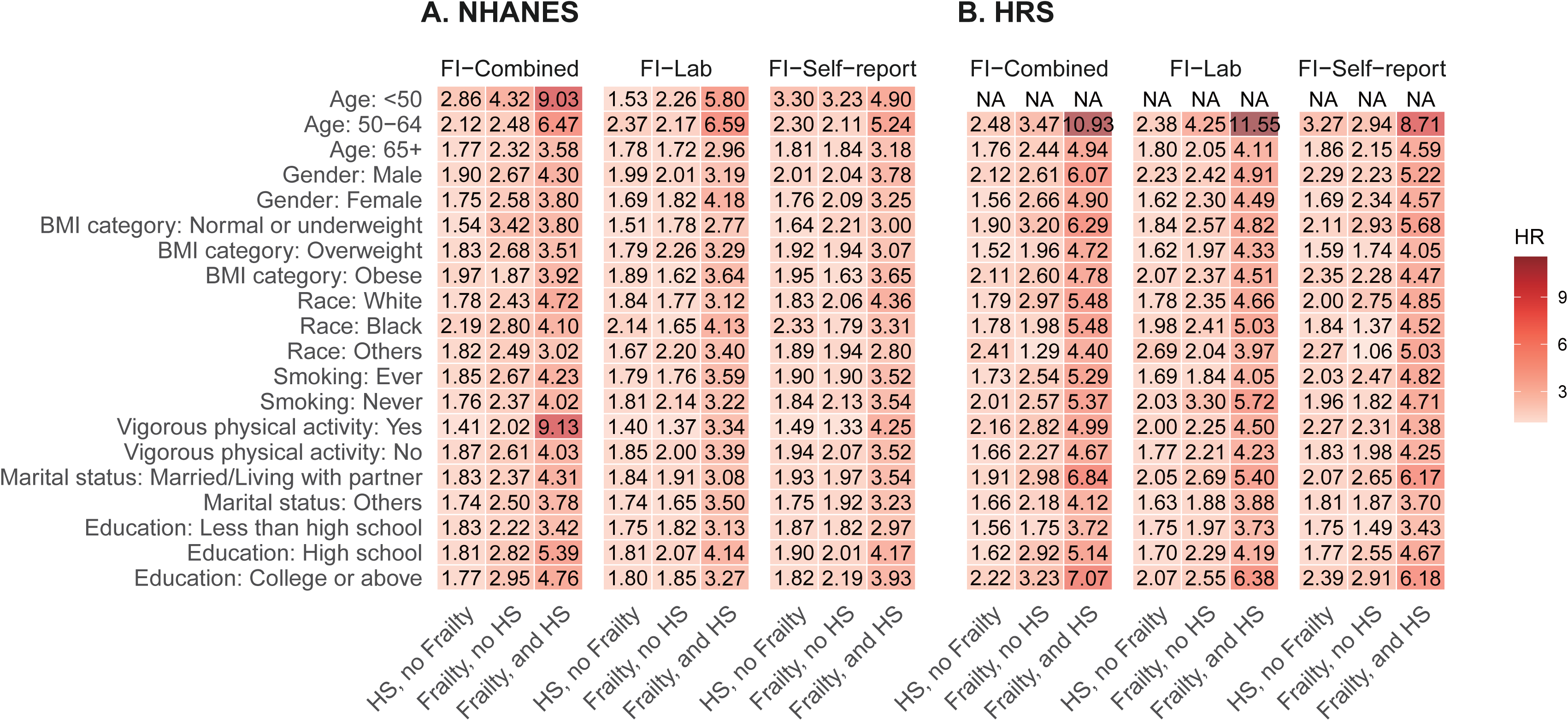
Association of frailty and HS with all-cause mortality in subgroups. Heatmaps demonstrate the HRs estimated from Cox proportional hazard models with no frailty and HS group as reference in the NHANES (A) and the HRS (B) cohorts. Models were adjusted for age, gender, race/ethnicity, education, body mass index, smoking status, physical activity, and marital status. HR indicates hazard ratio. HS: heart stress. FI: frailty index. NHANES: National Health and Nutrition Examination Survey. HRS: Health and Retirement Study

### Sensitivity Analyses

When HS was alternatively defined using a fixed NT-proBNP cutoff, the individual and joint associations of frailty and HS with all-cause and cause-specific mortality remained consistent with the main analyses (**eTable 11-17**). Consistent patterns were also observed after excluding participants who died within 2 years post-baseline (**eTable 18-19**). In line with these results, Kaplan–Meier survival curves showed the highest cumulative mortality among participants with both frailty and HS (**eFigure 4-7**). **eFigure 8-11** demonstrate a clear positive dose–response relationship between FI and mortality.

## Discussion

To our knowledge, this represents the first large-scale study to comprehensively investigate the joint association of frailty and HS with mortality risk. Using data from more than 21,000 participants across the NHANES and HRS, we observed that the coexistence of frailty and HS was associated with the highest risk of all-cause mortality compared to those with no frailty or HS. Similar results were also observed for cause-specific mortality, including CVD, cancer and other causes. This significant association remained remarkably consistent across three complementary FI constructions, and was further confirmed by stratified and sensitivity analyses. Overall, these findings underscore the coexisting frailty and HS in the general population are associated with elevated risk of mortality, and the potential utility of HS for refining risk stratification among frail adults.

Our findings extend the existing literature on synergistic risk of frailty and adverse outcomes by shifting the focus from overt disease, such as HF, to subclinical cardiac stress^28^. In contrast to prior studies that primarily examined individuals with established structural heart disease^29^, our study focuses on subclinical myocardial stress, or more broadly, the abnormal cardiac biomarkers^30^. NT-proBNP reflects myocardial wall stress and neurohormonal activation, and therefore represents cardiac overload rather than a diagnosed disease state^15^. Accordingly, compared with risk stratification and intervention after a diagnosis of HF, the joint assessment of frailty at the stage of HS may enable earlier identification of high-risk individuals and facilitate the timely initiation of comprehensive interventions, such as nutritional support^31^, with the potential to modify subsequent disease trajectories.

Importantly, our findings suggest that NT-proBNP reflects a broader state of systemic vulnerability rather than cardiac-specific pathology alone. We demonstrated that HS, as defined by elevated NT-proBNP levels, effectively identified subgroups at heightened risk of CVD, cancer, and other causes of mortality across strata defined by demographic characteristics and frailty status. While these results align with established evidence supporting the prognostic value of NT-proBNP for CVD outcomes^15,32^, and further demonstrate its capacity to enhance risk stratification for non-cardiovascular outcomes, particularly cancer. Previous studies have reported higher NT-proBNP levels among cancer survivors and suggested its prognostic relevance in oncologic outcomes^33^. Mechanistically, NT-proBNP could also be produced by cancer cells and its secretion is strongly influenced by inflammatory cytokines (e.g., interleukins), broadening its indicative capacity beyond cardiac conditions to encompass a variety of inflammatory processes^34^. Taken together, our study provides compelling evidence that NT-proBNP captures a more comprehensive dimension of biological vulnerability beyond its established role in cardiovascular risk assessment.

Using three approaches to construct FI, we observed consistently joint associations between frailty and HS, underscoring the robustness and flexibility of frailty assessment across different operational definitions. While previous studies have shown that frailty captures multidimensional vulnerability in patients across age groups^35,36^, our researches extend this evidence by applying a comprehensive FI framework that spans from subclinical to clinical deficits^20^. Specifically, the FI-Self-report captures clinically visible deficits representing the accumulation of subcellular, tissue and organ deficits, whereas the FI-Lab identifies subclinical dysregulation representing preclinical and microscopic deficits^37,38^. From a practical perspective, the FI-Self-reported screening is a cost-effective and simple tool for community-based assessment, while the FI-Lab utilizes routinely and rapidly collected data for hospital admission with minimal patient involvement^37,38^. Collectively, our study provides novel evidence that FI assessment can be flexibly adapted to diverse resource availability for individuals with cardiac stress.

Our findings align with prior mechanistic evidence suggesting pathophysiological links between frailty with HS, which partially overlap with mechanisms implicated in other cardiac conditions. On the one hand, shared biological mechanisms, including endothelial dysfunction and neurohormonal dysregulation, simultaneously contribute to the development of both conditions^39^. The interplay of common mechanisms reinforces a vicious cycle of multisystemic decline^40^. On the other hand, bidirectional pathways, such as the muscle-heart axis and metabolic-frailty-cardiovascular axis, elucidate how the clinical manifestations of one condition can directly trigger or aggravate the progression of the other^39^.

This study presents significant implications for public health and clinical management. Our findings demonstrate that HS status could help identify a high-risk subgroup of frail individuals with underlying cardiac overload. Therefore, integrating HS into routine frailty assessment could refine risk stratification, enabling the identification of the most vulnerable patients who may benefit from earlier and multifaceted interventions. From a healthcare systems perspective, this integrated approach may be economically advantageous and improve the efficiency of resource allocation by enabling the tailoring of clinical management to the specific needs and circumstances of the frail individual^41^. Notably, the consistency of these associations across diverse demographic groups supports the universal applicability of this combined assessment strategy.

There are several strengths of our study. First, we leveraged two larger and nationally representative cohorts with long-term follow-up periods. Findings in our study may be more generalizability and reliability. Second, we used three distinct data to assess FI based on self-reported items, blood biomarkers and their combination, which may demonstrate the robust results. Last, we conducted multiple methods to validate our findings, including stratified analyses in subgroups, sensitivity analyses in different cutoff for NT-proBNP. Limitations should also be acknowledged. Despite comprehensive adjustment for confounders, potential residual confounding may remain. Owing to the limited availability of NT-proBNP, this study was restricted to NHANES and HRS, and validation in more diverse populations is needed. Additionally, we only utilized baseline NT-proBNP data and FI values, lacking research on the longitudinal variation trajectory. Finally, recall bias may exist due to self-reported items related to frailty assessment

## Conclusion

In this cohort study, we found that the co-occurrence of frailty and HS is common, and jointly associated with increased mortality risk in the general population. These findings highlight the potential clinical value of integrating HS into frailty assessment to refine mortality risk stratification and better inform targeted intervention strategies.

## Data Availability

The datasets analyzed in the study are publicly available after registration at https://hrs.isr.umich.edu/, and https://www.cdc.gov/nchs/nhanes/index.html. The datasets generated and analyzed during the current study are available from the corresponding author on reasonable request.

## Acknowledgements

The data and samples used for this research were obtained from the HRS and NHANES. We would like to thank the workers, researchers, and participants involved in these cohorts.

## Ethics approval and consent to participate

The HRS is sponsored by the National Institute on Aging (grant numbers NIA U01AG009740 and NIA R01AG073289) and is conducted by the University of Michigan. Written informed consent was obtained by HRS investigators from all participants or their proxy respondents. The NHANES was approved by the National Center for Health Statistics (NCHS) Research Ethics Review Board, and all participants provided written informed consent

## Authors’ contributions

Zhang H, Huang Y, Hao M, and Li Y designed and conducted the research. Huang Y, Tang Y, Hao M, Hu Z, Jiang S, Han L, and Zhang H, performed acquisition, analysis, or interpretation of data. Huang Y drafted the manuscript. Zhang H, Hao M, and Li Y supervised the whole study. Zhang H, Huang Y, and Li Y had primary responsibility for the final content, and all authors read and approved the final manuscript.

**eTable 1. Items included in FI-Self-reported in NHANES. eTable 2. Items included in FI-Self-reported in HRS. eTable 3. Items included in FI-Lab in NHANES and HRS.**

**eTable 4. Association of frailty and heart stress with risk of all-cause mortality in NHANES and HRS.**

**eTable 5. Association of frailty and heart stress with risk of cause-specific mortality in NHANES.**

**eTable 6. Association of frailty assessed by FI-Self-report and heart stress with risk of all-cause mortality in subgroups.**

**eTable 7. Association of frailty assessed by FI-Lab and heart stress with risk of all-cause mortality in subgroups.**

**eTable 8. Association of frailty assessed by FI-Combined and heart stress with risk of all-cause mortality in subgroups.**

**eTable 9. Association of frailty and heart stress with risk of CVD mortality in subgroups in NHANES.**

**eTable 10. Association of frailty and heart stress with risk of cancer mortality in subgroups in NHANES.**

**eTable 11. Association of frailty and heart stress re-defined by a fixed cutoff with risk of all-cause mortality in NHANES and HRS.**

**eTable 12. Association of frailty and heart stress re-defined by a fixed cutoff with risk of cause-specific mortality in NHANES.**

**eTable 13. Association of frailty assessed by FI-Self-report and heart stress re-defined by a fixed cutoff with risk of all-cause mortality in subgroups.**

**eTable 14. Association of frailty assessed by FI-Lab and heart stress re-defined by a fixed cutoff with risk of all-cause mortality in subgroups.**

**eTable 15. Association of frailty assessed by FI-Combined and heart stress re-defined by a fixed cutoff with risk of all-cause mortality in subgroups.**

**eTable 16. Association of frailty and heart stress re-defined by a fixed cutoff with risk of CVD mortality in subgroups in NHANES.**

**eTable 17. Association of frailty and heart stress re-defined by a fixed cutoff with risk of cancer mortality in subgroups in NHANES.**

**eTable 18. Association of frailty and heart stress with risk of all-cause mortality in NHANES and HRS after excluding individuals who died within 2 years post-baseline.**

**eTable 19. Association of frailty and heart stress with risk of cause-specific mortality in NHANES after excluding individuals who died within 2 years post-baseline.**

**eFigure 1. Flowchart of the study.** NHANES indicates US National Health and Nutrition Examination Survey; HRS, Health and Retirement Study; NT-proBNP, N-terminal pro-B-type natriuretic peptide.

**eFigure 2. Cause-specific mortality stratified by combined frailty and HS.** Kaplan-Meier plots demonstrate survival probabilities according to baseline frailty and HS status across 4 groups in the NHANES. Cause-specific mortality outcomes includes CVD, heart disease, cerebrovascular disease, cancer and other causes. HS: heart stress. NHANES: National Health and Nutrition Examination Survey. CVD: cardiovascular disease.

**eFigure 3. Association between baseline FI and cause-specific mortality stratified by HS status.** Restricted cubic spline analyses show estimated hazard ratios for cause-specific mortality across continuous levels of FI in the NHANES. Cause-specific mortality outcomes includes CVD, heart disease, cerebrovascular disease, cancer and other causes. Models were adjusted for age, gender, race/ethnicity, education, body mass index, smoking status, physical activity, and marital status. FI: frailty index. HS: heart stress. NHANES: National Health and Nutrition Examination Survey. CVD: cardiovascular disease.

**eFigure 4. All-cause mortality stratified by combined frailty and HS re-defined by a fixed cutoff in sensitivity analyses.** HS was re-defined by applying a fixed cutoff of NT-proBNP. Kaplan-Meier plots demonstrate survival probabilities according to baseline frailty and HS status across 4 groups in the NHANES (A) and the HRS (B) cohorts. HS: heart stress. NT-proBNP: N-terminal pro-B-type natriuretic peptide. NHANES: National Health and Nutrition Examination Survey. HRS, Health and Retirement Study.

**eFigure 5. Cause-specific mortality stratified by combined frailty and HS re-defined by a fixed cutoff in sensitivity analyses.** HS was re-defined by applying a fixed cutoff of NT-proBNP. Kaplan-Meier plots demonstrate survival probabilities according to baseline frailty and HS status across 4 groups in the NHANES. Cause-specific mortality outcomes includes CVD, heart disease, cerebrovascular disease, cancer and other causes. HS: heart stress. NT-proBNP: N-terminal pro-B-type natriuretic peptide. NHANES: National Health and Nutrition Examination Survey. CVD, cardiovascular disease.

**eFigure 6. All-cause mortality stratified by combined frailty and HS after excluding early deaths in sensitivity analyses.** Individuals who died within 2 years after baseline were excluded from analyses. Kaplan-Meier plots demonstrate survival probabilities according to baseline frailty and HS status across 4 groups in the NHANES (A) and HRS (B) cohorts. HS: heart stress. NHANES: National Health and Nutrition Examination Survey. HRS, Health and Retirement Study.

**eFigure 7. Cause-specific mortality stratified by combined frailty and HS after excluding early deaths in sensitivity analyses.** Individuals who died within 2 years after baseline were excluded from analyses. Kaplan-Meier plots demonstrate survival probabilities according to baseline frailty and HS status across 4 groups in the NHANES. Cause-specific mortality outcomes includes CVD, heart disease, cerebrovascular disease, cancer and other causes. HS, heart stress. NHANES, US National Health and Nutrition Examination Survey. CVD, cardiovascular disease.

**eFigure 8. Association between baseline FI and all-cause mortality stratified by HS status re-defined by a fixed cutoff in sensitivity analyses.** HS was re-defined by applying a fixed cutoff of NT-proBNP. Restricted cubic spline analyses show estimated hazard ratios for all-cause mortality across continuous levels of FI in the NHANES (A) and the HRS (B) cohorts. Models were adjusted for age, gender, race/ethnicity, education, BMI, smoking status, physical activity, and marital status. FI, frailty index. HS, heart stress. NT-proBNP, N-terminal pro-B-type natriuretic peptide. NHANES, US National Health and Nutrition Examination Survey. HRS, Health and Retirement Study.

**eFigure 9. Association between baseline FI and cause-specific mortality stratified by HS status re-defined by a fixed cutoff in sensitivity analyses.** HS was re-defined by applying a fixed cutoff of NT-proBNP. Restricted cubic spline analyses show estimated hazard ratios for cause-specific mortality across continuous levels of FI in the NHANES. Cause-specific mortality outcomes includes CVD, heart disease, cerebrovascular disease, cancer and other causes. Models were adjusted for age, gender, race/ethnicity, education, body mass index, smoking status, physical activity, and marital status. FI, frailty index. HS, heart stress. NT-proBNP, N-terminal pro-B-type natriuretic peptide. NHANES, US National Health and Nutrition Examination Survey. CVD, cardiovascular disease.

**eFigure 10. Association between baseline FI and all-cause mortality stratified by HS status after excluding early deaths in sensitivity analyses.** Individuals who died within 2 years after baseline were excluded from analyses. Restricted cubic spline analyses show estimated hazard ratios for all-cause mortality across continuous levels of FI in the NHANES (A) and the HRS (B) cohorts. Models were adjusted for age, gender, race/ethnicity, education, body mass index, smoking status, physical activity, and marital status. FI, frailty index. HS, heart stress. NHANES, US National Health and Nutrition Examination Survey. HRS, Health and Retirement Study.

**eFigure 11. Association between baseline FI and cause-specific mortality stratified by HS status after excluding early deaths in sensitivity analyses.** Individuals who died within 2 years after baseline were excluded from analyses. Restricted cubic spline analyses show estimated hazard ratios for cause-specific mortality across continuous levels of FI in the NHANES. Cause-specific mortality outcomes includes CVD, heart disease, cerebrovascular disease, cancer and other causes. Models were adjusted for age, gender, race/ethnicity, education, body mass index, smoking status, physical activity, and marital status. FI, frailty index. HS, heart stress. NHANES, US National Health and Nutrition Examination Survey. CVD, cardiovascular disease.

