## Supplementary materials for "Heart Stress, Frailty and Mortality Risk in two prospective cohorts"

**Supplementary Online Content**

**eTable 1.** Items included in FI-Self-reported in NHANES.

**eTable 2.** Items included in FI-Self-reported in HRS.

**eTable 3.** Items included in FI-Lab in NHANES and HRS.

**eTable 4.** Association of frailty and heart stress with risk of all-cause mortality in NHANES and HRS.

**eTable 5.** Association of frailty and heart stress with risk of cause-specific mortality in NHANES.

**eTable 6.** Association of frailty assessed by FI-Self-report and heart stress with risk of all-cause mortality in subgroups.

**eTable 7.** Association of frailty assessed by FI-Lab and heart stress with risk of all-cause mortality in subgroups.

**eTable 8.** Association of frailty assessed by FI-Combined and heart stress with risk of all-cause mortality in subgroups.

**eTable 9.** Association of frailty and heart stress with risk of CVD mortality in subgroups in NHANES.

**eTable 10.** Association of frailty and heart stress with risk of cancer mortality in subgroups in NHANES.

**eTable 11.** Association of frailty and heart stress re-defined by a fixed cutoff with risk of all-cause mortality in NHANES and HRS.

**eTable 12.** Association of frailty and heart stress re-defined by a fixed cutoff with risk of cause-specific mortality in NHANES.

**eTable 13.** Association of frailty assessed by FI-Self-report and heart stress re-defined by a fixed cutoff with risk of all-cause mortality in subgroups.

**eTable 14.** Association of frailty assessed by FI-Lab and heart stress re-defined by a fixed cutoff with risk of all-cause mortality in subgroups.

**eTable 15.** Association of frailty assessed by FI-Combined and heart stress re-defined by a fixed cutoff with risk of all-cause mortality in subgroups.

**eTable 16.** Association of frailty and heart stress re-defined by a fixed cutoff with risk of CVD mortality in subgroups in NHANES.

**eTable 17.** Association of frailty and heart stress re-defined by a fixed cutoff with risk of cancer mortality in subgroups in NHANES.

**eTable 18.** Association of frailty and heart stress with risk of all-cause mortality in NHANES and HRS after excluding individuals who died within 2 years post-baseline.

**eTable 19.** Association of frailty and heart stress with risk of cause-specific mortality in NHANES after excluding individuals who died within 2 years post-baseline.

**eFigure 1.** Flowchart of the study.

**eFigure 2.** Cause-specific mortality stratified by combined frailty and HS.

**eFigure 3.** Association between baseline FI and cause-specific mortality stratified by HS status.

**eFigure 4.** All-cause mortality stratified by combined frailty and HS re-defined by a fixed cutoff in sensitivity analyses.

**eFigure 5.** Cause-specific mortality stratified by combined frailty and HS re-defined by a fixed cutoff in sensitivity analyses.

**eFigure 6.** All-cause mortality stratified by combined frailty and HS after excluding early deaths in sensitivity analyses.

**eFigure 7.** Cause-specific mortality stratified by combined frailty and HS after excluding early deaths in sensitivity analyses.

**eFigure 8.** Association between baseline FI and all-cause mortality stratified by HS status re-defined by a fixed cutoff in sensitivity analyses.

**eFigure 9.** Association between baseline FI and cause-specific mortality stratified by HS status re-defined by a fixed cutoff in sensitivity analyses.

**eFigure 10.** Association between baseline FI and all-cause mortality stratified by HS status after excluding early deaths in sensitivity analyses.

**eFigure 11.** Association between baseline FI and cause-specific mortality stratified by HS status after excluding early deaths in sensitivity analyses

| eTable 1. Items included in FI-Self-reported in NHANES. | | | |
| --- | --- | --- | --- |
|  | **Code** | **Description** | **Encoding** |
| 1 | AUQ130 | General condition of hearing | 0: Good or excellent, 0.5: Little or moderate trouble, 1: Lot of trouble or deaf |
| 2 | HUQ010 | General health condition | 0: Excellent, 0.25: Very good, 0.5: Good, 0.75: Fair, 1: Poor |
| 3 | HUQ020 | Health now compared with 1 year ago | 0: better or same, 1: worse |
| 4 | OSQ010a | Broken or fractured a hip | 0: no, 1: yes |
| 5 | PFQ056/PFQ056/PFQ057 | Experience confusion/memory problems | 0: no, 1: yes |
| 6 | PFQ060a/PFQ060A/PFQ061A | Difficulty managing money | 0: no difficulty, 0.33: Some difficulty, 0.66: much difficulty, 1: unable to do |
| 7 | PFQ060b/PFQ060B/PFQ061B | Difficulty walking a quarter mile | 0: no difficulty, 0.33: Some difficulty, 0.66: much difficulty, 1: unable to do |
| 8 | PFQ060c/PFQ060C/PFQ061C | Difficulty walking 10 steps | 0: no difficulty, 0.33: Some difficulty, 0.66: much difficulty, 1: unable to do |
| 9 | PFQ060d/PFQ060D/PFQ061D | Difficulty kneeling/crouching | 0: no difficulty, 0.33: Some difficulty, 0.66: much difficulty, 1: unable to do |
| 10 | PFQ060e/PFQ060E/PFQ061E | Difficulty lifting or carrying | 0: no difficulty, 0.33: Some difficulty, 0.66: much difficulty, 1: unable to do |
| 11 | PFQ060f/PFQ060F/PFQ061F | Household chore difficulty | 0: no difficulty, 0.33: Some difficulty, 0.66: much difficulty, 1: unable to do |
| 12 | PFQ060g/PFQ060G/PFQ061G | Difficulty preparing meals | 0: no difficulty, 0.33: Some difficulty, 0.66: much difficulty, 1: unable to do |
| 13 | PFQ060h/PFQ060H/PFQ061H | Difficulty walking between rooms on same floor | 0: no difficulty, 0.33: Some difficulty, 0.66: much difficulty, 1: unable to do |
| 14 | PFQ060i/PFQ060I/PFQ061I | Difficulty standing from armless chair | 0: no difficulty, 0.33: Some difficulty, 0.66: much difficulty, 1: unable to do |
| 15 | PFQ060j/PFQ060J/PFQ061J | Difficulty getting in/out of bed | 0: no difficulty, 0.33: Some difficulty, 0.66: much difficulty, 1: unable to do |
| 16 | PFQ060k/PFQ060K/PFQ061K | Difficulty using fork, knife, drinking from cup | 0: no difficulty, 0.33: Some difficulty, 0.66: much difficulty, 1: unable to do |
| 17 | PFQ060l/PFQ060L/PFQ061L | Difficulty dressing self | 0: no difficulty, 0.33: Some difficulty, 0.66: much difficulty, 1: unable to do |
| 18 | PFQ060m/PFQ060M/PFQ061M | Difficulty standing for long periods | 0: no difficulty, 0.33: Some difficulty, 0.66: much difficulty, 1: unable to do |
| 19 | PFQ060n/PFQ060N/PFQ061N | Difficulty sitting for long periods | 0: no difficulty, 0.33: Some difficulty, 0.66: much difficulty, 1: unable to do |
| 20 | PFQ060o/PFQ060O/PFQ061O | Difficulty reaching over head | 0: no difficulty, 0.33: Some difficulty, 0.66: much difficulty, 1: unable to do |
| 21 | PFQ060p/PFQ060P/PFQ061P | Difficulty grasping/holding small objects | 0: no difficulty, 0.33: Some difficulty, 0.66: much difficulty, 1: unable to do |
| 22 | PFQ060q/PFQ060Q/PFQ061Q | Difficulty going to movies, events | 0: no difficulty, 0.33: Some difficulty, 0.66: much difficulty, 1: unable to do |
| 23 | PFQ060r/PFQ060R/PFQ061R | Difficulty attending social events | 0: no difficulty, 0.33: Some difficulty, 0.66: much difficulty, 1: unable to do |
| 24 | PFQ060s/PFQ060S/PFQ061S | Difficulty leisure at home | 0: no difficulty, 0.33: Some difficulty, 0.66: much difficulty, 1: unable to do |
| 25 | RDQ030/RDD030/RDQ031 | Coughing most days - over 3 month period | 0: no, 1: yes |
| 26 | VIQ030/VIQ030/VIQ031 | General condition of eyesight | 0: Excellent, 0.25: Good, 0.5: Fair, 0.75: Poor, 1: Very poor |
| 27 | VIQ050C/VIQ050C/VIQ051C | Difficulty seeing steps/curbs - dim light | 0: no difficulty, 0.33: a little difficulty, 0.66: moderate difficulty, 1: extreme difficulty or unable to do because of eyesight |
| 28 | MCQ220 | Cancer | 0: no, 1: yes |
| 29 | MCQ160A | Arthritis | 0: no, 1: yes |
| 30 | DIQ010 | Diabetes | 0: no, 0.5: borderline, 1: yes |
| 31 | BPQ020 | High blood pressure | 0: no, 1: yes |
| 32 | HUQ050 | Frequency of healthcare use | 0: no, 0.5: 1–4, 1: ≥5 |
| 33 | HUQ070/HUD070/HUQ071 | Overnight hospital stays | 0: no, 1: yes |
| 34 | MCQ160F | Stroke | 0: no, 1: yes |

| **eTable 2.** Items included in FI-Self-reported in HRS. | | | |
| --- | --- | --- | --- |
|  | **Code** | **Description** | **Encoding** |
| 1 | RwARMSA | Difficulty reaching/extending arms up | 0: no, 1: yes/any |
| 2 | RwARTHRE | Ever diagnosed with arthritis | 0: no, 1: yes |
| 3 | RwBATHA | Difficulty bathing or showering | 0: no, 1: yes/any |
| 4 | RwBATHH | Gets help bathing, showering | 0: no, 1: yes |
| 5 | RwBEDA | Difficulty getting in/out of bed | 0: no, 1: yes/any |
| 6 | RwBEDE | Uses equipment to get in/out of bed | 0: no, 1: yes |
| 7 | RwBEDH | Gets help getting in/out of bed | 0: no, 1: yes |
| 8 | RwCANCRE | Ever diagnosed with cancer | 0: no, 1: yes |
| 9 | RwCHAIRA | Difficulty getting up from chair | 0: no, 1: yes/any |
| 10 | RwCLIM1A | Difficulty climbing one stair flight | 0: no, 1: yes/any |
| 11 | RwCLIMSA | Difficulty climbing several stair flights | 0: no, 1: yes/any |
| 12 | RwDIABE | Ever diagnosed with diabetes | 0: no, 1: yes |
| 13 | RwDIMEA | Difficulty picking up a dime | 0: no, 1: yes/any |
| 14 | RwDRESSA | Difficulty dressing | 0: no, 1: yes/any |
| 15 | RwDRESSH | Gets help dressing | 0: no, 1: yes |
| 16 | RwEATA | Difficulty eating | 0: no, 1: yes/any |
| 17 | RwEATH | Gets help eating | 0: no, 1: yes |
| 18 | RwHIBPE | Ever diagnosed with high blood pressure | 0: no, 1: yes |
| 19 | RwHOMCAR | Received home healthcare within previous 2 years | 0: no, 1: yes |
| 20 | RwHOSP | Had a hospital stay within previous 2 years | 0: no, 1: yes |
| 21 | RwLIFTA | Difficulty lifting/carrying 10 lbs | 0: no, 1: yes/any |
| 22 | RwLUNGE | Ever diagnosed with lung disease | 0: no, 1: yes |
| 23 | RwMONEYA | Difficulty managing money | 0: no, 1: yes/any |
| 24 | RwNHMLIV | Living in nursing home at time of interview | 0: no, 1: yes |
| 25 | RwNRSHOM | Had a nursing home stay within previous 2 years | 0: no, 1: yes |
| 26 | RwOUTPT | Had outpatient surgery within previous 2 years | 0: no, 1: yes |
| 27 | RwPHONEA | Difficulty using the telephone | 0: no, 1: yes/any |
| 28 | RwPUSHA | Difficulty pushing/pulling a large object | 0: no, 1: yes/any |
| 29 | RwSHLT | Self-reported health | 0: excellent-good, 1: fair-poor |
| 30 | RwSHOPA | Difficulty shopping for groceries | 0: no, 1: yes/any |
| 31 | RwSPCFAC | Visited a specialized health facility within previous 2 years | 0: no, 1: yes |
| 32 | RwSTOOPA | Difficulty stooping/kneeling/crouching | 0: no, 1: yes/any |
| 33 | RwSTROKE | Ever diagnosed with a stroke | 0: no, 1: yes |
| 34 | RwTOILTA | Difficulty using the toilet | 0: no, 1: yes/any |
| 35 | RwTOILTH | Gets help using the toilet | 0: no, 1: yes |
| 36 | RwWALK1A | Difficulty walking one block | 0: no, 1: yes/any |
| 37 | RwWALKRA | Difficulty walking across rooms | 0: no, 1: yes/any |
| 38 | RwWALKRE | Needs equipment to walk across rooms | 0: no, 1: yes |
| 39 | RwWALKRH | Gets help walking across rooms | 0: no, 1: yes |
| 40 | RwWALKSA | Difficulty walking several blocks | 0: no, 1: yes/any |

| **eTable 3.** Items included in FI-Lab in NHANES and HRS. | | | | |
| --- | --- | --- | --- | --- |
|  | **Code in NHANES** | **Code in HRS** | **Description** | **Encoding** |
| 1 | LBDSALSI | PALB | Albumin | 0: 32–45 g/L, 1: <32 or >45 g/L |
| 2 | LBXSAPSI/LBDSAPSI/LBXSAPSI | PALKP2 | Alkaline phosphatase | 0: 20–130 U/L, 1: <20 or >130 U/L |
| 3 | LBXSC3SI | PCO2 | Bicarbonate | 0: 21–28 mmol/L, 1: <21 or >28 mmol/L |
| 4 | LBDSTBSI | PBILT | Total bilirubin | 0: 2–21 μmol/L, 1: <2 or >21 μmol/L |
| 5 | LBDSBUSI | PBUN | Blood urea nitrogen | 0: 2.9–8.2 mmol/L, 1: <2.9 or >8.2 mmol/L |
| 6 | LBXCRP | PCRP | C-reactive protein | 0: 0–1 mg/dL, 1: >1 mg/dL |
| 7 | LBDSCRSI | PCR | Creatinine | Males: 0: 60–110 μmol/L, 1: <60 or >110 μmol/L; Females: 0: 45–90 μmol/L, 1: <45 or >90 μmol/L |
| 8 | LBDHDLSI/LBDHDLSI/LBDHDDSI | PHDLD | Direct HDL cholesterol | 0: ≥1.3 mmol/L, 1: <1.3 mmol/L |
| 9 | LBDSGLSI | PGLUFF | Serum glucose | 0: 3.9–6.1 mmol/L, 1: <3.9 or >6.1 mmol/L |
| 10 | LBXHGB | PHGB | Hemoglobin | Males: 0: 13.5–18 g/dL, 1: <13.5 or >18 g/dL; Females: 0: 12–16 g/dL, 1: <12 or >16 g/dL |
| 11 | LBXMCVSI | PMCV | Mean cell volume | 0: 80–96 fL, 1: <80 or >96 fL |
| 12 | LBXPLTSI | PPLT | Platelet count | 0: 150–450 ×10³/μL, 1: <150 or >450 ×10³/μL |
| 13 | LBDSTPSI | PTP | Total protein | 0: 60–78 g/L, 1: <60 or >78 g/L |
| 14 | LBXRDW | PRDW | Red cell distribution width | 0: 11.6–14.6 %, 1: <11.6 or >14.6 % |
| 15 | LBXNEPCT | PNEUT | Segmented neutrophils percent | 0: 40–80 %, 1: <40 or >80 % |
| 16 | LBXSNASI | PNA | Sodium | 0: 136–142 mmol/L, 1: <136 or >142 mmol/L |
| 17 | LBDSCASI | PCA | Total calcium | 0: 2.30–2.74 mmol/L, 1: <2.30 or >2.74 mmol/L |
| 18 | LBDSCHSI | PCHOL | Total cholesterol | 0: 3.88–6.47 mmol/L, 1: <3.88 or >6.47 mmol/L |
| 19 | LBDSTRSI | PTGF | Triglycerides | 0: 0.11–2.74 mmol/L, 1: <0.11 or >2.74 mmol/L |
| 20 | LBXHCT | PHCT | Hematocrit | Males: 0: 0.41–0.53 L/L, 1: <0.41 or >0.53 L/L; Females: 0: 0.36–0.46 L/L, 1: <0.36 or >0.46 L/L |
| 21 | LBXLYPCT | PLYMP | Lymphocytes | 0: 22–44 %, 1: <22 or >44 % |
| 22 | LBXMCHSI | PMCH | Mean corpuscular hemoglobin | 0: 26–34 pg, 1: <26 or >34 pg |
| 23 | LBXMOPCT | PMONO | Monocytes | 0: ≤8%, 1: >8% |
| 24 | LBXRBCSI | PRBC | Red blood cells | Males: 0: 4.5–5.9 ×10¹²/L, 1: <4.5 or >5.9 ×10¹²/L; Females: 0: 4.0–5.2 ×10¹²/L, 1: <4.0 or >5.2 ×10¹²/L |
| 25 | LBXWBCSI | PWBC | White blood cells | 0: 1.8–7.8 ×10⁹/L, 1: <1.8 or >7.8 ×10⁹/L |
| 26 | LBXMPSI | PMPV | Mean platelet volume | 0: 7–13 fL, 1: <7 or >13 fL |
| 27 | LBDFERSI | PFERTN | Ferritin | Males: 0: 20–250 μg/L, 1: <20 or >250 μg/L; Females: 0: 10–120 μg/L, 1: <10 or >120 μg/L |
| 28 | LBXSKSI | PK | Potassium | 0: 3.8–5.0 mmol/L, 1: <3.8 or >5.0 mmol/L |
| 29 | LBXSATSI | PALT | Alanine transaminase | 0: 10–50 U/L, 1: <10 or >50 U/L |
| 30 | LBXSASSI | PAST | Aspartate aminotransferase | 0: 8–33 U/L, 1: <8 or >33 U/L |
| 31 | LBDBANO | PABAS | Basophils | 0: 0–0.1 ×10⁹/L, 1: >0.1 ×10⁹/L |
| 32 | LBDEONO | PAEOS | Eosinophils | 0: 0–0.4 ×10⁹/L, 1: >0.4 ×10⁹/L |

| **eTable 4.** Association of frailty and heart stress with risk of all-cause mortality in NHANES and HRS. | | | | | | | | |
| --- | --- | --- | --- | --- | --- | --- | --- | --- |
| **A.** FI-Self-report | | | | | | | | |
|  |  |  | **Model 1** | | **Model 2** | | **Model 3** | |
|  |  | **Event/N (%)** | **HR (95%CI)** | **P-value** | **HR 95%CI** | **P-value** | **HR (95%CI)** | **P-value** |
| **NHANES** |  |  |  |  |  |  |  |  |
| No Frailty | No Heart Stress | 1310/3128 (41.88) | Ref |  | Ref |  | Ref |  |
|  | Heart Stress | 925/1352 (68.42) | 2.28 (2.10-2.48) | 1.18E-81 | 1.89 (1.73-2.05) | 3.97E-47 | 1.88 (1.73-2.05) | 8.40E-47 |
| Frailty | No Heart Stress | 404/685 (58.98) | 1.76 (1.58-1.97) | 2.83E-23 | 2.02 (1.81-2.26) | 2.17E-34 | 2.06 (1.84-2.31) | 3.34E-35 |
|  | Heart Stress | 470/556 (84.53) | 4.35 (3.91-4.84) | 1.21E-159 | 3.57 (3.19-4.00) | 2.03E-108 | 3.58 (3.20-4.01) | 1.61E-107 |
| **HRS** |  |  |  |  |  |  |  |  |
| No Frailty | No Heart Stress | 511/5575 (9.17) | Ref |  | Ref |  | Ref |  |
|  | Heart Stress | 478/1913 (24.99) | 2.96 (2.61-3.35) | 3.34E-65 | 2.02 (1.78-2.29) | 7.47E-27 | 1.99 (1.75-2.27) | 6.06E-26 |
| Frailty | No Heart Stress | 252/1086 (23.20) | 2.74 (2.36-3.19) | 3.25E-39 | 2.46 (2.11-2.87) | 7.20E-31 | 2.29 (1.95-2.68) | 6.02E-24 |
|  | Heart Stress | 458/914 (50.11) | 7.41 (6.53-8.40) | 3.79E-211 | 4.91 (4.29-5.62) | 3.43E-118 | 5.02 (4.38-5.76) | 1.21E-116 |
| **B.** FI-Lab | | | | | | | | |
|  |  |  | **Model 1** | | **Model 2** | | **Model 3** | |
|  |  | **Event/N (%)** | **HR (95%CI)** | **P-value** | **HR 95%CI** | **P-value** | **HR (95%CI)** | **P-value** |
| **NHANES** |  |  |  |  |  |  |  |  |
| No Frailty | No Heart Stress | 1813/8607 (21.06) | Ref |  | Ref |  | Ref |  |
|  | Heart Stress | 1109/2350 (47.19) | 2.83 (2.63-3.05) | 1.28E-163 | 1.81 (1.67-1.95) | 2.19E-50 | 1.81 (1.67-1.96) | 1.51E-50 |
| Frailty | No Heart Stress | 235/770 (30.52) | 1.54 (1.34-1.76) | 4.63E-10 | 1.92 (1.67-2.20) | 8.38E-21 | 1.91 (1.67-2.19) | 1.78E-20 |
|  | Heart Stress | 362/525 (68.95) | 5.79 (5.17-6.49) | 9.63E-203 | 3.39 (3.01-3.81) | 1.79E-91 | 3.43 (3.04-3.86) | 2.26E-92 |
| **HRS** |  |  |  |  |  |  |  |  |
| No Frailty | No Heart Stress | 534/5736 (9.31) | Ref |  | Ref |  | Ref |  |
|  | Heart Stress | 465/1924 (24.17) | 2.81 (2.48-3.18) | 1.53E-59 | 1.86 (1.64-2.12) | 2.60E-21 | 1.86 (1.64-2.11) | 3.03E-21 |
| Frailty | No Heart Stress | 209/755 (27.68) | 3.34 (2.84-3.91) | 2.64E-49 | 2.56 (2.18-3.02) | 1.16E-29 | 2.44 (2.07-2.88) | 3.05E-26 |
|  | Heart Stress | 429/827 (51.87) | 7.68 (6.76-8.72) | 1.69E-215 | 4.88 (4.27-5.59) | 9.52E-119 | 4.73 (4.13-5.41) | 1.87E-112 |
| **C.** FI-Combined | | | | | | | | |
|  |  |  | **Model 1** | | **Model 2** | | **Model 3** | |
|  |  | **Event/N (%)** | **HR (95%CI)** | **P-value** | **HR 95%CI** | **P-value** | **HR (95%CI)** | **P-value** |
| **NHANES** |  |  |  |  |  |  |  |  |
| No Frailty | No Heart Stress | 1434/3387 (42.34) | Ref |  | Ref |  | Ref |  |
|  | Heart Stress | 977/1440 (67.85) | 2.22 (2.05-2.41) | 7.59E-82 | 1.82 (1.68-1.98) | 4.64E-45 | 1.82 (1.67-1.98) | 6.96E-45 |
| Frailty | No Heart Stress | 279/418 (66.75) | 2.22 (1.96-2.53) | 3.24E-34 | 2.57 (2.26-2.92) | 2.90E-46 | 2.64 (2.31-3.01) | 3.79E-47 |
|  | Heart Stress | 424/476 (89.08) | 5.29 (4.73-5.90) | 1.13E-191 | 4.11 (3.66-4.61) | 5.29E-127 | 4.15 (3.70-4.67) | 1.47E-126 |
| **HRS** |  |  |  |  |  |  |  |  |
| No Frailty | No Heart Stress | 517/5690 (9.09) | Ref |  | Ref |  | Ref |  |
|  | Heart Stress | 441/1892 (23.31) | 2.75 (2.42-3.12) | 8.11E-55 | 1.86 (1.63-2.12) | 1.12E-20 | 1.85 (1.62-2.10) | 3.96E-20 |
| Frailty | No Heart Stress | 238/869 (27.39) | 3.34 (2.86-3.89) | 2.42E-53 | 2.82 (2.42-3.29) | 3.09E-39 | 2.64 (2.24-3.11) | 1.76E-31 |
|  | Heart Stress | 470/891 (52.75) | 8.09 (7.13-9.17) | 4.65E-234 | 5.38 (4.71-6.15) | 3.43E-136 | 5.54 (4.84-6.35) | 6.86E-136 |

| **eTable 5.** Association of frailty and heart stress with risk of cause-specific mortality in NHANES. | | | | | | | | |
| --- | --- | --- | --- | --- | --- | --- | --- | --- |
| **A.** FI-Self-report | | | | | | | | |
|  |  |  | **Model 1** | | **Model 2** | | **Model 3** | |
|  |  | **Event/N (%)** | **HR (95%CI)** | **P-value** | **HR 95%CI** | **P-value** | **HR (95%CI)** | **P-value** |
| **Heart disease** |  |  |  |  |  |  |  |  |
| No Frailty | No Heart Stress | 278/2096 (13.26) |  |  |  |  |  |  |
|  | Heart Stress | 319/746 (42.76) | 4.12 (3.51-4.84) | 1.94E-66 | 2.99 (2.54-3.53) | 2.00E-38 | 3.00 (2.54-3.55) | 1.60E-38 |
| Frailty | No Heart Stress | 107/388 (27.58) | 2.38 (1.90-2.97) | 2.93E-14 | 2.99 (2.39-3.75) | 1.95E-21 | 2.76 (2.18-3.49) | 2.29E-17 |
|  | Heart Stress | 165/251 (65.74) | 9.07 (7.46-11.02) | 3.23E-109 | 7.17 (5.77-8.91) | 4.14E-71 | 7.11 (5.72-8.83) | 5.09E-70 |
| **Cerebrovascular disease** | |  |  |  |  |  |  |  |
| No Frailty | No Heart Stress | 75/1893 (3.96) |  |  |  |  |  |  |
|  | Heart Stress | 50/477 (10.48) | 2.79 (1.95-3.99) | 1.94E-08 | 2.16 (1.50-3.10) | 3.16E-05 | 2.08 (1.44-3.00) | 8.61E-05 |
| Frailty | No Heart Stress | 25/306 (8.17) | 2.14 (1.36-3.36) | 1.00E-03 | 2.57 (1.63-4.05) | 4.96E-05 | 2.34 (1.45-3.76) | 4.80E-04 |
|  | Heart Stress | 26/112 (23.21) | 6.78 (4.34-10.59) | 4.36E-17 | 4.19 (2.53-6.92) | 2.45E-08 | 3.72 (2.23-6.23) | 5.21E-07 |
| **CVD** |  |  |  |  |  |  |  |  |
| No Frailty | No Heart Stress | 353/2171 (16.26) |  |  |  |  |  |  |
|  | Heart Stress | 369/796 (46.36) | 3.70 (3.20-4.28) | 8.39E-69 | 2.68 (2.31-3.12) | 9.85E-38 | 2.69 (2.31-3.12) | 8.50E-38 |
| Frailty | No Heart Stress | 132/413 (31.96) | 2.28 (1.86-2.78) | 7.97E-16 | 2.75 (2.25-3.37) | 1.16E-22 | 2.71 (2.20-3.34) | 4.19E-21 |
|  | Heart Stress | 191/277 (68.95) | 7.93 (6.63-9.47) | 5.06E-115 | 5.98 (4.91-7.28) | 7.40E-71 | 5.91 (4.85-7.20) | 2.74E-69 |
| **Cancer** |  |  |  |  |  |  |  |  |
| No Frailty | No Heart Stress | 304/2122 (14.33) |  |  |  |  |  |  |
|  | Heart Stress | 157/584 (26.88) | 2.10 (1.73-2.54) | 4.69E-14 | 1.91 (1.57-2.32) | 7.37E-11 | 1.86 (1.53-2.26) | 5.72E-10 |
| Frailty | No Heart Stress | 90/371 (24.26) | 1.81 (1.43-2.29) | 7.90E-07 | 2.15 (1.70-2.73) | 2.31E-10 | 2.24 (1.76-2.86) | 6.84E-11 |
|  | Heart Stress | 65/151 (43.05) | 4.10 (3.14-5.36) | 6.18E-25 | 3.90 (2.93-5.18) | 7.34E-21 | 3.64 (2.72-4.85) | 2.05E-18 |
| **Others** | |  |  |  |  |  |  |  |
| No Frailty | No Heart Stress | 653/2471 (26.43) |  |  |  |  |  |  |
|  | Heart Stress | 399/826 (48.31) | 2.25 (1.99-2.55) | 2.69E-37 | 1.82 (1.60-2.06) | 2.02E-20 | 1.80 (1.59-2.05) | 9.34E-20 |
| Frailty | No Heart Stress | 182/463 (39.31) | 1.74 (1.47-2.05) | 4.54E-11 | 2.05 (1.74-2.42) | 2.18E-17 | 2.02 (1.70-2.39) | 5.27E-16 |
|  | Heart Stress | 214/300 (71.33) | 4.81 (4.12-5.62) | 3.04E-87 | 3.60 (3.05-4.24) | 1.21E-52 | 3.40 (2.87-4.02) | 2.71E-46 |
| **B.** FI-Lab | | | | | | | | |
|  |  |  | **Model 1** | | **Model 2** | | **Model 3** | |
|  |  | **Event/N (%)** | **HR (95%CI)** | **P-value** | **HR 95%CI** | **P-value** | **HR (95%CI)** | **P-value** |
| **Heart disease** |  |  |  |  |  |  |  |  |
| No Frailty | No Heart Stress | 394/7188 (5.48) |  |  |  |  |  |  |
|  | Heart Stress | 383/1624 (23.58) | 4.90 (4.26-5.64) | 1.30E-108 | 2.77 (2.39-3.22) | 6.04E-42 | 2.79 (2.41-3.24) | 2.95E-42 |
| Frailty | No Heart Stress | 45/580 (7.76) | 1.39 (1.02-1.90) | 3.48E-02 | 2.05 (1.50-2.79) | 5.31E-06 | 2.13 (1.56-2.90) | 2.00E-06 |
|  | Heart Stress | 114/277 (41.16) | 10.07 (8.17-12.41) | 2.72E-104 | 5.01 (4.00-6.27) | 1.46E-44 | 5.18 (4.13-6.49) | 5.02E-46 |
| **Cerebrovascular disease** | |  |  |  |  |  |  |  |
| No Frailty | No Heart Stress | 110/6904 (1.59) |  |  |  |  |  |  |
|  | Heart Stress | 62/1303 (4.76) | 3.07 (2.25-4.19) | 1.72E-12 | 2.06 (1.50-2.83) | 7.92E-06 | 2.06 (1.50-2.84) | 8.11E-06 |
| Frailty | No Heart Stress | 8/543 (1.47) | 0.89 (0.44-1.83) | 7.57E-01 | 1.79 (0.87-3.67) | 1.14E-01 | 1.58 (0.74-3.41) | 2.39E-01 |
|  | Heart Stress | 20/183 (10.93) | 7.10 (4.41-11.44) | 7.53E-16 | 3.49 (2.10-5.79) | 1.49E-06 | 3.74 (2.26-6.18) | 2.74E-07 |
| **CVD** |  |  |  |  |  |  |  |  |
| No Frailty | No Heart Stress | 504/7298 (6.91) |  |  |  |  |  |  |
|  | Heart Stress | 445/1686 (26.39) | 4.41 (3.88-5.01) | 4.92E-115 | 2.50 (2.18-2.85) | 3.48E-41 | 2.51 (2.19-2.87) | 2.18E-41 |
| Frailty | No Heart Stress | 53/588 (9.01) | 1.28 (0.97-1.70) | 8.43E-02 | 1.89 (1.42-2.51) | 1.12E-05 | 1.85 (1.39-2.47) | 2.44E-05 |
|  | Heart Stress | 134/297 (45.12) | 9.05 (7.48-10.95) | 2.62E-113 | 4.32 (3.53-5.30) | 6.77E-45 | 4.50 (3.67-5.52) | 5.72E-47 |
| **Cancer** |  |  |  |  |  |  |  |  |
| No Frailty | No Heart Stress | 449/7243 (6.20) |  |  |  |  |  |  |
|  | Heart Stress | 188/1429 (13.16) | 2.25 (1.90-2.67) | 9.93E-21 | 1.78 (1.49-2.12) | 9.06E-11 | 1.77 (1.49-2.11) | 1.75E-10 |
| Frailty | No Heart Stress | 57/592 (9.63) | 1.56 (1.19-2.06) | 1.48E-03 | 2.32 (1.76-3.05) | 2.74E-09 | 2.31 (1.75-3.05) | 3.61E-09 |
|  | Heart Stress | 50/213 (23.47) | 4.38 (3.27-5.87) | 3.95E-23 | 3.59 (2.65-4.86) | 1.26E-16 | 3.71 (2.73-5.04) | 4.64E-17 |
| **Others** | |  |  |  |  |  |  |  |
| No Frailty | No Heart Stress | 860/7654 (11.24) |  |  |  |  |  |  |
|  | Heart Stress | 476/1717 (27.72) | 2.79 (2.50-3.12) | 3.46E-72 | 1.80 (1.60-2.02) | 2.37E-23 | 1.79 (1.60-2.01) | 6.08E-23 |
| Frailty | No Heart Stress | 125/660 (18.94) | 1.74 (1.44-2.10) | 6.92E-09 | 2.39 (1.98-2.88) | 1.45E-19 | 2.32 (1.92-2.81) | 2.17E-18 |
|  | Heart Stress | 178/341 (52.20) | 6.77 (5.76-7.96) | 7.96E-119 | 3.78 (3.19-4.48) | 2.80E-53 | 3.81 (3.21-4.51) | 2.17E-53 |
| **C.** FI-Combined | | | | | | | | |
|  |  |  | **Model 1** | | **Model 2** | | **Model 3** | |
|  |  | **Event/N (%)** | **HR (95%CI)** | **P-value** | **HR 95%CI** | **P-value** | **HR (95%CI)** | **P-value** |
| **Heart disease** |  |  |  |  |  |  |  |  |
| No Frailty | No Heart Stress | 323/2276 (14.19) |  |  |  |  |  |  |
|  | Heart Stress | 341/804 (42.41) | 3.82 (3.28-4.44) | 2.37E-66 | 2.80 (2.39-3.27) | 1.35E-37 | 2.82 (2.40-3.30) | 6.64E-38 |
| Frailty | No Heart Stress | 64/203 (31.53) | 2.66 (2.04-3.48) | 8.02E-13 | 3.37 (2.57-4.41) | 1.54E-18 | 3.13 (2.36-4.16) | 3.35E-15 |
|  | Heart Stress | 144/196 (73.47) | 10.70 (8.77-13.06) | 1.09E-120 | 7.37 (5.92-9.19) | 1.60E-70 | 7.46 (5.98-9.32) | 3.16E-70 |
| **Cerebrovascular disease** | |  |  |  |  |  |  |  |
| No Frailty | No Heart Stress | 85/2038 (4.17) |  |  |  |  |  |  |
|  | Heart Stress | 51/514 (9.92) | 2.50 (1.77-3.54) | 2.34E-07 | 1.94 (1.36-2.75) | 2.33E-04 | 1.85 (1.30-2.64) | 6.85E-04 |
| Frailty | No Heart Stress | 15/154 (9.74) | 2.42 (1.40-4.19) | 1.61E-03 | 3.24 (1.87-5.61) | 2.91E-05 | 2.81 (1.58-4.98) | 4.28E-04 |
|  | Heart Stress | 26/78 (33.33) | 9.93 (6.40-15.41) | 1.39E-24 | 5.43 (3.29-8.96) | 3.56E-11 | 5.41 (3.29-8.88) | 2.63E-11 |
| **CVD** |  |  |  |  |  |  |  |  |
| No Frailty | No Heart Stress | 408/2361 (17.28) |  |  |  |  |  |  |
|  | Heart Stress | 392/855 (45.85) | 3.43 (2.99-3.94) | 9.88E-68 | 2.51 (2.18-2.90) | 2.11E-36 | 2.52 (2.19-2.91) | 1.27E-36 |
| Frailty | No Heart Stress | 79/218 (36.24) | 2.57 (2.02-3.27) | 1.78E-14 | 3.13 (2.45-3.99) | 3.68E-20 | 3.06 (2.38-3.93) | 2.26E-18 |
|  | Heart Stress | 170/222 (76.58) | 9.40 (7.85-11.27) | 1.20E-129 | 6.21 (5.09-7.58) | 2.02E-72 | 6.29 (5.15-7.69) | 3.47E-72 |
| **Cancer** |  |  |  |  |  |  |  |  |
| No Frailty | No Heart Stress | 335/2288 (14.64) |  |  |  |  |  |  |
|  | Heart Stress | 171/634 (26.97) | 2.06 (1.72-2.48) | 1.28E-14 | 1.87 (1.55-2.25) | 4.49E-11 | 1.82 (1.51-2.20) | 3.38E-10 |
| Frailty | No Heart Stress | 59/198 (29.80) | 2.24 (1.70-2.96) | 1.08E-08 | 2.79 (2.11-3.69) | 4.78E-13 | 2.96 (2.22-3.96) | 1.55E-13 |
|  | Heart Stress | 51/103 (49.51) | 5.01 (3.73-6.73) | 9.12E-27 | 4.32 (3.16-5.90) | 4.41E-20 | 4.18 (3.05-5.73) | 6.57E-19 |
| **Others** | |  |  |  |  |  |  |  |
| No Frailty | No Heart Stress | 691/2644 (26.13) |  |  |  |  |  |  |
|  | Heart Stress | 414/877 (47.21) | 2.20 (1.95-2.49) | 9.31E-37 | 1.74 (1.54-1.97) | 2.29E-18 | 1.72 (1.52-1.95) | 1.05E-17 |
| Frailty | No Heart Stress | 141/280 (50.36) | 2.50 (2.09-3.00) | 3.14E-23 | 2.98 (2.48-3.57) | 1.29E-31 | 2.99 (2.48-3.61) | 1.87E-30 |
|  | Heart Stress | 203/255 (79.61) | 6.56 (5.60-7.68) | 7.72E-120 | 4.68 (3.95-5.53) | 4.56E-72 | 4.53 (3.82-5.38) | 5.41E-67 |

| **eTable 6.** Association of frailty assessed by FI-Self-report and heart stress with risk of all-cause mortality in subgroups. | | | | | | | | |
| --- | --- | --- | --- | --- | --- | --- | --- | --- |
| **Subgroups** | **No Heart Stress or Frailty** | **Heart Stress, no Frailty** | | **Frailty, no Heart Stress** | | **Frailty, and Heart Stress** | |  |
|  |  | **HR (95% CI)** | **P-value** | **HR (95% CI)** | **P-value** | **HR (95% CI)** | **P-value** | **P-int** |
| **NHANES** |  |  |  |  |  |  |  |  |
| Age group |  |  |  |  |  |  |  |  |
| 20-49 | 1.00 (Reference) | 3.30 (1.99-5.46) | 3.71E-06 | 3.23 (1.99-5.25) | 2.27E-06 | 4.90 (2.60-9.22) | 8.66E-07 | 2.27E-02 |
| 50-64 | 1.00 (Reference) | 2.30 (1.84-2.88) | 2.65E-13 | 2.11 (1.65-2.69) | 1.82E-09 | 5.24 (3.84-7.15) | 1.10E-25 |  |
| 65 years or older | 1.00 (Reference) | 1.81 (1.64-1.99) | 4.93E-34 | 1.84 (1.60-2.11) | 9.35E-18 | 3.18 (2.80-3.61) | 3.06E-71 |  |
| Gender |  |  |  |  |  |  |  |  |
| Male | 1.00 (Reference) | 2.01 (1.79-2.25) | 8.88E-32 | 2.04 (1.74-2.39) | 1.20E-18 | 3.78 (3.20-4.46) | 2.73E-55 | 2.17E-01 |
| Female | 1.00 (Reference) | 1.76 (1.55-2.01) | 4.91E-18 | 2.09 (1.77-2.47) | 3.47E-18 | 3.25 (2.78-3.81) | 3.36E-49 |  |
| Body mass index |  |  |  |  |  |  |  |  |
| Normal or underweight | 1.00 (Reference) | 1.64 (1.40-1.91) | 4.74E-10 | 2.21 (1.76-2.79) | 1.64E-11 | 3.00 (2.40-3.74) | 1.80E-22 | 1.73E-01 |
| Overweight | 1.00 (Reference) | 1.92 (1.66-2.21) | 2.06E-19 | 1.94 (1.58-2.37) | 1.60E-10 | 3.07 (2.48-3.81) | 7.26E-25 |  |
| Obese | 1.00 (Reference) | 1.95 (1.65-2.30) | 5.73E-15 | 1.63 (1.34-1.99) | 1.26E-06 | 3.65 (2.98-4.48) | 1.98E-35 |  |
| Race/Ethnicity |  |  |  |  |  |  |  |  |
| White/Caucasian | 1.00 (Reference) | 1.83 (1.64-2.03) | 2.55E-28 | 2.06 (1.76-2.41) | 4.16E-19 | 4.36 (3.79-5.03) | 7.26E-93 | 1.35E-02 |
| Black/African American | 1.00 (Reference) | 2.33 (1.86-2.91) | 1.29E-13 | 1.79 (1.36-2.35) | 2.58E-05 | 3.31 (2.52-4.35) | 8.55E-18 |  |
| Others | 1.00 (Reference) | 1.89 (1.55-2.30) | 1.58E-10 | 1.94 (1.56-2.42) | 3.60E-09 | 2.80 (2.20-3.56) | 3.68E-17 |  |
| Smoking status |  |  |  |  |  |  |  |  |
| Ever | 1.00 (Reference) | 1.90 (1.69-2.13) | 2.90E-28 | 1.90 (1.64-2.21) | 1.44E-17 | 3.52 (3.03-4.10) | 1.88E-59 | 1.37E-01 |
| Never | 1.00 (Reference) | 1.84 (1.61-2.10) | 3.39E-19 | 2.13 (1.77-2.56) | 1.50E-15 | 3.54 (2.98-4.20) | 1.34E-46 |  |
| Vigorous physical activity |  |  |  |  |  |  |  |  |
| Yes | 1.00 (Reference) | 1.49 (1.17-1.89) | 1.38E-03 | 1.33 (0.82-2.14) | 2.51E-01 | 4.25 (1.97-9.16) | 2.20E-04 | 8.61E-02 |
| No | 1.00 (Reference) | 1.94 (1.76-2.12) | 4.69E-44 | 2.07 (1.84-2.33) | 5.07E-33 | 3.52 (3.13-3.95) | 6.29E-99 |  |
| Marital status |  |  |  |  |  |  |  |  |
| Married/Living with partner | 1.00 (Reference) | 1.93 (1.72-2.17) | 4.11E-29 | 1.97 (1.68-2.31) | 1.67E-16 | 3.54 (2.98-4.19) | 1.10E-47 | 7.36E-01 |
| Others | 1.00 (Reference) | 1.75 (1.52-2.00) | 1.10E-15 | 1.92 (1.61-2.28) | 2.04E-13 | 3.23 (2.75-3.80) | 1.42E-46 |  |
| Educational status |  |  |  |  |  |  |  |  |
| Less than high school | 1.00 (Reference) | 1.87 (1.63-2.15) | 4.17E-19 | 1.82 (1.55-2.13) | 3.13E-13 | 2.97 (2.54-3.48) | 2.13E-41 | 3.26E-02 |
| High school | 1.00 (Reference) | 1.90 (1.60-2.26) | 2.05E-13 | 2.01 (1.55-2.59) | 8.70E-08 | 4.17 (3.26-5.33) | 4.75E-30 |  |
| College or above | 1.00 (Reference) | 1.82 (1.57-2.11) | 1.33E-15 | 2.19 (1.75-2.75) | 1.15E-11 | 3.93 (3.10-4.97) | 5.98E-30 |  |
| **HRS** |  |  |  |  |  |  |  |  |
| Age group |  |  |  |  |  |  |  |  |
| 50-64 | 1.00 (Reference) | 3.27 (2.31-4.62) | 1.98E-11 | 2.94 (2.02-4.29) | 1.98E-08 | 8.71 (6.03-12.58) | 8.96E-31 | 2.26E-02 |
| 65 years or older | 1.00 (Reference) | 1.86 (1.62-2.14) | 5.96E-19 | 2.15 (1.80-2.56) | 3.35E-17 | 4.59 (3.96-5.31) | 4.76E-91 |  |
| Gender |  |  |  |  |  |  |  |  |
| Male | 1.00 (Reference) | 2.29 (1.92-2.73) | 2.67E-20 | 2.23 (1.75-2.85) | 9.21E-11 | 5.22 (4.27-6.38) | 1.83E-58 | 7.07E-01 |
| Female | 1.00 (Reference) | 1.69 (1.40-2.04) | 4.38E-08 | 2.34 (1.89-2.90) | 9.00E-15 | 4.57 (3.79-5.52) | 2.11E-56 |  |
| Body mass index |  |  |  |  |  |  |  |  |
| Normal or underweight | 1.00 (Reference) | 2.11 (1.71-2.60) | 3.10E-12 | 2.93 (2.18-3.94) | 1.13E-12 | 5.68 (4.44-7.27) | 2.39E-43 | 4.83E-02 |
| Overweight | 1.00 (Reference) | 1.59 (1.27-1.99) | 4.54E-05 | 1.74 (1.28-2.35) | 3.50E-04 | 4.05 (3.20-5.12) | 2.95E-31 |  |
| Obese | 1.00 (Reference) | 2.35 (1.84-3.01) | 7.44E-12 | 2.28 (1.77-2.95) | 2.84E-10 | 4.47 (3.50-5.71) | 6.07E-33 |  |
| Race/Ethnicity |  |  |  |  |  |  |  |  |
| White/Caucasian | 1.00 (Reference) | 2.00 (1.73-2.30) | 4.77E-21 | 2.75 (2.29-3.31) | 4.36E-27 | 4.85 (4.13-5.71) | 1.43E-81 | 9.86E-01 |
| Black/African American | 1.00 (Reference) | 1.84 (1.31-2.60) | 4.90E-04 | 1.37 (0.95-1.98) | 9.13E-02 | 4.52 (3.32-6.16) | 1.04E-21 |  |
| Others | 1.00 (Reference) | 2.27 (1.31-3.95) | 3.55E-03 | 1.06 (0.53-2.12) | 8.75E-01 | 5.03 (2.94-8.61) | 3.79E-09 |  |
| Smoking status |  |  |  |  |  |  |  |  |
| Ever | 1.00 (Reference) | 2.03 (1.72-2.40) | 6.54E-17 | 2.47 (2.03-3.01) | 2.48E-19 | 4.82 (4.04-5.75) | 1.21E-68 | 1.86E-01 |
| Never | 1.00 (Reference) | 1.96 (1.60-2.40) | 1.32E-10 | 1.82 (1.38-2.41) | 2.60E-05 | 4.71 (3.77-5.90) | 1.01E-41 |  |
| Vigorous physical activity |  |  |  |  |  |  |  |  |
| Yes | 1.00 (Reference) | 2.27 (1.83-2.81) | 7.81E-14 | 2.31 (1.59-3.37) | 1.15E-05 | 4.38 (3.17-6.06) | 4.53E-19 | 8.53E-01 |
| No | 1.00 (Reference) | 1.83 (1.56-2.15) | 1.87E-13 | 1.98 (1.65-2.37) | 1.56E-13 | 4.25 (3.64-4.96) | 3.70E-75 |  |
| Marital status |  |  |  |  |  |  |  |  |
| Married/Living with partner | 1.00 (Reference) | 2.07 (1.74-2.47) | 4.79E-16 | 2.65 (2.12-3.31) | 7.02E-18 | 6.17 (5.11-7.45) | 3.71E-80 | 7.30E-01 |
| Others | 1.00 (Reference) | 1.81 (1.50-2.18) | 7.78E-10 | 1.87 (1.49-2.36) | 9.21E-08 | 3.70 (3.04-4.51) | 8.56E-39 |  |
| Educational status |  |  |  |  |  |  |  |  |
| Less than high school | 1.00 (Reference) | 1.75 (1.33-2.30) | 7.13E-05 | 1.49 (1.11-1.99) | 7.74E-03 | 3.43 (2.64-4.44) | 1.85E-20 | 3.84E-13 |
| High school | 1.00 (Reference) | 1.77 (1.43-2.20) | 1.59E-07 | 2.55 (1.96-3.32) | 3.64E-12 | 4.67 (3.73-5.84) | 1.30E-41 |  |
| College or above | 1.00 (Reference) | 2.39 (1.96-2.91) | 7.44E-18 | 2.91 (2.23-3.80) | 4.38E-15 | 6.18 (4.89-7.82) | 4.87E-52 |  |

| **eTable 7.** Association of frailty assessed by FI-Lab and heart stress with risk of all-cause mortality in subgroups. | | | | | | | | |
| --- | --- | --- | --- | --- | --- | --- | --- | --- |
| **Subgroups** | **No Heart Stress or Frailty** | **Heart Stress, no Frailty** | | **Frailty, no Heart Stress** | | **Frailty, and Heart Stress** | |  |
|  |  | **HR (95% CI)** | **P-value** | **HR (95% CI)** | **P-value** | **HR (95% CI)** | **P-value** | **P-int** |
| **NHANES** |  |  |  |  |  |  |  |  |
| Age group |  |  |  |  |  |  |  |  |
| 20-49 | 1.00 (Reference) | 1.53 (1.12-2.09) | 7.17E-03 | 2.26 (1.59-3.23) | 6.77E-06 | 5.80 (3.92-8.59) | 1.59E-18 | 4.19E-02 |
| 50-64 | 1.00 (Reference) | 2.37 (1.97-2.86) | 1.63E-19 | 2.17 (1.65-2.84) | 2.54E-08 | 6.59 (4.91-8.85) | 3.58E-36 |  |
| 65 years or older | 1.00 (Reference) | 1.78 (1.62-1.94) | 4.27E-36 | 1.72 (1.44-2.06) | 3.29E-09 | 2.96 (2.58-3.40) | 7.75E-54 |  |
| Gender |  |  |  |  |  |  |  |  |
| Male | 1.00 (Reference) | 1.99 (1.78-2.23) | 2.19E-33 | 2.01 (1.70-2.38) | 2.42E-16 | 3.19 (2.73-3.71) | 1.59E-49 | 1.26E-01 |
| Female | 1.00 (Reference) | 1.69 (1.51-1.88) | 2.42E-21 | 1.82 (1.43-2.32) | 1.01E-06 | 4.18 (3.47-5.04) | 9.07E-51 |  |
| Body mass index |  |  |  |  |  |  |  |  |
| Normal or underweight | 1.00 (Reference) | 1.51 (1.31-1.75) | 1.84E-08 | 1.78 (1.36-2.34) | 2.76E-05 | 2.77 (2.23-3.44) | 3.59E-20 | 2.25E-02 |
| Overweight | 1.00 (Reference) | 1.79 (1.57-2.05) | 3.51E-18 | 2.26 (1.81-2.83) | 1.04E-12 | 3.29 (2.65-4.08) | 3.26E-27 |  |
| Obese | 1.00 (Reference) | 1.89 (1.64-2.18) | 2.49E-18 | 1.62 (1.26-2.08) | 1.51E-04 | 3.64 (2.90-4.58) | 1.20E-28 |  |
| Race/Ethnicity |  |  |  |  |  |  |  |  |
| White/Caucasian | 1.00 (Reference) | 1.84 (1.67-2.02) | 1.13E-34 | 1.77 (1.45-2.16) | 2.35E-08 | 3.12 (2.66-3.67) | 1.62E-43 | 7.91E-01 |
| Black/African American | 1.00 (Reference) | 2.14 (1.73-2.63) | 1.30E-12 | 1.65 (1.26-2.18) | 3.39E-04 | 4.13 (3.23-5.27) | 5.31E-30 |  |
| Others | 1.00 (Reference) | 1.67 (1.41-1.98) | 3.47E-09 | 2.20 (1.69-2.87) | 3.72E-09 | 3.40 (2.63-4.39) | 5.45E-21 |  |
| Smoking status |  |  |  |  |  |  |  |  |
| Ever | 1.00 (Reference) | 1.79 (1.62-1.99) | 1.15E-28 | 1.76 (1.48-2.10) | 2.86E-10 | 3.59 (3.09-4.17) | 5.62E-62 | 1.39E-01 |
| Never | 1.00 (Reference) | 1.81 (1.60-2.03) | 3.21E-22 | 2.14 (1.72-2.66) | 7.25E-12 | 3.22 (2.66-3.90) | 8.98E-33 |  |
| Vigorous physical activity |  |  |  |  |  |  |  |  |
| Yes | 1.00 (Reference) | 1.40 (1.11-1.77) | 4.59E-03 | 1.37 (0.91-2.07) | 1.31E-01 | 3.34 (2.19-5.11) | 2.36E-08 | 5.18E-02 |
| No | 1.00 (Reference) | 1.85 (1.70-2.01) | 1.48E-47 | 2.00 (1.73-2.31) | 1.29E-20 | 3.39 (3.00-3.84) | 4.61E-83 |  |
| Marital status |  |  |  |  |  |  |  |  |
| Married/Living with partner | 1.00 (Reference) | 1.84 (1.66-2.05) | 1.76E-29 | 1.91 (1.58-2.32) | 3.47E-11 | 3.08 (2.60-3.66) | 1.34E-37 | 8.74E-01 |
| Others | 1.00 (Reference) | 1.74 (1.55-1.96) | 8.26E-20 | 1.65 (1.34-2.03) | 2.03E-06 | 3.50 (2.95-4.15) | 8.26E-47 |  |
| Educational status |  |  |  |  |  |  |  |  |
| Less than high school | 1.00 (Reference) | 1.75 (1.56-1.97) | 1.62E-20 | 1.82 (1.48-2.23) | 1.20E-08 | 3.13 (2.64-3.71) | 1.33E-39 | 1.85E-03 |
| High school | 1.00 (Reference) | 1.81 (1.54-2.12) | 2.57E-13 | 2.07 (1.55-2.77) | 9.70E-07 | 4.14 (3.20-5.37) | 7.19E-27 |  |
| College or above | 1.00 (Reference) | 1.80 (1.57-2.07) | 4.00E-17 | 1.85 (1.45-2.36) | 5.85E-07 | 3.27 (2.63-4.07) | 2.24E-26 |  |
| **HRS** |  |  |  |  |  |  |  |  |
| Age group |  |  |  |  |  |  |  |  |
| 50-64 | 1.00 (Reference) | 2.42 (1.65-3.54) | 6.37E-06 | 4.55 (3.14-6.59) | 1.33E-15 | 11.52 (8.18-16.22) | 1.64E-44 | 1.26E-06 |
| 65 years or older | 1.00 (Reference) | 1.80 (1.57-2.06) | 3.33E-17 | 2.14 (1.78-2.57) | 6.33E-16 | 4.15 (3.59-4.79) | 7.22E-83 |  |
| Gender |  |  |  |  |  |  |  |  |
| Male | 1.00 (Reference) | 2.23 (1.83-2.71) | 1.10E-15 | 2.46 (1.97-3.07) | 3.23E-15 | 4.92 (4.07-5.95) | 9.96E-61 | 8.28E-01 |
| Female | 1.00 (Reference) | 1.63 (1.37-1.93) | 1.75E-08 | 2.50 (1.95-3.19) | 2.68E-13 | 4.54 (3.74-5.50) | 3.26E-53 |  |
| Body mass index |  |  |  |  |  |  |  |  |
| Normal or underweight | 1.00 (Reference) | 1.81 (1.46-2.25) | 6.38E-08 | 2.62 (1.92-3.57) | 1.42E-09 | 4.72 (3.73-5.98) | 4.43E-38 | 5.14E-01 |
| Overweight | 1.00 (Reference) | 1.66 (1.32-2.08) | 1.33E-05 | 2.25 (1.67-3.02) | 8.07E-08 | 4.50 (3.54-5.71) | 5.10E-35 |  |
| Obese | 1.00 (Reference) | 2.08 (1.65-2.62) | 4.68E-10 | 2.42 (1.86-3.16) | 7.18E-11 | 4.45 (3.52-5.64) | 3.74E-35 |  |
| Race/Ethnicity |  |  |  |  |  |  |  |  |
| White/Caucasian | 1.00 (Reference) | 1.76 (1.53-2.04) | 7.04E-15 | 2.53 (2.07-3.08) | 3.22E-20 | 4.72 (4.05-5.51) | 2.23E-86 | 9.39E-01 |
| Black/African American | 1.00 (Reference) | 2.05 (1.44-2.93) | 7.64E-05 | 2.27 (1.58-3.26) | 8.34E-06 | 4.82 (3.47-6.70) | 6.15E-21 |  |
| Others | 1.00 (Reference) | 2.81 (1.64-4.79) | 1.58E-04 | 2.33 (1.23-4.40) | 9.02E-03 | 4.29 (2.43-7.55) | 4.69E-07 |  |
| Smoking status |  |  |  |  |  |  |  |  |
| Ever | 1.00 (Reference) | 1.72 (1.46-2.02) | 1.33E-10 | 2.00 (1.62-2.47) | 1.09E-10 | 4.09 (3.44-4.85) | 7.81E-58 | 1.04E-01 |
| Never | 1.00 (Reference) | 1.99 (1.61-2.45) | 1.12E-10 | 3.22 (2.47-4.22) | 1.21E-17 | 5.68 (4.56-7.07) | 2.89E-54 |  |
| Vigorous physical activity |  |  |  |  |  |  |  |  |
| Yes | 1.00 (Reference) | 2.08 (1.63-2.64) | 2.14E-09 | 2.62 (1.90-3.62) | 5.28E-09 | 4.51 (3.41-5.96) | 3.62E-26 | 5.87E-01 |
| No | 1.00 (Reference) | 1.75 (1.50-2.03) | 8.88E-13 | 2.24 (1.85-2.71) | 2.66E-16 | 4.27 (3.66-4.98) | 1.13E-75 |  |
| Marital status |  |  |  |  |  |  |  |  |
| Married/Living with partner | 1.00 (Reference) | 2.06 (1.72-2.46) | 5.35E-15 | 2.74 (2.20-3.42) | 3.02E-19 | 5.44 (4.50-6.57) | 5.07E-69 | 9.27E-01 |
| Others | 1.00 (Reference) | 1.63 (1.36-1.95) | 1.51E-07 | 2.03 (1.57-2.61) | 4.58E-08 | 3.90 (3.22-4.73) | 7.85E-44 |  |
| Educational status |  |  |  |  |  |  |  |  |
| Less than high school | 1.00 (Reference) | 1.79 (1.37-2.32) | 1.53E-05 | 2.14 (1.57-2.90) | 1.17E-06 | 3.84 (2.96-4.97) | 3.00E-24 | 4.71E-03 |
| High school | 1.00 (Reference) | 1.68 (1.36-2.09) | 2.01E-06 | 2.37 (1.80-3.13) | 9.30E-10 | 4.28 (3.43-5.35) | 2.37E-37 |  |
| College or above | 1.00 (Reference) | 2.08 (1.70-2.54) | 1.48E-12 | 2.64 (2.00-3.48) | 6.23E-12 | 6.16 (4.93-7.71) | 4.05E-57 |  |

| **eTable 8.** Association of frailty assessed by FI-Combined and heart stress with risk of all-cause mortality in subgroups. | | | | | | | | |
| --- | --- | --- | --- | --- | --- | --- | --- | --- |
| **Subgroups** | **No Heart Stress or Frailty** | **Heart Stress, no Frailty** | | **Frailty, no Heart Stress** | | **Frailty, and Heart Stress** | |  |
|  |  | **HR (95% CI)** | **P-value** | **HR (95% CI)** | **P-value** | **HR (95% CI)** | **P-value** | **P-int** |
| **NHANES** |  |  |  |  |  |  |  |  |
| Age group |  |  |  |  |  |  |  |  |
| 20-49 | 1.00 (Reference) | 2.86 (1.76-4.64) | 2.30E-05 | 4.32 (2.64-7.08) | 6.38E-09 | 9.03 (4.85-16.80) | 3.70E-12 | 1.09E-03 |
| 50-64 | 1.00 (Reference) | 2.12 (1.70-2.63) | 1.52E-11 | 2.48 (1.88-3.28) | 1.40E-10 | 6.47 (4.80-8.71) | 9.79E-35 |  |
| 65 years or older | 1.00 (Reference) | 1.77 (1.61-1.94) | 1.61E-33 | 2.32 (1.97-2.74) | 3.82E-24 | 3.58 (3.14-4.08) | 1.50E-80 |  |
| Gender |  |  |  |  |  |  |  |  |
| Male | 1.00 (Reference) | 1.90 (1.69-2.14) | 1.22E-27 | 2.67 (2.23-3.19) | 8.78E-27 | 4.30 (3.65-5.07) | 1.33E-68 | 4.25E-01 |
| Female | 1.00 (Reference) | 1.75 (1.56-1.98) | 6.81E-20 | 2.58 (2.12-3.14) | 1.98E-21 | 3.80 (3.21-4.49) | 8.64E-55 |  |
| Body mass index |  |  |  |  |  |  |  |  |
| Normal or underweight | 1.00 (Reference) | 1.54 (1.33-1.80) | 1.75E-08 | 3.42 (2.61-4.47) | 4.32E-19 | 3.80 (3.03-4.77) | 1.60E-30 | 1.52E-01 |
| Overweight | 1.00 (Reference) | 1.83 (1.60-2.11) | 7.34E-18 | 2.68 (2.09-3.44) | 9.97E-15 | 3.51 (2.81-4.39) | 1.73E-28 |  |
| Obese | 1.00 (Reference) | 1.97 (1.68-2.31) | 3.54E-17 | 1.87 (1.49-2.33) | 4.01E-08 | 3.92 (3.17-4.85) | 2.99E-36 |  |
| Race/Ethnicity |  |  |  |  |  |  |  |  |
| White/Caucasian | 1.00 (Reference) | 1.78 (1.60-1.97) | 7.09E-28 | 2.43 (2.01-2.93) | 3.58E-20 | 4.72 (4.03-5.53) | 3.88E-82 | 1.32E-02 |
| Black/African American | 1.00 (Reference) | 2.19 (1.76-2.74) | 3.53E-12 | 2.80 (2.09-3.75) | 6.11E-12 | 4.10 (3.15-5.33) | 5.78E-26 |  |
| Others | 1.00 (Reference) | 1.82 (1.51-2.19) | 4.87E-10 | 2.49 (1.94-3.20) | 1.07E-12 | 3.02 (2.37-3.86) | 5.41E-19 |  |
| Smoking status |  |  |  |  |  |  |  |  |
| Ever | 1.00 (Reference) | 1.85 (1.65-2.06) | 1.35E-27 | 2.67 (2.25-3.16) | 9.11E-30 | 4.23 (3.62-4.93) | 3.45E-75 | 1.45E-01 |
| Never | 1.00 (Reference) | 1.76 (1.55-2.00) | 5.60E-18 | 2.37 (1.91-2.94) | 3.89E-15 | 4.02 (3.36-4.81) | 4.00E-52 |  |
| Vigorous physical activity |  |  |  |  |  |  |  |  |
| Yes | 1.00 (Reference) | 1.41 (1.10-1.80) | 6.00E-03 | 2.02 (1.03-3.97) | 4.09E-02 | 9.13 (4.82-17.30) | 1.19E-11 | 8.19E-02 |
| No | 1.00 (Reference) | 1.87 (1.71-2.05) | 7.09E-43 | 2.61 (2.28-2.99) | 1.50E-43 | 4.03 (3.58-4.55) | 9.61E-115 |  |
| Marital status |  |  |  |  |  |  |  |  |
| Married/Living with partner | 1.00 (Reference) | 1.83 (1.64-2.05) | 5.49E-26 | 2.37 (1.96-2.88) | 2.59E-18 | 4.31 (3.62-5.13) | 2.57E-60 | 6.33E-01 |
| Others | 1.00 (Reference) | 1.74 (1.53-1.98) | 5.46E-17 | 2.50 (2.06-3.03) | 1.52E-20 | 3.78 (3.21-4.46) | 2.28E-56 |  |
| Educational status |  |  |  |  |  |  |  |  |
| Less than high school | 1.00 (Reference) | 1.83 (1.60-2.08) | 1.04E-19 | 2.22 (1.85-2.66) | 8.08E-18 | 3.42 (2.91-4.03) | 2.25E-49 | 1.60E-02 |
| High school | 1.00 (Reference) | 1.81 (1.53-2.14) | 4.58E-12 | 2.82 (2.08-3.84) | 3.48E-11 | 5.39 (4.17-6.96) | 3.91E-38 |  |
| College or above | 1.00 (Reference) | 1.77 (1.53-2.04) | 1.79E-14 | 2.95 (2.27-3.83) | 6.41E-16 | 4.76 (3.76-6.01) | 4.67E-39 |  |
| **HRS** |  |  |  |  |  |  |  |  |
| Age group |  |  |  |  |  |  |  |  |
| 50-64 | 1.00 (Reference) | 2.55 (1.74-3.72) | 1.26E-06 | 3.74 (2.55-5.48) | 1.45E-11 | 10.83 (7.65-15.34) | 4.30E-41 | 4.64E-04 |
| 65 years or older | 1.00 (Reference) | 1.78 (1.55-2.04) | 4.84E-16 | 2.44 (2.04-2.92) | 3.21E-22 | 4.90 (4.24-5.66) | 8.16E-102 |  |
| Gender |  |  |  |  |  |  |  |  |
| Male | 1.00 (Reference) | 2.15 (1.79-2.58) | 3.37E-16 | 2.60 (2.05-3.30) | 5.01E-15 | 5.94 (4.89-7.21) | 6.36E-72 | 5.07E-01 |
| Female | 1.00 (Reference) | 1.57 (1.30-1.89) | 1.95E-06 | 2.73 (2.18-3.41) | 1.11E-18 | 4.93 (4.09-5.95) | 1.65E-62 |  |
| Body mass index |  |  |  |  |  |  |  |  |
| Normal or underweight | 1.00 (Reference) | 1.90 (1.53-2.35) | 3.72E-09 | 3.21 (2.37-4.36) | 5.56E-14 | 6.20 (4.88-7.87) | 2.58E-50 | 2.29E-02 |
| Overweight | 1.00 (Reference) | 1.55 (1.24-1.95) | 1.57E-04 | 2.03 (1.50-2.76) | 5.59E-06 | 4.67 (3.69-5.92) | 1.77E-37 |  |
| Obese | 1.00 (Reference) | 2.14 (1.67-2.75) | 1.82E-09 | 2.63 (2.04-3.41) | 1.57E-13 | 4.79 (3.77-6.10) | 2.84E-37 |  |
| Race/Ethnicity |  |  |  |  |  |  |  |  |
| White/Caucasian | 1.00 (Reference) | 1.80 (1.56-2.09) | 1.71E-15 | 3.03 (2.51-3.67) | 1.26E-30 | 5.44 (4.65-6.36) | 3.24E-99 | 9.32E-01 |
| Black/African American | 1.00 (Reference) | 1.80 (1.25-2.59) | 1.49E-03 | 1.93 (1.35-2.77) | 3.17E-04 | 5.37 (3.91-7.36) | 1.86E-25 |  |
| Others | 1.00 (Reference) | 2.50 (1.45-4.32) | 1.00E-03 | 1.36 (0.67-2.78) | 3.96E-01 | 4.52 (2.55-7.98) | 2.16E-07 |  |
| Smoking status |  |  |  |  |  |  |  |  |
| Ever | 1.00 (Reference) | 1.73 (1.46-2.05) | 2.44E-10 | 2.50 (2.04-3.06) | 8.12E-19 | 5.25 (4.43-6.22) | 3.98E-81 | 2.60E-01 |
| Never | 1.00 (Reference) | 2.04 (1.66-2.51) | 1.92E-11 | 2.71 (2.06-3.56) | 1.22E-12 | 5.29 (4.22-6.62) | 2.19E-47 |  |
| Vigorous physical activity |  |  |  |  |  |  |  |  |
| Yes | 1.00 (Reference) | 2.20 (1.77-2.74) | 1.60E-12 | 2.91 (2.00-4.24) | 2.41E-08 | 4.88 (3.55-6.70) | 1.14E-22 | 9.48E-01 |
| No | 1.00 (Reference) | 1.67 (1.41-1.96) | 1.04E-09 | 2.27 (1.89-2.73) | 2.52E-18 | 4.62 (3.97-5.37) | 1.23E-86 |  |
| Marital status |  |  |  |  |  |  |  |  |
| Married/Living with partner | 1.00 (Reference) | 1.87 (1.56-2.24) | 6.73E-12 | 2.74 (2.18-3.44) | 7.55E-18 | 6.67 (5.54-8.01) | 2.12E-90 | 6.28E-01 |
| Others | 1.00 (Reference) | 1.74 (1.43-2.10) | 1.86E-08 | 2.42 (1.92-3.05) | 8.79E-14 | 4.14 (3.40-5.04) | 8.20E-46 |  |
| Educational status |  |  |  |  |  |  |  |  |
| Less than high school | 1.00 (Reference) | 1.62 (1.22-2.14) | 8.33E-04 | 1.79 (1.33-2.41) | 1.17E-04 | 3.65 (2.82-4.73) | 1.08E-22 | 4.98E-12 |
| High school | 1.00 (Reference) | 1.65 (1.33-2.06) | 7.88E-06 | 3.05 (2.33-3.98) | 3.23E-16 | 5.16 (4.14-6.43) | 1.89E-48 |  |
| College or above | 1.00 (Reference) | 2.18 (1.79-2.66) | 1.74E-14 | 3.08 (2.34-4.05) | 1.02E-15 | 6.96 (5.54-8.76) | 5.07E-62 |  |

| **eTable 9.** Association of frailty and heart stress with risk of CVD mortality in subgroups in NHANES. | | | | | | | | |
| --- | --- | --- | --- | --- | --- | --- | --- | --- |
| **A.** FI-Self-report | | | | | | | | |
| **Subgroups** | **No Heart Stress or Frailty** | **Heart Stress, no Frailty** | | **Frailty, no Heart Stress** | | **Frailty, and Heart Stress** | |  |
|  |  | **HR (95% CI)** | **P-value** | **HR (95% CI)** | **P-value** | **HR (95% CI)** | **P-value** | **P-int** |
| Age group |  |  |  |  |  |  |  |  |
| 20-64 | 1.00 (Reference) | 4.58 (3.20-6.55) | 7.77E-17 | 2.40 (1.56-3.69) | 7.03E-05 | 11.59 (7.15-18.79) | 2.70E-23 | 6.65E-01 |
| 65 years or older | 1.00 (Reference) | 2.39 (2.02-2.82) | 9.33E-25 | 2.47 (1.94-3.14) | 1.42E-13 | 4.71 (3.77-5.89) | 2.84E-42 |  |
| Gender |  |  |  |  |  |  |  |  |
| Male | 1.00 (Reference) | 2.81 (2.31-3.43) | 2.14E-24 | 2.34 (1.76-3.10) | 3.86E-09 | 6.92 (5.25-9.12) | 6.09E-43 | 1.86E-01 |
| Female | 1.00 (Reference) | 2.59 (2.05-3.28) | 2.08E-15 | 2.78 (2.03-3.80) | 1.88E-10 | 4.56 (3.41-6.10) | 1.44E-24 |  |
| Body mass index |  |  |  |  |  |  |  |  |
| Normal or underweight | 1.00 (Reference) | 2.37 (1.78-3.15) | 3.16E-09 | 3.68 (2.35-5.78) | 1.44E-08 | 3.99 (2.63-6.07) | 8.90E-11 | 1.42E-01 |
| Overweight | 1.00 (Reference) | 2.78 (2.17-3.56) | 7.71E-16 | 2.64 (1.85-3.76) | 9.78E-08 | 5.82 (3.98-8.52) | 1.13E-19 |  |
| Obese | 1.00 (Reference) | 2.57 (1.94-3.40) | 4.68E-11 | 1.89 (1.33-2.69) | 3.73E-04 | 5.55 (3.93-7.84) | 2.75E-22 |  |
| Race/Ethnicity |  |  |  |  |  |  |  |  |
| White/Caucasian | 1.00 (Reference) | 2.44 (2.02-2.94) | 8.11E-21 | 2.27 (1.70-3.02) | 2.01E-08 | 6.50 (4.93-8.56) | 3.97E-40 | 4.59E-02 |
| Black/African American | 1.00 (Reference) | 4.16 (2.84-6.09) | 2.50E-13 | 2.21 (1.33-3.66) | 2.24E-03 | 5.72 (3.67-8.93) | 1.43E-14 |  |
| Others | 1.00 (Reference) | 2.84 (2.00-4.02) | 4.73E-09 | 2.96 (1.97-4.45) | 1.85E-07 | 3.90 (2.53-6.03) | 8.54E-10 |  |
| Smoking status |  |  |  |  |  |  |  |  |
| Ever | 1.00 (Reference) | 2.74 (2.24-3.37) | 4.25E-22 | 2.39 (1.83-3.13) | 2.53E-10 | 5.77 (4.41-7.55) | 2.32E-37 | 3.04E-01 |
| Never | 1.00 (Reference) | 2.61 (2.08-3.28) | 9.49E-17 | 2.89 (2.07-4.03) | 4.37E-10 | 5.83 (4.33-7.83) | 2.09E-31 |  |
| Vigorous physical activity |  |  |  |  |  |  |  |  |
| Yes | 1.00 (Reference) | 2.34 (1.51-3.62) | 1.44E-04 | 1.19 (0.50-2.85) | 6.96E-01 | 7.14 (2.36-21.58) | 4.97E-04 | 2.49E-02 |
| No | 1.00 (Reference) | 2.70 (2.29-3.17) | 2.61E-33 | 2.72 (2.20-3.38) | 7.31E-20 | 5.68 (4.63-6.97) | 1.17E-62 |  |
| Marital status |  |  |  |  |  |  |  |  |
| Married/Living with partner | 1.00 (Reference) | 2.57 (2.11-3.14) | 1.24E-20 | 2.39 (1.78-3.23) | 9.53E-09 | 5.10 (3.77-6.91) | 6.70E-26 | 9.10E-01 |
| Others | 1.00 (Reference) | 2.64 (2.07-3.37) | 5.29E-15 | 2.42 (1.75-3.35) | 8.74E-08 | 5.66 (4.27-7.52) | 3.84E-33 |  |
| Educational status |  |  |  |  |  |  |  |  |
| Less than high school | 1.00 (Reference) | 2.87 (2.26-3.65) | 5.76E-18 | 2.64 (1.99-3.49) | 1.20E-11 | 4.92 (3.74-6.49) | 1.12E-29 | 2.05E-02 |
| High school | 1.00 (Reference) | 2.50 (1.85-3.40) | 3.56E-09 | 2.12 (1.27-3.54) | 4.18E-03 | 7.24 (4.59-11.42) | 1.71E-17 |  |
| College or above | 1.00 (Reference) | 2.44 (1.89-3.16) | 1.23E-11 | 2.39 (1.55-3.68) | 7.40E-05 | 5.42 (3.62-8.12) | 2.14E-16 |  |
| **B.** FI-Lab | | | | | | | | |
| **Subgroups** | **No Heart Stress or Frailty** | **Heart Stress, no Frailty** | | **Frailty, no Heart Stress** | | **Frailty, and Heart Stress** | |  |
|  |  | **HR (95% CI)** | **P-value** | **HR (95% CI)** | **P-value** | **HR (95% CI)** | **P-value** | **P-int** |
| Age group |  |  |  |  |  |  |  |  |
| 20-64 | 1.00 (Reference) | 3.43 (2.58-4.55) | 1.32E-17 | 1.51 (0.90-2.53) | 1.23E-01 | 9.92 (6.57-14.97) | 8.72E-28 | 9.09E-01 |
| 65 years or older | 1.00 (Reference) | 2.28 (1.96-2.66) | 1.88E-26 | 1.84 (1.30-2.60) | 6.28E-04 | 3.41 (2.70-4.32) | 1.42E-24 |  |
| Gender |  |  |  |  |  |  |  |  |
| Male | 1.00 (Reference) | 2.87 (2.39-3.46) | 7.84E-29 | 2.05 (1.44-2.92) | 6.22E-05 | 4.14 (3.19-5.38) | 2.09E-26 | 3.37E-02 |
| Female | 1.00 (Reference) | 2.22 (1.82-2.69) | 9.76E-16 | 1.79 (1.08-2.97) | 2.32E-02 | 5.55 (3.97-7.75) | 9.38E-24 |  |
| Body mass index |  |  |  |  |  |  |  |  |
| Normal or underweight | 1.00 (Reference) | 2.08 (1.60-2.70) | 4.63E-08 | 2.26 (1.26-4.06) | 6.17E-03 | 2.41 (1.57-3.72) | 6.25E-05 | 1.17E-01 |
| Overweight | 1.00 (Reference) | 2.44 (1.94-3.06) | 1.71E-14 | 2.45 (1.52-3.94) | 2.33E-04 | 5.08 (3.61-7.16) | 1.37E-20 |  |
| Obese | 1.00 (Reference) | 2.55 (2.01-3.23) | 1.29E-14 | 1.42 (0.86-2.35) | 1.75E-01 | 5.30 (3.55-7.89) | 2.71E-16 |  |
| Race/Ethnicity |  |  |  |  |  |  |  |  |
| White/Caucasian | 1.00 (Reference) | 2.43 (2.06-2.88) | 2.72E-25 | 2.09 (1.39-3.15) | 4.24E-04 | 3.60 (2.73-4.75) | 1.21E-19 | 5.10E-01 |
| Black/African American | 1.00 (Reference) | 3.45 (2.42-4.92) | 7.63E-12 | 1.84 (1.10-3.09) | 2.03E-02 | 7.55 (4.92-11.58) | 2.09E-20 |  |
| Others | 1.00 (Reference) | 2.32 (1.74-3.11) | 1.43E-08 | 1.38 (0.67-2.83) | 3.82E-01 | 3.49 (2.18-5.58) | 1.89E-07 |  |
| Smoking status |  |  |  |  |  |  |  |  |
| Ever | 1.00 (Reference) | 2.52 (2.10-3.02) | 2.14E-23 | 2.09 (1.44-3.01) | 9.02E-05 | 4.86 (3.73-6.34) | 1.29E-31 | 6.62E-01 |
| Never | 1.00 (Reference) | 2.45 (2.00-2.99) | 2.05E-18 | 1.72 (1.08-2.74) | 2.21E-02 | 3.89 (2.80-5.41) | 6.54E-16 |  |
| Vigorous physical activity |  |  |  |  |  |  |  |  |
| Yes | 1.00 (Reference) | 2.23 (1.49-3.33) | 9.99E-05 | 1.43 (0.65-3.13) | 3.71E-01 | 5.21 (2.09-12.95) | 3.90E-04 | 8.03E-01 |
| No | 1.00 (Reference) | 2.51 (2.18-2.90) | 9.70E-37 | 1.93 (1.42-2.63) | 2.91E-05 | 4.23 (3.42-5.23) | 2.15E-40 |  |
| Marital status |  |  |  |  |  |  |  |  |
| Married/Living with partner | 1.00 (Reference) | 2.45 (2.04-2.95) | 1.41E-21 | 1.80 (1.20-2.70) | 4.24E-03 | 3.52 (2.61-4.76) | 2.81E-16 | 3.55E-01 |
| Others | 1.00 (Reference) | 2.45 (2.00-3.01) | 8.38E-18 | 1.73 (1.13-2.64) | 1.23E-02 | 4.69 (3.50-6.30) | 6.93E-25 |  |
| Educational status |  |  |  |  |  |  |  |  |
| Less than high school | 1.00 (Reference) | 2.51 (2.06-3.07) | 8.51E-20 | 2.30 (1.50-3.51) | 1.23E-04 | 4.03 (3.03-5.36) | 1.13E-21 | 4.15E-02 |
| High school | 1.00 (Reference) | 2.37 (1.79-3.15) | 1.86E-09 | 1.86 (0.97-3.57) | 6.26E-02 | 5.56 (3.40-9.07) | 7.01E-12 |  |
| College or above | 1.00 (Reference) | 2.44 (1.92-3.10) | 3.36E-13 | 1.56 (0.94-2.60) | 8.56E-02 | 4.08 (2.76-6.02) | 1.55E-12 |  |
| **C.** FI-Combined | | | | | | | | |
| **Subgroups** | **No Heart Stress or Frailty** | **Heart Stress, no Frailty** | | **Frailty, no Heart Stress** | | **Frailty, and Heart Stress** | |  |
|  |  | **HR (95% CI)** | **P-value** | **HR (95% CI)** | **P-value** | **HR (95% CI)** | **P-value** | **P-int** |
| Age group |  |  |  |  |  |  |  |  |
| 20-64 | 1.00 (Reference) | 3.96 (2.83-5.54) | 1.09E-15 | 2.04 (1.19-3.49) | 9.41E-03 | 13.93 (8.72-22.24) | 3.04E-28 | 6.53E-01 |
| 65 years or older | 1.00 (Reference) | 2.28 (1.94-2.67) | 3.58E-24 | 2.86 (2.14-3.83) | 1.78E-12 | 4.89 (3.90-6.13) | 4.16E-43 |  |
| Gender |  |  |  |  |  |  |  |  |
| Male | 1.00 (Reference) | 2.67 (2.20-3.24) | 1.29E-23 | 2.77 (1.96-3.90) | 6.07E-09 | 6.78 (5.13-8.97) | 2.58E-41 | 8.26E-01 |
| Female | 1.00 (Reference) | 2.40 (1.93-2.99) | 4.74E-15 | 2.86 (1.97-4.16) | 3.79E-08 | 5.24 (3.88-7.08) | 3.71E-27 |  |
| Body mass index |  |  |  |  |  |  |  |  |
| Normal or underweight | 1.00 (Reference) | 2.02 (1.54-2.66) | 4.43E-07 | 4.54 (2.55-8.09) | 2.91E-07 | 4.61 (3.04-7.00) | 7.79E-13 | 4.21E-01 |
| Overweight | 1.00 (Reference) | 2.57 (2.02-3.26) | 1.35E-14 | 3.38 (2.13-5.35) | 2.24E-07 | 5.53 (3.77-8.11) | 2.18E-18 |  |
| Obese | 1.00 (Reference) | 2.65 (2.04-3.43) | 1.83E-13 | 1.88 (1.24-2.84) | 2.72E-03 | 5.55 (3.86-7.98) | 2.03E-20 |  |
| Race/Ethnicity |  |  |  |  |  |  |  |  |
| White/Caucasian | 1.00 (Reference) | 2.38 (1.99-2.85) | 2.13E-21 | 2.42 (1.72-3.38) | 2.89E-07 | 7.49 (5.61-10.00) | 1.44E-42 | 1.24E-02 |
| Black/African American | 1.00 (Reference) | 3.56 (2.46-5.13) | 1.15E-11 | 2.78 (1.53-5.04) | 7.59E-04 | 6.79 (4.38-10.54) | 1.36E-17 |  |
| Others | 1.00 (Reference) | 2.46 (1.78-3.40) | 5.14E-08 | 3.38 (1.95-5.87) | 1.44E-05 | 3.71 (2.43-5.67) | 1.40E-09 |  |
| Smoking status |  |  |  |  |  |  |  |  |
| Ever | 1.00 (Reference) | 2.60 (2.14-3.16) | 3.93E-22 | 3.15 (2.27-4.38) | 6.43E-12 | 6.25 (4.74-8.26) | 2.73E-38 | 2.46E-01 |
| Never | 1.00 (Reference) | 2.39 (1.92-2.97) | 3.37E-15 | 2.64 (1.76-3.95) | 2.48E-06 | 6.32 (4.69-8.51) | 6.63E-34 |  |
| Vigorous physical activity |  |  |  |  |  |  |  |  |
| Yes | 1.00 (Reference) | 2.31 (1.51-3.53) | 1.03E-04 | 0.54 (0.07-4.17) | 5.57E-01 | 14.99 (4.26-52.73) | 2.45E-05 | 7.69E-02 |
| No | 1.00 (Reference) | 2.53 (2.17-2.95) | 1.90E-32 | 3.07 (2.38-3.96) | 8.82E-18 | 6.00 (4.88-7.37) | 6.13E-65 |  |
| Marital status |  |  |  |  |  |  |  |  |
| Married/Living with partner | 1.00 (Reference) | 2.45 (2.02-2.97) | 1.02E-19 | 2.49 (1.71-3.64) | 2.30E-06 | 5.94 (4.37-8.09) | 1.09E-29 | 2.89E-01 |
| Others | 1.00 (Reference) | 2.48 (1.99-3.11) | 1.61E-15 | 2.92 (2.01-4.24) | 1.99E-08 | 5.88 (4.45-7.79) | 2.83E-35 |  |
| Educational status |  |  |  |  |  |  |  |  |
| Less than high school | 1.00 (Reference) | 2.68 (2.15-3.34) | 1.99E-18 | 3.15 (2.28-4.36) | 4.54E-12 | 5.25 (3.98-6.93) | 1.18E-31 | 1.65E-02 |
| High school | 1.00 (Reference) | 2.26 (1.68-3.05) | 8.50E-08 | 2.48 (1.29-4.74) | 6.26E-03 | 9.92 (6.24-15.78) | 3.17E-22 |  |
| College or above | 1.00 (Reference) | 2.39 (1.86-3.08) | 1.22E-11 | 2.01 (1.12-3.58) | 1.87E-02 | 5.67 (3.79-8.49) | 3.20E-17 |  |

| **eTable 10.** Association of frailty and heart stress with risk of cancer mortality in subgroups in NHANES. | | | | | | | | |
| --- | --- | --- | --- | --- | --- | --- | --- | --- |
| **A.** FI-Self-report | | | | | | | | |
| **Subgroups** | **No Heart Stress or Frailty** | **Heart Stress, no Frailty** | | **Frailty, no Heart Stress** | | **Frailty, and Heart Stress** | |  |
|  |  | **HR (95% CI)** | **P-value** | **HR (95% CI)** | **P-value** | **HR (95% CI)** | **P-value** | **P-int** |
| Age group |  |  |  |  |  |  |  |  |
| 20-64 | 1.00 (Reference) | 2.05 (1.36-3.09) | 5.76E-04 | 2.65 (1.77-3.96) | 2.20E-06 | 5.44 (3.13-9.45) | 1.84E-09 | 7.34E-01 |
| 65 years or older | 1.00 (Reference) | 1.77 (1.41-2.21) | 6.06E-07 | 1.59 (1.16-2.18) | 3.96E-03 | 2.89 (2.02-4.14) | 6.38E-09 |  |
| Gender |  |  |  |  |  |  |  |  |
| Male | 1.00 (Reference) | 2.09 (1.63-2.69) | 6.79E-09 | 2.08 (1.49-2.92) | 1.80E-05 | 3.46 (2.16-5.53) | 2.38E-07 | 7.13E-01 |
| Female | 1.00 (Reference) | 1.46 (1.07-2.01) | 1.88E-02 | 2.10 (1.46-3.01) | 6.08E-05 | 3.70 (2.53-5.41) | 1.40E-11 |  |
| Body mass index |  |  |  |  |  |  |  |  |
| Normal or underweight | 1.00 (Reference) | 1.79 (1.24-2.59) | 1.79E-03 | 2.05 (1.22-3.44) | 6.65E-03 | 5.19 (3.08-8.75) | 6.54E-10 | 4.27E-01 |
| Overweight | 1.00 (Reference) | 1.54 (1.12-2.13) | 8.64E-03 | 1.58 (1.03-2.42) | 3.69E-02 | 2.35 (1.34-4.13) | 2.86E-03 |  |
| Obese | 1.00 (Reference) | 2.40 (1.66-3.46) | 2.85E-06 | 1.90 (1.26-2.88) | 2.20E-03 | 3.63 (2.12-6.22) | 2.50E-06 |  |
| Race/Ethnicity |  |  |  |  |  |  |  |  |
| White/Caucasian | 1.00 (Reference) | 1.84 (1.44-2.35) | 1.31E-06 | 2.25 (1.60-3.15) | 3.02E-06 | 6.43 (4.42-9.35) | 1.86E-22 | 3.40E-04 |
| Black/African American | 1.00 (Reference) | 2.43 (1.48-4.02) | 4.91E-04 | 2.14 (1.24-3.72) | 6.61E-03 | 3.38 (1.53-7.49) | 2.66E-03 |  |
| Others | 1.00 (Reference) | 1.62 (1.04-2.50) | 3.10E-02 | 1.78 (1.11-2.85) | 1.70E-02 | 2.05 (1.13-3.72) | 1.83E-02 |  |
| Smoking status |  |  |  |  |  |  |  |  |
| Ever | 1.00 (Reference) | 1.75 (1.38-2.23) | 5.14E-06 | 1.78 (1.33-2.39) | 1.06E-04 | 3.28 (2.26-4.76) | 3.73E-10 | 1.90E-09 |
| Never | 1.00 (Reference) | 2.01 (1.43-2.82) | 5.26E-05 | 2.46 (1.56-3.87) | 1.00E-04 | 4.72 (2.95-7.55) | 1.04E-10 |  |
| Vigorous physical activity |  |  |  |  |  |  |  |  |
| Yes | 1.00 (Reference) | 1.23 (0.73-2.07) | 4.31E-01 | 1.19 (0.47-3.05) | 7.15E-01 | 0.00 (0.00-Inf) | 9.97E-01 | 2.40E-02 |
| No | 1.00 (Reference) | 1.96 (1.59-2.43) | 6.00E-10 | 2.05 (1.58-2.64) | 4.20E-08 | 3.83 (2.85-5.14) | 4.68E-19 |  |
| Marital status |  |  |  |  |  |  |  |  |
| Married/Living with partner | 1.00 (Reference) | 1.88 (1.46-2.41) | 1.05E-06 | 1.85 (1.34-2.57) | 2.06E-04 | 3.13 (1.98-4.92) | 9.09E-07 | 2.25E-01 |
| Others | 1.00 (Reference) | 1.58 (1.13-2.20) | 7.34E-03 | 2.07 (1.39-3.08) | 3.58E-04 | 4.00 (2.66-6.02) | 2.85E-11 |  |
| Educational status |  |  |  |  |  |  |  |  |
| Less than high school | 1.00 (Reference) | 1.79 (1.30-2.46) | 3.31E-04 | 1.65 (1.15-2.37) | 6.00E-03 | 3.36 (2.22-5.09) | 8.93E-09 | 1.70E-01 |
| High school | 1.00 (Reference) | 1.99 (1.33-2.97) | 8.39E-04 | 2.02 (1.22-3.35) | 6.23E-03 | 5.70 (3.22-10.09) | 2.40E-09 |  |
| College or above | 1.00 (Reference) | 1.80 (1.30-2.49) | 3.94E-04 | 2.46 (1.56-3.88) | 1.01E-04 | 2.87 (1.54-5.36) | 9.06E-04 |  |
| **B.** FI-Lab | | | | | | | | |
| **Subgroups** | **No Heart Stress or Frailty** | **Heart Stress, no Frailty** | | **Frailty, no Heart Stress** | | **Frailty, and Heart Stress** | |  |
|  |  | **HR (95% CI)** | **P-value** | **HR (95% CI)** | **P-value** | **HR (95% CI)** | **P-value** | **P-int** |
| Age group |  |  |  |  |  |  |  |  |
| 20-64 | 1.00 (Reference) | 1.63 (1.20-2.23) | 2.01E-03 | 1.67 (1.06-2.62) | 2.67E-02 | 4.47 (2.64-7.57) | 2.40E-08 | 2.23E-02 |
| 65 years or older | 1.00 (Reference) | 1.80 (1.45-2.23) | 1.09E-07 | 3.07 (2.14-4.40) | 1.01E-09 | 3.22 (2.18-4.75) | 4.08E-09 |  |
| Gender |  |  |  |  |  |  |  |  |
| Male | 1.00 (Reference) | 1.98 (1.55-2.53) | 5.54E-08 | 2.61 (1.88-3.61) | 7.53E-09 | 3.43 (2.39-4.92) | 2.05E-11 | 7.90E-01 |
| Female | 1.00 (Reference) | 1.61 (1.26-2.07) | 1.70E-04 | 1.89 (1.09-3.29) | 2.45E-02 | 5.61 (3.06-10.27) | 2.41E-08 |  |
| Body mass index |  |  |  |  |  |  |  |  |
| Normal or underweight | 1.00 (Reference) | 1.63 (1.15-2.30) | 5.52E-03 | 2.19 (1.22-3.93) | 8.31E-03 | 3.02 (1.87-4.87) | 6.09E-06 | 4.88E-02 |
| Overweight | 1.00 (Reference) | 1.49 (1.11-2.02) | 8.69E-03 | 3.31 (2.18-5.04) | 2.06E-08 | 2.79 (1.45-5.36) | 2.13E-03 |  |
| Obese | 1.00 (Reference) | 2.13 (1.57-2.88) | 9.50E-07 | 1.74 (1.06-2.86) | 2.78E-02 | 5.96 (3.09-11.52) | 1.08E-07 |  |
| Race/Ethnicity |  |  |  |  |  |  |  |  |
| White/Caucasian | 1.00 (Reference) | 1.90 (1.53-2.37) | 9.83E-09 | 2.46 (1.66-3.66) | 7.61E-06 | 4.12 (2.68-6.32) | 9.61E-11 | 3.41E-02 |
| Black/African American | 1.00 (Reference) | 2.37 (1.50-3.75) | 2.24E-04 | 1.52 (0.80-2.88) | 1.97E-01 | 4.23 (2.33-7.69) | 2.23E-06 |  |
| Others | 1.00 (Reference) | 1.37 (0.93-2.03) | 1.15E-01 | 2.90 (1.75-4.80) | 3.68E-05 | 2.53 (1.33-4.83) | 4.76E-03 |  |
| Smoking status |  |  |  |  |  |  |  |  |
| Ever | 1.00 (Reference) | 1.65 (1.32-2.04) | 6.99E-06 | 2.16 (1.55-3.02) | 6.67E-06 | 3.28 (2.22-4.84) | 2.27E-09 | 2.88E-04 |
| Never | 1.00 (Reference) | 1.87 (1.39-2.53) | 4.33E-05 | 2.67 (1.63-4.37) | 1.01E-04 | 5.59 (3.41-9.17) | 9.71E-12 |  |
| Vigorous physical activity |  |  |  |  |  |  |  |  |
| Yes | 1.00 (Reference) | 1.20 (0.74-1.96) | 4.55E-01 | 0.97 (0.42-2.25) | 9.44E-01 | 6.21 (2.43-15.86) | 1.34E-04 | 8.60E-01 |
| No | 1.00 (Reference) | 1.79 (1.48-2.17) | 2.01E-09 | 2.70 (2.00-3.63) | 6.54E-11 | 3.64 (2.63-5.04) | 7.65E-15 |  |
| Marital status |  |  |  |  |  |  |  |  |
| Married/Living with partner | 1.00 (Reference) | 1.72 (1.36-2.16) | 4.36E-06 | 2.33 (1.61-3.37) | 7.43E-06 | 2.61 (1.63-4.18) | 6.36E-05 | 5.64E-01 |
| Others | 1.00 (Reference) | 1.76 (1.33-2.35) | 9.88E-05 | 1.96 (1.25-3.05) | 3.16E-03 | 4.69 (3.02-7.27) | 5.28E-12 |  |
| Educational status |  |  |  |  |  |  |  |  |
| Less than high school | 1.00 (Reference) | 1.70 (1.29-2.25) | 1.74E-04 | 2.77 (1.82-4.22) | 2.25E-06 | 3.34 (2.09-5.31) | 3.90E-07 | 1.01E-02 |
| High school | 1.00 (Reference) | 1.93 (1.36-2.76) | 2.75E-04 | 2.17 (1.21-3.88) | 9.12E-03 | 4.90 (2.49-9.62) | 3.92E-06 |  |
| College or above | 1.00 (Reference) | 1.56 (1.15-2.11) | 3.78E-03 | 2.10 (1.29-3.43) | 3.01E-03 | 3.70 (2.21-6.18) | 6.39E-07 |  |
| **C.** FI-Combined | | | | | | | | |
| **Subgroups** | **No Heart Stress or Frailty** | **Heart Stress, no Frailty** | | **Frailty, no Heart Stress** | | **Frailty, and Heart Stress** | |  |
|  |  | **HR (95% CI)** | **P-value** | **HR (95% CI)** | **P-value** | **HR (95% CI)** | **P-value** | **P-int** |
| Age group |  |  |  |  |  |  |  |  |
| 20-64 | 1.00 (Reference) | 1.92 (1.31-2.81) | 7.76E-04 | 2.65 (1.63-4.31) | 7.93E-05 | 5.60 (3.07-10.21) | 1.83E-08 | 4.76E-01 |
| 65 years or older | 1.00 (Reference) | 1.77 (1.42-2.20) | 2.39E-07 | 2.60 (1.81-3.74) | 2.66E-07 | 3.53 (2.39-5.22) | 2.18E-10 |  |
| Gender |  |  |  |  |  |  |  |  |
| Male | 1.00 (Reference) | 1.96 (1.52-2.52) | 1.77E-07 | 3.07 (2.09-4.51) | 1.13E-08 | 5.23 (3.41-8.02) | 3.84E-14 | 3.50E-01 |
| Female | 1.00 (Reference) | 1.61 (1.21-2.13) | 9.48E-04 | 2.67 (1.72-4.14) | 1.10E-05 | 3.33 (2.05-5.41) | 1.11E-06 |  |
| Body mass index |  |  |  |  |  |  |  |  |
| Normal or underweight | 1.00 (Reference) | 1.96 (1.38-2.80) | 1.90E-04 | 5.56 (2.93-10.54) | 1.47E-07 | 3.94 (2.25-6.91) | 1.64E-06 | 8.68E-02 |
| Overweight | 1.00 (Reference) | 1.44 (1.05-1.98) | 2.29E-02 | 2.24 (1.28-3.92) | 4.55E-03 | 3.79 (2.11-6.79) | 7.58E-06 |  |
| Obese | 1.00 (Reference) | 2.41 (1.71-3.39) | 4.41E-07 | 2.38 (1.51-3.75) | 1.86E-04 | 4.13 (2.19-7.77) | 1.14E-05 |  |
| Race/Ethnicity |  |  |  |  |  |  |  |  |
| White/Caucasian | 1.00 (Reference) | 1.84 (1.45-2.32) | 3.92E-07 | 2.71 (1.78-4.15) | 3.99E-06 | 9.69 (6.37-14.76) | 3.44E-26 | 2.14E-04 |
| Black/African American | 1.00 (Reference) | 2.36 (1.46-3.83) | 4.85E-04 | 3.13 (1.71-5.74) | 2.27E-04 | 3.94 (1.85-8.36) | 3.66E-04 |  |
| Others | 1.00 (Reference) | 1.58 (1.04-2.41) | 3.29E-02 | 2.59 (1.50-4.48) | 6.07E-04 | 2.26 (1.20-4.27) | 1.19E-02 |  |
| Smoking status |  |  |  |  |  |  |  |  |
| Ever | 1.00 (Reference) | 1.77 (1.41-2.23) | 1.14E-06 | 2.96 (2.08-4.22) | 1.99E-09 | 3.77 (2.47-5.74) | 6.63E-10 | 3.63E-10 |
| Never | 1.00 (Reference) | 1.88 (1.35-2.61) | 1.56E-04 | 2.81 (1.68-4.68) | 7.92E-05 | 5.55 (3.42-9.01) | 3.86E-12 |  |
| Vigorous physical activity |  |  |  |  |  |  |  |  |
| Yes | 1.00 (Reference) | 1.18 (0.69-2.00) | 5.41E-01 | 2.11 (0.63-7.11) | 2.28E-01 | 4.67 (0.49-44.68) | 1.81E-01 | 7.86E-02 |
| No | 1.00 (Reference) | 1.94 (1.58-2.37) | 1.66E-10 | 2.85 (2.11-3.84) | 8.19E-12 | 4.50 (3.25-6.22) | 8.42E-20 |  |
| Marital status |  |  |  |  |  |  |  |  |
| Married/Living with partner | 1.00 (Reference) | 1.80 (1.41-2.30) | 2.76E-06 | 2.66 (1.79-3.95) | 1.30E-06 | 3.69 (2.26-6.02) | 1.84E-07 | 6.23E-01 |
| Others | 1.00 (Reference) | 1.70 (1.25-2.32) | 6.83E-04 | 2.40 (1.53-3.75) | 1.36E-04 | 4.37 (2.82-6.76) | 3.87E-11 |  |
| Educational status |  |  |  |  |  |  |  |  |
| Less than high school | 1.00 (Reference) | 1.79 (1.33-2.42) | 1.31E-04 | 2.23 (1.44-3.47) | 3.32E-04 | 3.44 (2.22-5.32) | 3.02E-08 | 1.43E-01 |
| High school | 1.00 (Reference) | 1.96 (1.34-2.86) | 4.79E-04 | 2.55 (1.35-4.82) | 4.05E-03 | 12.05 (5.86-24.78) | 1.28E-11 |  |
| College or above | 1.00 (Reference) | 1.76 (1.28-2.42) | 5.21E-04 | 4.38 (2.65-7.24) | 8.33E-09 | 4.38 (2.24-8.56) | 1.60E-05 |  |

| **eTable 11.** Association of frailty and heart stress re-defined by a fixed cutoff with risk of all-cause mortality in NHANES and HRS. | | | | | | | | |
| --- | --- | --- | --- | --- | --- | --- | --- | --- |
| **A.** FI-Self-report | | | | | | | | |
|  |  |  | **Model 1** | | **Model 2** | | **Model 3** | |
|  |  | **Event/N (%)** | **HR (95%CI)** | **P-value** | **HR 95%CI** | **P-value** | **HR (95%CI)** | **P-value** |
| **NHANES** |  |  |  |  |  |  |  |  |
| No Frailty | No Heart Stress | 923/2672 (34.54) | Ref |  | Ref |  | Ref |  |
|  | Heart Stress | 1312/1808 (72.57) | 3.24 (2.98-3.53) | 5.25E-162 | 1.77 (1.61-1.94) | 7.44E-34 | 1.77 (1.61-1.94) | 1.89E-33 |
| Frailty | No Heart Stress | 248/520 (47.69) | 1.60 (1.39-1.85) | 3.90E-11 | 1.94 (1.68-2.23) | 6.96E-20 | 1.96 (1.70-2.27) | 3.30E-20 |
|  | Heart Stress | 626/721 (86.82) | 5.83 (5.26-6.48) | 2.33E-239 | 3.57 (3.18-4.01) | 2.04E-101 | 3.59 (3.19-4.04) | 7.30E-101 |
| **HRS** |  |  |  |  |  |  |  |  |
| No Frailty | No Heart Stress | 293/4569 (6.41) | Ref |  | Ref |  | Ref |  |
|  | Heart Stress | 696/2919 (23.84) | 4.05 (3.53-4.64) | 1.29E-89 | 2.21 (1.90-2.56) | 1.61E-25 | 2.17 (1.87-2.52) | 2.37E-24 |
| Frailty | No Heart Stress | 135/805 (16.77) | 2.80 (2.28-3.43) | 4.55E-23 | 2.86 (2.32-3.51) | 1.40E-23 | 2.61 (2.10-3.25) | 3.98E-18 |
|  | Heart Stress | 575/1195 (48.12) | 9.92 (8.62-11.43) | 3.81E-223 | 5.77 (4.92-6.77) | 5.35E-103 | 5.93 (5.04-6.97) | 2.23E-102 |
| **B.** FI-Lab | | | | | | | | |
|  |  |  | **Model 1** | | **Model 2** | | **Model 3** | |
|  |  | **Event/N (%)** | **HR (95%CI)** | **P-value** | **HR 95%CI** | **P-value** | **HR (95%CI)** | **P-value** |
| **NHANES** |  |  |  |  |  |  |  |  |
| No Frailty | No Heart Stress | 1351/8530 (15.84) | Ref |  | Ref |  | Ref |  |
|  | Heart Stress | 1571/2427 (64.73) | 6.37 (5.92-6.86) | 0.00E+00 | 1.76 (1.62-1.92) | 1.22E-39 | 1.77 (1.62-1.92) | 6.84E-40 |
| Frailty | No Heart Stress | 173/788 (21.95) | 1.41 (1.20-1.65) | 2.23E-05 | 1.98 (1.69-2.33) | 2.45E-17 | 1.96 (1.67-2.30) | 1.07E-16 |
|  | Heart Stress | 424/507 (83.63) | 12.71 (11.37-14.21) | 0.00E+00 | 3.54 (3.13-4.01) | 1.23E-88 | 3.56 (3.15-4.04) | 3.71E-89 |
| **HRS** |  |  |  |  |  |  |  |  |
| No Frailty | No Heart Stress | 303/4687 (6.46) | Ref |  | Ref |  | Ref |  |
|  | Heart Stress | 696/2973 (23.41) | 3.92 (3.42-4.49) | 1.35E-87 | 2.01 (1.73-2.33) | 2.55E-20 | 1.99 (1.72-2.31) | 6.54E-20 |
| Frailty | No Heart Stress | 112/542 (20.66) | 3.48 (2.80-4.32) | 1.85E-29 | 2.95 (2.37-3.68) | 5.39E-22 | 2.80 (2.24-3.50) | 1.09E-19 |
|  | Heart Stress | 526/1040 (50.58) | 10.63 (9.22-12.25) | 3.48E-234 | 5.86 (4.99-6.87) | 4.65E-104 | 5.68 (4.83-6.67) | 6.18E-99 |
| **C.** FI-Combined | | | | | | | | |
|  |  |  | **Model 1** | | **Model 2** | | **Model 3** | |
|  |  | **Event/N (%)** | **HR (95%CI)** | **P-value** | **HR 95%CI** | **P-value** | **HR (95%CI)** | **P-value** |
| **NHANES** |  |  |  |  |  |  |  |  |
| No Frailty | No Heart Stress | 997/2874 (34.69) | Ref |  | Ref |  | Ref |  |
|  | Heart Stress | 1414/1953 (72.40) | 3.22 (2.97-3.49) | 1.66E-172 | 1.72 (1.57-1.88) | 8.80E-33 | 1.72 (1.57-1.88) | 1.71E-32 |
| Frailty | No Heart Stress | 173/312 (55.45) | 2.02 (1.72-2.38) | 1.16E-17 | 2.55 (2.16-3.00) | 3.33E-29 | 2.61 (2.21-3.08) | 1.77E-29 |
|  | Heart Stress | 530/582 (91.07) | 7.21 (6.47-8.05) | 8.89E-275 | 4.29 (3.80-4.84) | 8.81E-124 | 4.33 (3.83-4.89) | 5.67E-123 |
| **HRS** |  |  |  |  |  |  |  |  |
| No Frailty | No Heart Stress | 299/4653 (6.43) | Ref |  | Ref |  | Ref |  |
|  | Heart Stress | 659/2929 (22.50) | 3.77 (3.29-4.32) | 1.25E-80 | 1.97 (1.70-2.29) | 5.11E-19 | 1.94 (1.67-2.26) | 3.64E-18 |
| Frailty | No Heart Stress | 124/631 (19.65) | 3.30 (2.68-4.07) | 5.13E-29 | 3.09 (2.51-3.82) | 7.42E-26 | 2.83 (2.27-3.53) | 2.61E-20 |
|  | Heart Stress | 584/1129 (51.73) | 11.02 (9.58-12.67) | 4.81E-248 | 6.53 (5.58-7.64) | 1.66E-121 | 6.76 (5.76-7.93) | 1.02E-121 |

| **eTable 12.** Association of frailty and heart stress re-defined by a fixed cutoff with risk of cause-specific mortality in NHANES. | | | | | | | | |
| --- | --- | --- | --- | --- | --- | --- | --- | --- |
| **A.** FI-Self-report | | | | | | | | |
|  |  |  | **Model 1** | | **Model 2** | | **Model 3** | |
|  |  | **Event/N (%)** | **HR (95%CI)** | **P-value** | **HR 95%CI** | **P-value** | **HR (95%CI)** | **P-value** |
| **Heart disease** |  |  |  |  |  |  |  |  |
| No Frailty | No Heart Stress | 185/1934 (9.57) |  |  |  |  |  |  |
|  | Heart Stress | 412/908 (45.37) | 6.30 (5.30-7.50) | 1.50E-95 | 2.79 (2.31-3.37) | 2.12E-26 | 2.79 (2.31-3.37) | 2.97E-26 |
| Frailty | No Heart Stress | 61/333 (18.32) | 2.06 (1.54-2.75) | 9.71E-07 | 2.65 (1.98-3.54) | 5.98E-11 | 2.37 (1.75-3.20) | 1.78E-08 |
|  | Heart Stress | 211/306 (68.95) | 13.93 (11.39-17.04) | 2.90E-145 | 8.04 (6.37-10.16) | 1.71E-68 | 8.00 (6.32-10.12) | 5.46E-67 |
| **Cerebrovascular disease** | |  |  |  |  |  |  |  |
| No Frailty | No Heart Stress | 47/1796 (2.62) |  |  |  |  |  |  |
|  | Heart Stress | 78/574 (13.59) | 5.62 (3.91-8.08) | 8.88E-21 | 1.91 (1.29-2.83) | 1.19E-03 | 1.91 (1.28-2.83) | 1.38E-03 |
| Frailty | No Heart Stress | 13/285 (4.56) | 1.76 (0.95-3.26) | 7.06E-02 | 2.28 (1.23-4.23) | 8.64E-03 | 2.07 (1.09-3.94) | 2.60E-02 |
|  | Heart Stress | 38/133 (28.57) | 13.46 (8.77-20.66) | 1.36E-32 | 5.51 (3.29-9.21) | 8.27E-11 | 4.96 (2.94-8.37) | 1.83E-09 |
| **CVD** |  |  |  |  |  |  |  |  |
| No Frailty | No Heart Stress | 232/1981 (11.71) |  |  |  |  |  |  |
|  | Heart Stress | 490/986 (49.70) | 5.80 (4.95-6.78) | 6.11E-107 | 2.52 (2.13-2.99) | 2.63E-26 | 2.52 (2.12-2.99) | 2.96E-26 |
| Frailty | No Heart Stress | 74/346 (21.39) | 1.98 (1.52-2.57) | 3.38E-07 | 2.54 (1.95-3.31) | 4.43E-12 | 2.49 (1.90-3.26) | 3.24E-11 |
|  | Heart Stress | 249/344 (72.38) | 12.36 (10.30-14.84) | 1.26E-160 | 6.69 (5.41-8.28) | 8.40E-69 | 6.65 (5.37-8.25) | 5.75E-67 |
| **Cancer** |  |  |  |  |  |  |  |  |
| No Frailty | No Heart Stress | 233/1982 (11.76) |  |  |  |  |  |  |
|  | Heart Stress | 228/724 (31.49) | 3.12 (2.60-3.75) | 2.80E-34 | 1.67 (1.37-2.03) | 4.00E-07 | 1.66 (1.36-2.02) | 6.21E-07 |
| Frailty | No Heart Stress | 72/344 (20.93) | 1.89 (1.45-2.47) | 2.14E-06 | 2.22 (1.70-2.89) | 4.63E-09 | 2.27 (1.73-2.99) | 4.16E-09 |
|  | Heart Stress | 83/178 (46.63) | 5.53 (4.30-7.11) | 1.33E-40 | 3.54 (2.69-4.68) | 3.75E-19 | 3.26 (2.46-4.32) | 2.38E-16 |
| **Others** | |  |  |  |  |  |  |  |
| No Frailty | No Heart Stress | 458/2207 (20.75) |  |  |  |  |  |  |
|  | Heart Stress | 594/1090 (54.50) | 3.56 (3.15-4.02) | 8.88E-92 | 1.73 (1.51-1.97) | 7.22E-16 | 1.72 (1.50-1.96) | 1.91E-15 |
| Frailty | No Heart Stress | 102/374 (27.27) | 1.42 (1.14-1.76) | 1.46E-03 | 1.75 (1.41-2.17) | 3.94E-07 | 1.72 (1.38-2.14) | 1.39E-06 |
|  | Heart Stress | 294/389 (75.58) | 7.25 (6.25-8.42) | 3.61E-149 | 3.97 (3.35-4.69) | 4.57E-58 | 3.77 (3.17-4.48) | 2.72E-51 |
| **B.** FI-Lab | | | | | | | | |
|  |  |  | **Model 1** | | **Model 2** | | **Model 3** | |
|  |  | **Event/N (%)** | **HR (95%CI)** | **P-value** | **HR 95%CI** | **P-value** | **HR (95%CI)** | **P-value** |
| **Heart disease** |  |  |  |  |  |  |  |  |
| No Frailty | No Heart Stress | 264/7443 (3.55) |  |  |  |  |  |  |
|  | Heart Stress | 513/1369 (37.47) | 13.59 (11.71-15.77) | 1.22E-258 | 2.91 (2.45-3.45) | 2.11E-34 | 2.91 (2.45-3.45) | 1.94E-34 |
| Frailty | No Heart Stress | 36/651 (5.53) | 1.51 (1.07-2.14) | 1.99E-02 | 2.39 (1.69-3.39) | 9.29E-07 | 2.45 (1.73-3.48) | 5.05E-07 |
|  | Heart Stress | 123/206 (59.71) | 28.65 (23.10-35.54) | 8.09E-205 | 6.45 (5.00-8.32) | 9.52E-47 | 6.42 (4.98-8.28) | 1.55E-46 |
| **Cerebrovascular disease** | |  |  |  |  |  |  |  |
| No Frailty | No Heart Stress | 76/7255 (1.05) |  |  |  |  |  |  |
|  | Heart Stress | 96/952 (10.08) | 10.37 (7.67-14.01) | 2.53E-52 | 1.91 (1.36-2.67) | 1.67E-04 | 1.90 (1.35-2.66) | 2.03E-04 |
| Frailty | No Heart Stress | 5/620 (0.81) | 0.74 (0.30-1.83) | 5.14E-01 | 1.34 (0.54-3.33) | 5.22E-01 | 1.12 (0.42-3.02) | 8.21E-01 |
|  | Heart Stress | 23/106 (21.70) | 23.54 (14.77-37.55) | 3.54E-40 | 5.46 (3.22-9.28) | 3.30E-10 | 5.39 (3.19-9.08) | 2.68E-10 |
| **CVD** |  |  |  |  |  |  |  |  |
| No Frailty | No Heart Stress | 340/7519 (4.52) |  |  |  |  |  |  |
|  | Heart Stress | 609/1465 (41.57) | 12.18 (10.66-13.91) | 2.27E-296 | 2.57 (2.20-2.99) | 2.13E-33 | 2.57 (2.20-2.99) | 1.99E-33 |
| Frailty | No Heart Stress | 41/656 (6.25) | 1.34 (0.97-1.85) | 7.67E-02 | 2.13 (1.54-2.94) | 5.20E-06 | 2.09 (1.51-2.90) | 1.06E-05 |
|  | Heart Stress | 146/229 (63.76) | 25.16 (20.69-30.59) | 1.84E-229 | 5.49 (4.37-6.91) | 3.18E-48 | 5.62 (4.47-7.06) | 1.34E-49 |
| **Cancer** |  |  |  |  |  |  |  |  |
| No Frailty | No Heart Stress | 376/7555 (4.98) |  |  |  |  |  |  |
|  | Heart Stress | 261/1117 (23.37) | 5.38 (4.59-6.30) | 1.12E-96 | 1.56 (1.30-1.86) | 9.76E-07 | 1.57 (1.32-1.88) | 5.76E-07 |
| Frailty | No Heart Stress | 43/658 (6.53) | 1.29 (0.94-1.77) | 1.10E-01 | 2.04 (1.49-2.80) | 9.69E-06 | 2.02 (1.47-2.77) | 1.43E-05 |
|  | Heart Stress | 64/147 (43.54) | 12.67 (9.72-16.52) | 1.64E-78 | 3.77 (2.82-5.04) | 3.18E-19 | 3.90 (2.91-5.23) | 8.87E-20 |
| **Others** | |  |  |  |  |  |  |  |
| No Frailty | No Heart Stress | 635/7814 (8.13) |  |  |  |  |  |  |
|  | Heart Stress | 701/1557 (45.02) | 7.38 (6.63-8.22) | 3.66E-289 | 1.80 (1.59-2.04) | 7.31E-21 | 1.80 (1.59-2.03) | 1.07E-20 |
| Frailty | No Heart Stress | 89/704 (12.64) | 1.55 (1.24-1.94) | 1.04E-04 | 2.38 (1.90-2.97) | 2.51E-14 | 2.34 (1.87-2.92) | 9.09E-14 |
|  | Heart Stress | 214/297 (72.05) | 17.32 (14.81-20.26) | 2.96E-279 | 4.49 (3.76-5.36) | 4.59E-62 | 4.41 (3.69-5.28) | 4.37E-60 |
| **C.** FI-Combined | | | | | | | | |
|  |  |  | **Model 1** | | **Model 2** | | **Model 3** | |
|  |  | **Event/N (%)** | **HR (95%CI)** | **P-value** | **HR 95%CI** | **P-value** | **HR (95%CI)** | **P-value** |
| **Heart disease** |  |  |  |  |  |  |  |  |
| No Frailty | No Heart Stress | 212/2089 (10.15) |  |  |  |  |  |  |
|  | Heart Stress | 452/991 (45.61) | 6.03 (5.12-7.10) | 1.32E-102 | 2.67 (2.24-3.20) | 2.99E-27 | 2.67 (2.24-3.20) | 3.51E-27 |
| Frailty | No Heart Stress | 35/174 (20.11) | 2.18 (1.53-3.12) | 1.93E-05 | 3.10 (2.16-4.44) | 6.90E-10 | 2.80 (1.93-4.06) | 5.77E-08 |
|  | Heart Stress | 173/225 (76.89) | 16.91 (13.77-20.77) | 1.48E-160 | 9.45 (7.42-12.02) | 1.50E-74 | 9.64 (7.55-12.31) | 1.10E-73 |
| **Cerebrovascular disease** | |  |  |  |  |  |  |  |
| No Frailty | No Heart Stress | 52/1929 (2.70) |  |  |  |  |  |  |
|  | Heart Stress | 84/623 (13.48) | 5.43 (3.84-7.68) | 8.91E-22 | 1.82 (1.25-2.66) | 1.76E-03 | 1.81 (1.24-2.65) | 2.26E-03 |
| Frailty | No Heart Stress | 8/147 (5.44) | 2.04 (0.97-4.29) | 6.12E-02 | 3.11 (1.47-6.55) | 2.94E-03 | 2.67 (1.22-5.87) | 1.42E-02 |
|  | Heart Stress | 33/85 (38.82) | 19.15 (12.37-29.65) | 5.07E-40 | 7.66 (4.52-12.98) | 4.10E-14 | 7.06 (4.14-12.01) | 6.14E-13 |
| **CVD** |  |  |  |  |  |  |  |  |
| No Frailty | No Heart Stress | 264/2141 (12.33) |  |  |  |  |  |  |
|  | Heart Stress | 536/1075 (49.86) | 5.57 (4.81-6.46) | 1.34E-114 | 2.43 (2.07-2.85) | 4.63E-27 | 2.43 (2.07-2.86) | 4.24E-27 |
| Frailty | No Heart Stress | 43/182 (23.63) | 2.14 (1.55-2.95) | 3.93E-06 | 3.10 (2.24-4.29) | 8.36E-12 | 3.02 (2.16-4.21) | 8.35E-11 |
|  | Heart Stress | 206/258 (79.84) | 15.02 (12.47-18.11) | 3.55E-178 | 7.83 (6.29-9.74) | 4.13E-76 | 7.97 (6.38-9.94) | 4.63E-75 |
| **Cancer** |  |  |  |  |  |  |  |  |
| No Frailty | No Heart Stress | 257/2134 (12.04) |  |  |  |  |  |  |
|  | Heart Stress | 249/788 (31.60) | 3.06 (2.57-3.64) | 4.08E-36 | 1.63 (1.35-1.97) | 3.19E-07 | 1.63 (1.35-1.97) | 5.02E-07 |
| Frailty | No Heart Stress | 48/187 (25.67) | 2.30 (1.69-3.14) | 1.12E-07 | 2.82 (2.07-3.85) | 4.84E-11 | 2.90 (2.10-3.99) | 8.31E-11 |
|  | Heart Stress | 62/114 (54.39) | 7.02 (5.31-9.26) | 6.55E-43 | 4.24 (3.13-5.74) | 1.14E-20 | 4.09 (3.02-5.55) | 1.09E-19 |
| **Others** | |  |  |  |  |  |  |  |
| No Frailty | No Heart Stress | 476/2353 (20.23) |  |  |  |  |  |  |
|  | Heart Stress | 629/1168 (53.85) | 3.60 (3.19-4.05) | 1.21E-97 | 1.66 (1.45-1.89) | 3.31E-14 | 1.64 (1.44-1.87) | 8.19E-14 |
| Frailty | No Heart Stress | 82/221 (37.10) | 2.13 (1.68-2.69) | 2.73E-10 | 2.75 (2.17-3.48) | 6.40E-17 | 2.76 (2.16-3.51) | 2.87E-16 |
|  | Heart Stress | 262/314 (83.44) | 9.90 (8.48-11.56) | 1.69E-185 | 5.24 (4.41-6.24) | 5.70E-78 | 5.08 (4.26-6.07) | 8.46E-72 |

| **eTable 13.** Association of frailty assessed by FI-Self-report and heart stress re-defined by a fixed cutoff with risk of all-cause mortality in subgroups. | | | | | | | | |
| --- | --- | --- | --- | --- | --- | --- | --- | --- |
| **Subgroups** | **No Heart Stress or Frailty** | **Heart Stress, no Frailty** | | **Frailty, no Heart Stress** | | **Frailty, and Heart Stress** | |  |
|  |  | **HR (95% CI)** | **P-value** | **HR (95% CI)** | **P-value** | **HR (95% CI)** | **P-value** | **P-int** |
| **NHANES** |  |  |  |  |  |  |  |  |
| Age group |  |  |  |  |  |  |  |  |
| 20-49 | 1.00 (Reference) | 4.28 (2.40-7.64) | 8.48E-07 | 2.82 (1.81-4.40) | 4.60E-06 | 5.91 (2.96-11.78) | 4.65E-07 | 1.02E-01 |
| 50-64 | 1.00 (Reference) | 2.20 (1.77-2.73) | 8.36E-13 | 2.08 (1.60-2.70) | 3.51E-08 | 4.51 (3.38-6.02) | 1.89E-24 |  |
| 65 years or older | 1.00 (Reference) | 1.62 (1.46-1.79) | 6.04E-20 | 1.64 (1.35-2.00) | 6.93E-07 | 3.15 (2.75-3.61) | 9.71E-62 |  |
| Gender |  |  |  |  |  |  |  |  |
| Male | 1.00 (Reference) | 1.88 (1.66-2.13) | 9.34E-24 | 1.95 (1.61-2.36) | 7.37E-12 | 3.67 (3.11-4.34) | 6.45E-53 | 3.83E-01 |
| Female | 1.00 (Reference) | 1.64 (1.43-1.89) | 3.97E-12 | 1.99 (1.60-2.48) | 7.04E-10 | 3.36 (2.84-3.97) | 1.47E-45 |  |
| Body mass index |  |  |  |  |  |  |  |  |
| Normal or underweight | 1.00 (Reference) | 1.67 (1.40-1.99) | 1.41E-08 | 2.09 (1.51-2.88) | 6.88E-06 | 3.08 (2.44-3.90) | 4.53E-21 | 8.86E-02 |
| Overweight | 1.00 (Reference) | 1.67 (1.44-1.94) | 8.86E-12 | 1.63 (1.26-2.11) | 1.94E-04 | 3.43 (2.76-4.26) | 7.84E-29 |  |
| Obese | 1.00 (Reference) | 1.87 (1.57-2.22) | 9.89E-13 | 1.68 (1.33-2.12) | 1.21E-05 | 3.12 (2.54-3.83) | 1.48E-27 |  |
| Race/Ethnicity |  |  |  |  |  |  |  |  |
| White/Caucasian | 1.00 (Reference) | 1.69 (1.50-1.91) | 3.49E-18 | 2.04 (1.65-2.52) | 4.50E-11 | 3.81 (3.24-4.48) | 1.48E-58 | 3.32E-02 |
| Black/African American | 1.00 (Reference) | 2.16 (1.71-2.72) | 7.68E-11 | 1.76 (1.30-2.38) | 2.78E-04 | 3.38 (2.55-4.47) | 1.77E-17 |  |
| Others | 1.00 (Reference) | 1.80 (1.48-2.18) | 3.05E-09 | 1.69 (1.30-2.20) | 1.01E-04 | 2.99 (2.37-3.77) | 1.78E-20 |  |
| Smoking status |  |  |  |  |  |  |  |  |
| Ever | 1.00 (Reference) | 1.78 (1.57-2.01) | 3.85E-20 | 1.87 (1.57-2.24) | 6.13E-12 | 3.31 (2.83-3.87) | 1.98E-51 | 1.48E-01 |
| Never | 1.00 (Reference) | 1.75 (1.52-2.02) | 8.42E-15 | 1.87 (1.46-2.40) | 7.64E-07 | 3.88 (3.24-4.65) | 1.76E-48 |  |
| Vigorous physical activity |  |  |  |  |  |  |  |  |
| Yes | 1.00 (Reference) | 1.56 (1.23-1.98) | 2.77E-04 | 1.30 (0.76-2.22) | 3.43E-01 | 2.45 (1.30-4.63) | 5.85E-03 | 9.96E-02 |
| No | 1.00 (Reference) | 1.80 (1.63-1.99) | 2.61E-30 | 1.98 (1.70-2.30) | 4.32E-19 | 3.54 (3.13-4.00) | 1.69E-90 |  |
| Marital status |  |  |  |  |  |  |  |  |
| Married/Living with partner | 1.00 (Reference) | 1.85 (1.64-2.08) | 1.77E-23 | 1.88 (1.54-2.30) | 4.21E-10 | 3.43 (2.90-4.06) | 1.03E-46 | 6.59E-01 |
| Others | 1.00 (Reference) | 1.51 (1.30-1.75) | 7.36E-08 | 1.72 (1.37-2.15) | 2.18E-06 | 3.11 (2.61-3.70) | 2.31E-37 |  |
| Educational status |  |  |  |  |  |  |  |  |
| Less than high school | 1.00 (Reference) | 1.72 (1.48-1.99) | 5.17E-13 | 1.67 (1.37-2.03) | 4.20E-07 | 2.91 (2.46-3.43) | 1.74E-36 | 3.88E-02 |
| High school | 1.00 (Reference) | 1.77 (1.47-2.13) | 1.09E-09 | 1.53 (1.09-2.14) | 1.34E-02 | 4.57 (3.57-5.85) | 1.24E-33 |  |
| College or above | 1.00 (Reference) | 1.79 (1.53-2.10) | 6.06E-13 | 2.52 (1.89-3.35) | 2.54E-10 | 3.54 (2.77-4.52) | 6.85E-24 |  |
| **HRS** |  |  |  |  |  |  |  |  |
| Age group |  |  |  |  |  |  |  |  |
| 50-64 | 1.00 (Reference) | 3.37 (2.42-4.70) | 8.15E-13 | 3.18 (2.13-4.74) | 1.59E-08 | 9.03 (6.24-13.06) | 1.57E-31 | 5.78E-02 |
| 65 years or older | 1.00 (Reference) | 1.97 (1.67-2.32) | 3.90E-16 | 2.46 (1.89-3.19) | 1.62E-11 | 5.31 (4.44-6.34) | 6.85E-76 |  |
| Gender |  |  |  |  |  |  |  |  |
| Male | 1.00 (Reference) | 2.25 (1.84-2.75) | 2.66E-15 | 2.15 (1.54-2.98) | 5.56E-06 | 5.66 (4.48-7.13) | 1.60E-48 | 9.08E-01 |
| Female | 1.00 (Reference) | 2.07 (1.66-2.59) | 1.41E-10 | 3.03 (2.25-4.09) | 3.20E-13 | 5.75 (4.56-7.25) | 2.41E-49 |  |
| Body mass index |  |  |  |  |  |  |  |  |
| Normal or underweight | 1.00 (Reference) | 2.29 (1.75-2.98) | 1.03E-09 | 3.84 (2.45-6.04) | 5.06E-09 | 7.52 (5.54-10.20) | 2.37E-38 | 2.87E-02 |
| Overweight | 1.00 (Reference) | 1.97 (1.53-2.53) | 1.21E-07 | 2.10 (1.37-3.21) | 6.37E-04 | 4.69 (3.52-6.26) | 6.29E-26 |  |
| Obese | 1.00 (Reference) | 2.26 (1.73-2.95) | 2.60E-09 | 2.29 (1.67-3.13) | 2.53E-07 | 4.64 (3.54-6.08) | 1.57E-28 |  |
| Race/Ethnicity |  |  |  |  |  |  |  |  |
| White/Caucasian | 1.00 (Reference) | 2.18 (1.83-2.59) | 1.50E-18 | 3.72 (2.87-4.83) | 4.61E-23 | 6.19 (5.08-7.54) | 2.90E-73 | 9.64E-01 |
| Black/African American | 1.00 (Reference) | 2.34 (1.65-3.32) | 2.10E-06 | 1.49 (0.96-2.32) | 7.70E-02 | 4.70 (3.34-6.63) | 8.58E-19 |  |
| Others | 1.00 (Reference) | 1.88 (1.06-3.32) | 3.08E-02 | 0.90 (0.40-2.05) | 8.06E-01 | 4.40 (2.50-7.73) | 2.61E-07 |  |
| Smoking status |  |  |  |  |  |  |  |  |
| Ever | 1.00 (Reference) | 2.43 (2.00-2.96) | 6.58E-19 | 3.06 (2.36-3.98) | 5.68E-17 | 6.06 (4.91-7.47) | 7.44E-64 | 2.55E-01 |
| Never | 1.00 (Reference) | 1.84 (1.46-2.32) | 2.69E-07 | 1.78 (1.19-2.69) | 5.54E-03 | 5.09 (3.89-6.64) | 7.43E-33 |  |
| Vigorous physical activity |  |  |  |  |  |  |  |  |
| Yes | 1.00 (Reference) | 2.17 (1.71-2.76) | 2.48E-10 | 2.83 (1.74-4.61) | 2.87E-05 | 4.51 (3.18-6.39) | 2.99E-17 | 7.69E-01 |
| No | 1.00 (Reference) | 2.14 (1.77-2.60) | 5.94E-15 | 2.25 (1.75-2.90) | 2.45E-10 | 5.12 (4.23-6.19) | 6.46E-63 |  |
| Marital status |  |  |  |  |  |  |  |  |
| Married/Living with partner | 1.00 (Reference) | 2.16 (1.77-2.63) | 1.92E-14 | 3.04 (2.28-4.05) | 2.40E-14 | 7.17 (5.77-8.91) | 6.83E-71 | 6.86E-01 |
| Others | 1.00 (Reference) | 2.06 (1.64-2.58) | 5.14E-10 | 2.09 (1.50-2.93) | 1.66E-05 | 4.12 (3.24-5.25) | 1.81E-30 |  |
| Educational status |  |  |  |  |  |  |  |  |
| Less than high school | 1.00 (Reference) | 1.95 (1.43-2.66) | 2.28E-05 | 1.61 (1.07-2.42) | 2.14E-02 | 3.24 (2.38-4.41) | 8.94E-14 | 1.46E-12 |
| High school | 1.00 (Reference) | 2.16 (1.68-2.79) | 2.93E-09 | 2.59 (1.79-3.74) | 4.46E-07 | 6.11 (4.66-8.02) | 5.17E-39 |  |
| College or above | 1.00 (Reference) | 2.29 (1.83-2.88) | 9.55E-13 | 3.81 (2.69-5.40) | 4.69E-14 | 7.24 (5.53-9.46) | 2.38E-47 |  |

| **eTable 14.** Association of frailty assessed by FI-Lab and heart stress re-defined by a fixed cutoff with risk of all-cause mortality in subgroups. | | | | | | | | |
| --- | --- | --- | --- | --- | --- | --- | --- | --- |
| **Subgroups** | **No Heart Stress or Frailty** | **Heart Stress, no Frailty** | | **Frailty, no Heart Stress** | | **Frailty, and Heart Stress** | |  |
|  |  | **HR (95% CI)** | **P-value** | **HR (95% CI)** | **P-value** | **HR (95% CI)** | **P-value** | **P-int** |
| **NHANES** |  |  |  |  |  |  |  |  |
| Age group |  |  |  |  |  |  |  |  |
| 20-49 | 1.00 (Reference) | 2.20 (1.46-3.31) | 1.66E-04 | 2.41 (1.76-3.30) | 4.50E-08 | 8.26 (5.19-13.16) | 5.55E-19 | 2.18E-02 |
| 50-64 | 1.00 (Reference) | 2.19 (1.83-2.62) | 1.18E-17 | 2.04 (1.53-2.71) | 1.26E-06 | 6.77 (5.11-8.97) | 1.46E-40 |  |
| 65 years or older | 1.00 (Reference) | 1.63 (1.48-1.80) | 1.62E-22 | 1.71 (1.34-2.19) | 2.06E-05 | 3.04 (2.63-3.53) | 7.96E-50 |  |
| Gender |  |  |  |  |  |  |  |  |
| Male | 1.00 (Reference) | 1.88 (1.67-2.12) | 1.78E-25 | 2.12 (1.75-2.58) | 3.99E-14 | 3.26 (2.78-3.82) | 1.78E-47 | 2.92E-02 |
| Female | 1.00 (Reference) | 1.67 (1.48-1.88) | 7.42E-17 | 1.80 (1.37-2.37) | 2.47E-05 | 4.40 (3.60-5.37) | 5.05E-48 |  |
| Body mass index |  |  |  |  |  |  |  |  |
| Normal or underweight | 1.00 (Reference) | 1.56 (1.33-1.85) | 1.09E-07 | 2.14 (1.57-2.90) | 1.19E-06 | 2.64 (2.07-3.36) | 3.44E-15 | 5.32E-03 |
| Overweight | 1.00 (Reference) | 1.71 (1.49-1.97) | 2.36E-14 | 2.37 (1.79-3.14) | 1.31E-09 | 3.40 (2.74-4.22) | 8.03E-29 |  |
| Obese | 1.00 (Reference) | 1.82 (1.56-2.11) | 7.91E-15 | 1.57 (1.19-2.06) | 1.37E-03 | 3.71 (2.94-4.69) | 3.63E-28 |  |
| Race/Ethnicity |  |  |  |  |  |  |  |  |
| White/Caucasian | 1.00 (Reference) | 1.73 (1.55-1.93) | 3.65E-23 | 1.77 (1.37-2.29) | 1.08E-05 | 3.24 (2.73-3.85) | 1.04E-40 | 5.81E-01 |
| Black/African American | 1.00 (Reference) | 2.03 (1.63-2.53) | 3.30E-10 | 1.82 (1.35-2.45) | 8.65E-05 | 3.74 (2.89-4.84) | 1.16E-23 |  |
| Others | 1.00 (Reference) | 1.78 (1.50-2.11) | 4.71E-11 | 2.17 (1.63-2.89) | 1.32E-07 | 3.85 (2.98-4.97) | 6.68E-25 |  |
| Smoking status |  |  |  |  |  |  |  |  |
| Ever | 1.00 (Reference) | 1.72 (1.54-1.92) | 1.46E-21 | 1.85 (1.50-2.27) | 5.79E-09 | 3.44 (2.94-4.03) | 8.04E-53 | 4.47E-01 |
| Never | 1.00 (Reference) | 1.84 (1.61-2.09) | 5.72E-20 | 2.20 (1.71-2.84) | 9.47E-10 | 3.84 (3.14-4.70) | 6.36E-39 |  |
| Vigorous physical activity |  |  |  |  |  |  |  |  |
| Yes | 1.00 (Reference) | 1.56 (1.24-1.97) | 1.58E-04 | 1.56 (1.03-2.35) | 3.51E-02 | 3.06 (1.97-4.76) | 6.45E-07 | 5.64E-02 |
| No | 1.00 (Reference) | 1.78 (1.62-1.95) | 3.75E-35 | 2.05 (1.72-2.44) | 5.10E-16 | 3.54 (3.11-4.04) | 5.28E-79 |  |
| Marital status |  |  |  |  |  |  |  |  |
| Married/Living with partner | 1.00 (Reference) | 1.84 (1.64-2.05) | 1.33E-26 | 1.90 (1.52-2.38) | 1.83E-08 | 3.38 (2.83-4.04) | 2.46E-41 | 8.93E-01 |
| Others | 1.00 (Reference) | 1.59 (1.39-1.82) | 1.19E-11 | 1.79 (1.40-2.28) | 2.56E-06 | 3.19 (2.66-3.84) | 3.29E-35 |  |
| Educational status |  |  |  |  |  |  |  |  |
| Less than high school | 1.00 (Reference) | 1.71 (1.51-1.95) | 1.90E-16 | 1.96 (1.55-2.48) | 1.74E-08 | 3.18 (2.65-3.81) | 8.03E-36 | 1.98E-03 |
| High school | 1.00 (Reference) | 1.87 (1.58-2.23) | 1.04E-12 | 2.41 (1.74-3.33) | 1.27E-07 | 4.28 (3.26-5.61) | 9.40E-26 |  |
| College or above | 1.00 (Reference) | 1.68 (1.45-1.95) | 5.94E-12 | 1.75 (1.29-2.37) | 3.06E-04 | 3.44 (2.75-4.31) | 3.62E-27 |  |
| **HRS** |  |  |  |  |  |  |  |  |
| Age group |  |  |  |  |  |  |  |  |
| 50-64 | 1.00 (Reference) | 2.56 (1.79-3.66) | 2.56E-07 | 4.55 (3.06-6.75) | 6.18E-14 | 12.52 (8.87-17.67) | 6.18E-47 | 3.90E-06 |
| 65 years or older | 1.00 (Reference) | 1.89 (1.61-2.22) | 1.01E-14 | 2.29 (1.75-3.00) | 2.01E-09 | 4.72 (3.96-5.62) | 1.20E-67 |  |
| Gender |  |  |  |  |  |  |  |  |
| Male | 1.00 (Reference) | 2.24 (1.79-2.79) | 1.35E-12 | 2.83 (2.11-3.81) | 5.62E-12 | 5.61 (4.47-7.05) | 4.45E-50 | 4.29E-01 |
| Female | 1.00 (Reference) | 1.81 (1.49-2.21) | 4.27E-09 | 2.74 (1.95-3.87) | 8.33E-09 | 5.54 (4.41-6.97) | 1.37E-48 |  |
| Body mass index |  |  |  |  |  |  |  |  |
| Normal or underweight | 1.00 (Reference) | 1.95 (1.49-2.54) | 9.26E-07 | 2.95 (1.82-4.76) | 9.87E-06 | 5.61 (4.19-7.51) | 3.57E-31 | 5.62E-01 |
| Overweight | 1.00 (Reference) | 1.95 (1.50-2.54) | 5.74E-07 | 2.76 (1.82-4.17) | 1.48E-06 | 6.00 (4.47-8.05) | 6.52E-33 |  |
| Obese | 1.00 (Reference) | 2.01 (1.56-2.58) | 5.35E-08 | 2.63 (1.89-3.64) | 6.96E-09 | 4.82 (3.69-6.28) | 5.59E-31 |  |
| Race/Ethnicity |  |  |  |  |  |  |  |  |
| White/Caucasian | 1.00 (Reference) | 1.83 (1.54-2.17) | 3.34E-12 | 2.62 (1.96-3.50) | 6.86E-11 | 5.58 (4.61-6.74) | 2.57E-70 | 8.16E-01 |
| Black/African American | 1.00 (Reference) | 2.83 (1.96-4.10) | 3.55E-08 | 3.47 (2.27-5.30) | 8.91E-09 | 6.24 (4.31-9.05) | 4.18E-22 |  |
| Others | 1.00 (Reference) | 2.24 (1.30-3.85) | 3.76E-03 | 2.51 (1.18-5.36) | 1.71E-02 | 4.75 (2.65-8.53) | 1.68E-07 |  |
| Smoking status |  |  |  |  |  |  |  |  |
| Ever | 1.00 (Reference) | 2.05 (1.70-2.48) | 1.06E-13 | 2.45 (1.85-3.23) | 3.59E-10 | 4.98 (4.05-6.12) | 2.04E-52 | 3.12E-01 |
| Never | 1.00 (Reference) | 1.81 (1.42-2.30) | 1.32E-06 | 3.41 (2.35-4.94) | 1.03E-10 | 6.65 (5.12-8.64) | 9.96E-46 |  |
| Vigorous physical activity |  |  |  |  |  |  |  |  |
| Yes | 1.00 (Reference) | 2.19 (1.68-2.85) | 5.67E-09 | 3.98 (2.70-5.87) | 3.40E-12 | 4.78 (3.47-6.58) | 7.80E-22 | 5.50E-01 |
| No | 1.00 (Reference) | 1.85 (1.54-2.21) | 2.09E-11 | 2.16 (1.64-2.84) | 3.19E-08 | 5.19 (4.31-6.27) | 2.76E-66 |  |
| Marital status |  |  |  |  |  |  |  |  |
| Married/Living with partner | 1.00 (Reference) | 1.95 (1.60-2.38) | 3.92E-11 | 2.98 (2.23-3.99) | 1.69E-13 | 6.14 (4.95-7.62) | 7.50E-61 | 9.67E-01 |
| Others | 1.00 (Reference) | 1.95 (1.56-2.43) | 3.83E-09 | 2.45 (1.72-3.49) | 6.74E-07 | 4.79 (3.76-6.09) | 2.91E-37 |  |
| Educational status |  |  |  |  |  |  |  |  |
| Less than high school | 1.00 (Reference) | 2.11 (1.56-2.85) | 1.19E-06 | 2.87 (1.91-4.32) | 4.05E-07 | 4.39 (3.21-6.00) | 1.92E-20 | 8.26E-03 |
| High school | 1.00 (Reference) | 2.01 (1.56-2.58) | 6.03E-08 | 2.37 (1.58-3.55) | 2.94E-05 | 5.65 (4.31-7.41) | 6.58E-36 |  |
| College or above | 1.00 (Reference) | 1.92 (1.52-2.42) | 3.07E-08 | 2.95 (2.07-4.21) | 2.22E-09 | 6.61 (5.10-8.56) | 2.13E-46 |  |

| **eTable 15.** Association of frailty assessed by FI-Combined and heart stress re-defined by a fixed cutoff with risk of all-cause mortality in subgroups. | | | | | | | | |
| --- | --- | --- | --- | --- | --- | --- | --- | --- |
| **Subgroups** | **No Heart Stress or Frailty** | **Heart Stress, no Frailty** | | **Frailty, no Heart Stress** | | **Frailty, and Heart Stress** | |  |
|  |  | **HR (95% CI)** | **P-value** | **HR (95% CI)** | **P-value** | **HR (95% CI)** | **P-value** | **P-int** |
| **NHANES** |  |  |  |  |  |  |  |  |
| Age group |  |  |  |  |  |  |  |  |
| 20-49 | 1.00 (Reference) | 3.33 (1.88-5.89) | 3.86E-05 | 3.65 (2.31-5.78) | 3.05E-08 | 10.32 (5.48-19.43) | 4.78E-13 | 1.66E-02 |
| 50-64 | 1.00 (Reference) | 2.03 (1.65-2.50) | 2.92E-11 | 2.49 (1.84-3.35) | 2.78E-09 | 5.82 (4.38-7.71) | 2.74E-34 |  |
| 65 years or older | 1.00 (Reference) | 1.60 (1.44-1.77) | 4.86E-20 | 2.22 (1.76-2.80) | 2.14E-11 | 3.68 (3.19-4.25) | 2.25E-71 |  |
| Gender |  |  |  |  |  |  |  |  |
| Male | 1.00 (Reference) | 1.78 (1.57-2.00) | 2.15E-20 | 2.59 (2.09-3.22) | 9.85E-18 | 4.44 (3.75-5.25) | 1.31E-67 | 5.73E-01 |
| Female | 1.00 (Reference) | 1.66 (1.46-1.90) | 3.48E-14 | 2.62 (2.02-3.39) | 3.95E-13 | 3.99 (3.34-4.77) | 2.09E-52 |  |
| Body mass index |  |  |  |  |  |  |  |  |
| Normal or underweight | 1.00 (Reference) | 1.55 (1.30-1.83) | 6.49E-07 | 3.74 (2.57-5.45) | 6.31E-12 | 4.01 (3.14-5.13) | 9.93E-29 | 6.52E-02 |
| Overweight | 1.00 (Reference) | 1.65 (1.43-1.90) | 7.50E-12 | 2.43 (1.77-3.34) | 4.17E-08 | 3.97 (3.16-4.99) | 5.00E-32 |  |
| Obese | 1.00 (Reference) | 1.84 (1.56-2.16) | 2.82E-13 | 1.82 (1.40-2.37) | 8.58E-06 | 3.69 (2.98-4.58) | 6.69E-33 |  |
| Race/Ethnicity |  |  |  |  |  |  |  |  |
| White/Caucasian | 1.00 (Reference) | 1.67 (1.49-1.87) | 1.65E-18 | 2.47 (1.90-3.21) | 1.34E-11 | 4.56 (3.85-5.41) | 2.21E-68 | 4.42E-02 |
| Black/African American | 1.00 (Reference) | 1.92 (1.52-2.41) | 3.09E-08 | 2.61 (1.84-3.70) | 7.45E-08 | 4.41 (3.36-5.79) | 1.02E-26 |  |
| Others | 1.00 (Reference) | 1.81 (1.50-2.18) | 4.44E-10 | 2.38 (1.78-3.19) | 5.27E-09 | 3.43 (2.69-4.37) | 3.11E-23 |  |
| Smoking status |  |  |  |  |  |  |  |  |
| Ever | 1.00 (Reference) | 1.70 (1.51-1.91) | 1.23E-18 | 2.56 (2.09-3.15) | 3.36E-19 | 4.30 (3.67-5.05) | 1.15E-71 | 1.69E-01 |
| Never | 1.00 (Reference) | 1.73 (1.51-1.99) | 3.09E-15 | 2.32 (1.73-3.10) | 1.52E-08 | 4.32 (3.58-5.23) | 7.56E-52 |  |
| Vigorous physical activity |  |  |  |  |  |  |  |  |
| Yes | 1.00 (Reference) | 1.46 (1.15-1.85) | 1.96E-03 | 1.65 (0.81-3.36) | 1.64E-01 | 9.68 (5.13-18.26) | 2.37E-12 | 9.49E-02 |
| No | 1.00 (Reference) | 1.76 (1.60-1.94) | 3.15E-30 | 2.61 (2.20-3.11) | 9.40E-28 | 4.18 (3.68-4.75) | 2.79E-108 |  |
| Marital status |  |  |  |  |  |  |  |  |
| Married/Living with partner | 1.00 (Reference) | 1.78 (1.58-2.00) | 7.60E-22 | 2.33 (1.83-2.97) | 8.57E-12 | 4.31 (3.62-5.13) | 6.55E-60 | 9.10E-01 |
| Others | 1.00 (Reference) | 1.52 (1.31-1.75) | 1.07E-08 | 2.38 (1.86-3.05) | 7.13E-12 | 3.80 (3.17-4.55) | 1.01E-47 |  |
| Educational status |  |  |  |  |  |  |  |  |
| Less than high school | 1.00 (Reference) | 1.72 (1.50-1.98) | 1.16E-14 | 2.28 (1.83-2.85) | 3.13E-13 | 3.43 (2.89-4.08) | 2.20E-44 | 1.74E-02 |
| High school | 1.00 (Reference) | 1.77 (1.48-2.12) | 4.85E-10 | 2.64 (1.80-3.87) | 6.16E-07 | 6.39 (4.87-8.37) | 5.56E-41 |  |
| College or above | 1.00 (Reference) | 1.64 (1.40-1.92) | 4.51E-10 | 2.63 (1.84-3.77) | 1.35E-07 | 4.75 (3.73-6.05) | 9.36E-37 |  |
| **HRS** |  |  |  |  |  |  |  |  |
| Age group |  |  |  |  |  |  |  |  |
| 50-64 | 1.00 (Reference) | 2.78 (1.95-3.95) | 1.30E-08 | 4.00 (2.67-6.01) | 2.32E-11 | 11.34 (7.98-16.12) | 7.93E-42 | 3.83E-03 |
| 65 years or older | 1.00 (Reference) | 1.81 (1.54-2.14) | 9.49E-13 | 2.48 (1.90-3.23) | 2.67E-11 | 5.83 (4.89-6.95) | 1.11E-86 |  |
| Gender |  |  |  |  |  |  |  |  |
| Male | 1.00 (Reference) | 2.07 (1.68-2.55) | 5.35E-12 | 2.41 (1.74-3.33) | 1.14E-07 | 6.68 (5.33-8.38) | 3.48E-61 | 9.99E-01 |
| Female | 1.00 (Reference) | 1.81 (1.46-2.25) | 8.79E-08 | 3.25 (2.39-4.43) | 6.23E-14 | 6.41 (5.10-8.06) | 5.58E-57 |  |
| Body mass index |  |  |  |  |  |  |  |  |
| Normal or underweight | 1.00 (Reference) | 2.05 (1.57-2.69) | 1.75E-07 | 4.18 (2.67-6.53) | 3.90E-10 | 8.78 (6.47-11.91) | 2.90E-44 | 1.59E-02 |
| Overweight | 1.00 (Reference) | 1.89 (1.46-2.43) | 9.81E-07 | 2.13 (1.36-3.34) | 9.37E-04 | 5.85 (4.39-7.78) | 9.79E-34 |  |
| Obese | 1.00 (Reference) | 1.94 (1.48-2.53) | 1.31E-06 | 2.56 (1.86-3.52) | 6.47E-09 | 5.04 (3.86-6.56) | 4.99E-33 |  |
| Race/Ethnicity |  |  |  |  |  |  |  |  |
| White/Caucasian | 1.00 (Reference) | 1.82 (1.54-2.17) | 6.20E-12 | 3.28 (2.49-4.31) | 2.79E-17 | 7.10 (5.86-8.58) | 2.13E-90 | 8.88E-01 |
| Black/African American | 1.00 (Reference) | 2.54 (1.75-3.69) | 1.04E-06 | 2.50 (1.63-3.83) | 2.63E-05 | 5.76 (4.03-8.23) | 6.16E-22 |  |
| Others | 1.00 (Reference) | 2.21 (1.25-3.91) | 6.26E-03 | 1.46 (0.64-3.33) | 3.68E-01 | 4.71 (2.60-8.56) | 3.44E-07 |  |
| Smoking status |  |  |  |  |  |  |  |  |
| Ever | 1.00 (Reference) | 2.02 (1.66-2.45) | 1.59E-12 | 2.87 (2.19-3.76) | 3.09E-14 | 6.59 (5.38-8.07) | 1.57E-74 | 3.99E-01 |
| Never | 1.00 (Reference) | 1.85 (1.46-2.35) | 4.21E-07 | 2.76 (1.88-4.06) | 2.61E-07 | 6.18 (4.72-8.10) | 7.27E-40 |  |
| Vigorous physical activity |  |  |  |  |  |  |  |  |
| Yes | 1.00 (Reference) | 2.10 (1.64-2.68) | 2.62E-09 | 3.79 (2.36-6.08) | 3.42E-08 | 5.41 (3.83-7.65) | 1.23E-21 | 9.00E-01 |
| No | 1.00 (Reference) | 1.85 (1.52-2.23) | 3.23E-10 | 2.32 (1.80-3.00) | 1.01E-10 | 5.71 (4.74-6.89) | 2.03E-74 |  |
| Marital status |  |  |  |  |  |  |  |  |
| Married/Living with partner | 1.00 (Reference) | 1.88 (1.55-2.29) | 2.44E-10 | 2.91 (2.15-3.93) | 3.62E-12 | 7.77 (6.28-9.60) | 2.09E-80 | 8.20E-01 |
| Others | 1.00 (Reference) | 1.93 (1.53-2.43) | 3.27E-08 | 2.68 (1.93-3.73) | 4.48E-09 | 4.99 (3.91-6.37) | 3.55E-38 |  |
| Educational status |  |  |  |  |  |  |  |  |
| Less than high school | 1.00 (Reference) | 1.91 (1.40-2.63) | 5.71E-05 | 2.24 (1.50-3.35) | 8.28E-05 | 3.88 (2.84-5.31) | 2.11E-17 | 4.94E-11 |
| High school | 1.00 (Reference) | 2.05 (1.58-2.65) | 6.34E-08 | 3.04 (2.09-4.42) | 6.84E-09 | 7.16 (5.46-9.40) | 9.25E-46 |  |
| College or above | 1.00 (Reference) | 1.88 (1.50-2.36) | 3.87E-08 | 3.18 (2.20-4.59) | 8.09E-10 | 7.93 (6.13-10.28) | 1.90E-55 |  |

| **eTable 16.** Association of frailty and heart stress re-defined by a fixed cutoff with risk of CVD mortality in subgroups in NHANES. | | | | | | | | |
| --- | --- | --- | --- | --- | --- | --- | --- | --- |
| **A.** FI-Self-report | | | | | | | | |
| **Subgroups** | **No Heart Stress or Frailty** | **Heart Stress, no Frailty** | | **Frailty, no Heart Stress** | | **Frailty, and Heart Stress** | |  |
|  |  | **HR (95% CI)** | **P-value** | **HR (95% CI)** | **P-value** | **HR (95% CI)** | **P-value** | **P-int** |
| Age group |  |  |  |  |  |  |  |  |
| 20-64 | 1.00 (Reference) | 4.31 (2.99-6.20) | 3.64E-15 | 2.26 (1.46-3.52) | 2.80E-04 | 9.01 (5.68-14.28) | 8.71E-21 | 8.00E-01 |
| 65 years or older | 1.00 (Reference) | 2.12 (1.75-2.57) | 1.68E-14 | 2.21 (1.56-3.12) | 6.98E-06 | 5.50 (4.27-7.08) | 8.94E-40 |  |
| Gender |  |  |  |  |  |  |  |  |
| Male | 1.00 (Reference) | 2.88 (2.30-3.60) | 2.09E-20 | 2.27 (1.61-3.20) | 3.00E-06 | 7.65 (5.70-10.27) | 1.10E-41 | 2.93E-01 |
| Female | 1.00 (Reference) | 2.22 (1.69-2.90) | 7.49E-09 | 2.22 (1.43-3.46) | 4.10E-04 | 5.42 (3.92-7.49) | 1.63E-24 |  |
| Body mass index |  |  |  |  |  |  |  |  |
| Normal or underweight | 1.00 (Reference) | 2.72 (1.93-3.85) | 1.29E-08 | 2.88 (1.49-5.58) | 1.63E-03 | 5.59 (3.52-8.88) | 3.15E-13 | 4.61E-02 |
| Overweight | 1.00 (Reference) | 2.20 (1.67-2.90) | 2.22E-08 | 2.28 (1.45-3.59) | 3.64E-04 | 5.81 (3.89-8.70) | 1.06E-17 |  |
| Obese | 1.00 (Reference) | 2.58 (1.90-3.49) | 9.05E-10 | 1.74 (1.13-2.69) | 1.24E-02 | 5.55 (3.85-8.01) | 5.03E-20 |  |
| Race/Ethnicity |  |  |  |  |  |  |  |  |
| White/Caucasian | 1.00 (Reference) | 2.27 (1.82-2.84) | 2.78E-13 | 2.04 (1.39-2.99) | 2.63E-04 | 6.85 (5.04-9.33) | 1.92E-34 | 1.27E-01 |
| Black/African American | 1.00 (Reference) | 3.50 (2.33-5.25) | 1.41E-09 | 1.73 (0.95-3.12) | 7.16E-02 | 5.99 (3.78-9.50) | 2.53E-14 |  |
| Others | 1.00 (Reference) | 2.92 (2.02-4.24) | 1.45E-08 | 2.66 (1.57-4.52) | 2.88E-04 | 5.03 (3.23-7.83) | 8.14E-13 |  |
| Smoking status |  |  |  |  |  |  |  |  |
| Ever | 1.00 (Reference) | 2.59 (2.06-3.26) | 4.94E-16 | 2.22 (1.58-3.12) | 3.84E-06 | 6.38 (4.79-8.49) | 6.23E-37 | 3.35E-01 |
| Never | 1.00 (Reference) | 2.47 (1.91-3.20) | 7.52E-12 | 2.48 (1.56-3.93) | 1.12E-04 | 6.62 (4.72-9.27) | 5.28E-28 |  |
| Vigorous physical activity |  |  |  |  |  |  |  |  |
| Yes | 1.00 (Reference) | 2.88 (1.82-4.56) | 6.24E-06 | 0.81 (0.26-2.47) | 7.06E-01 | 4.15 (1.54-11.19) | 4.96E-03 | 9.83E-03 |
| No | 1.00 (Reference) | 2.45 (2.04-2.95) | 1.86E-21 | 2.51 (1.90-3.32) | 9.25E-11 | 6.36 (5.09-7.96) | 3.26E-59 |  |
| Marital status |  |  |  |  |  |  |  |  |
| Married/Living with partner | 1.00 (Reference) | 2.46 (1.98-3.06) | 3.40E-16 | 2.28 (1.57-3.30) | 1.46E-05 | 5.18 (3.80-7.05) | 1.85E-25 | 4.01E-01 |
| Others | 1.00 (Reference) | 2.12 (1.59-2.83) | 3.38E-07 | 1.84 (1.19-2.84) | 5.74E-03 | 6.18 (4.46-8.55) | 5.69E-28 |  |
| Educational status |  |  |  |  |  |  |  |  |
| Less than high school | 1.00 (Reference) | 2.61 (1.99-3.41) | 2.97E-12 | 2.26 (1.58-3.25) | 1.01E-05 | 5.44 (4.03-7.33) | 1.09E-28 | 2.31E-02 |
| High school | 1.00 (Reference) | 2.19 (1.56-3.06) | 5.09E-06 | 0.98 (0.44-2.18) | 9.63E-01 | 7.75 (4.96-12.13) | 3.04E-19 |  |
| College or above | 1.00 (Reference) | 2.57 (1.89-3.48) | 1.28E-09 | 3.05 (1.81-5.17) | 3.08E-05 | 5.95 (3.73-9.49) | 7.33E-14 |  |
| **B.** FI-Lab | | | | | | | | |
| **Subgroups** | **No Heart Stress or Frailty** | **Heart Stress, no Frailty** | | **Frailty, no Heart Stress** | | **Frailty, and Heart Stress** | |  |
|  |  | **HR (95% CI)** | **P-value** | **HR (95% CI)** | **P-value** | **HR (95% CI)** | **P-value** | **P-int** |
| Age group |  |  |  |  |  |  |  |  |
| 20-64 | 1.00 (Reference) | 3.57 (2.66-4.78) | 2.08E-17 | 1.69 (1.05-2.74) | 3.13E-02 | 10.26 (6.62-15.89) | 1.85E-25 | 3.27E-01 |
| 65 years or older | 1.00 (Reference) | 2.23 (1.86-2.67) | 2.61E-18 | 2.30 (1.45-3.63) | 3.83E-04 | 4.35 (3.30-5.74) | 1.82E-25 |  |
| Gender |  |  |  |  |  |  |  |  |
| Male | 1.00 (Reference) | 3.08 (2.49-3.81) | 2.44E-25 | 2.31 (1.55-3.43) | 3.45E-05 | 5.05 (3.78-6.74) | 6.98E-28 | 3.82E-02 |
| Female | 1.00 (Reference) | 2.21 (1.76-2.77) | 8.26E-12 | 2.08 (1.14-3.79) | 1.71E-02 | 6.61 (4.52-9.66) | 1.97E-22 |  |
| Body mass index |  |  |  |  |  |  |  |  |
| Normal or underweight | 1.00 (Reference) | 2.27 (1.65-3.12) | 4.05E-07 | 2.13 (1.09-4.17) | 2.68E-02 | 3.37 (2.06-5.52) | 1.27E-06 | 1.19E-01 |
| Overweight | 1.00 (Reference) | 2.26 (1.76-2.90) | 1.80E-10 | 2.99 (1.66-5.37) | 2.45E-04 | 5.41 (3.72-7.87) | 1.00E-18 |  |
| Obese | 1.00 (Reference) | 2.78 (2.13-3.62) | 4.10E-14 | 1.51 (0.85-2.70) | 1.61E-01 | 5.99 (3.93-9.13) | 8.55E-17 |  |
| Race/Ethnicity |  |  |  |  |  |  |  |  |
| White/Caucasian | 1.00 (Reference) | 2.35 (1.93-2.87) | 2.98E-17 | 2.15 (1.30-3.56) | 2.93E-03 | 4.79 (3.50-6.58) | 2.41E-22 | 4.78E-01 |
| Black/African American | 1.00 (Reference) | 3.37 (2.27-4.99) | 1.63E-09 | 2.21 (1.25-3.88) | 6.06E-03 | 6.69 (4.20-10.64) | 1.08E-15 |  |
| Others | 1.00 (Reference) | 2.80 (2.05-3.82) | 9.03E-11 | 1.62 (0.78-3.38) | 1.97E-01 | 4.78 (2.87-7.96) | 1.92E-09 |  |
| Smoking status |  |  |  |  |  |  |  |  |
| Ever | 1.00 (Reference) | 2.61 (2.12-3.20) | 5.87E-20 | 2.14 (1.40-3.28) | 4.55E-04 | 5.98 (4.46-8.00) | 3.16E-33 | 6.44E-01 |
| Never | 1.00 (Reference) | 2.52 (2.00-3.17) | 5.98E-15 | 2.17 (1.29-3.65) | 3.71E-03 | 4.94 (3.41-7.15) | 2.54E-17 |  |
| Vigorous physical activity |  |  |  |  |  |  |  |  |
| Yes | 1.00 (Reference) | 3.28 (2.13-5.05) | 7.36E-08 | 1.79 (0.81-3.96) | 1.48E-01 | 5.08 (1.99-13.00) | 6.90E-04 | 7.90E-01 |
| No | 1.00 (Reference) | 2.46 (2.08-2.89) | 1.00E-26 | 2.18 (1.52-3.13) | 2.35E-05 | 5.32 (4.18-6.76) | 5.17E-42 |  |
| Marital status |  |  |  |  |  |  |  |  |
| Married/Living with partner | 1.00 (Reference) | 2.54 (2.08-3.11) | 1.22E-19 | 2.01 (1.28-3.16) | 2.34E-03 | 4.37 (3.15-6.05) | 9.15E-19 | 8.34E-01 |
| Others | 1.00 (Reference) | 2.25 (1.75-2.88) | 1.59E-10 | 1.83 (1.10-3.04) | 2.03E-02 | 4.96 (3.54-6.95) | 1.61E-20 |  |
| Educational status |  |  |  |  |  |  |  |  |
| Less than high school | 1.00 (Reference) | 2.68 (2.13-3.37) | 4.12E-17 | 2.72 (1.70-4.35) | 3.16E-05 | 5.00 (3.62-6.92) | 2.16E-22 | 4.41E-02 |
| High school | 1.00 (Reference) | 2.49 (1.82-3.41) | 1.31E-08 | 1.88 (0.86-4.13) | 1.15E-01 | 6.60 (3.94-11.06) | 6.94E-13 |  |
| College or above | 1.00 (Reference) | 2.35 (1.79-3.09) | 9.45E-10 | 1.92 (1.05-3.50) | 3.34E-02 | 5.15 (3.33-7.96) | 1.62E-13 |  |
| **C.** FI-Combined | | | | | | | | |
| **Subgroups** | **No Heart Stress or Frailty** | **Heart Stress, no Frailty** | | **Frailty, no Heart Stress** | | **Frailty, and Heart Stress** | |  |
|  |  | **HR (95% CI)** | **P-value** | **HR (95% CI)** | **P-value** | **HR (95% CI)** | **P-value** | **P-int** |
| Age group |  |  |  |  |  |  |  |  |
| 20-64 | 1.00 (Reference) | 3.68 (2.62-5.17) | 6.17E-14 | 2.01 (1.17-3.45) | 1.18E-02 | 11.68 (7.41-18.41) | 3.62E-26 | 8.62E-01 |
| 65 years or older | 1.00 (Reference) | 2.11 (1.76-2.53) | 1.44E-15 | 3.13 (2.04-4.80) | 1.70E-07 | 6.20 (4.73-8.12) | 5.55E-40 |  |
| Gender |  |  |  |  |  |  |  |  |
| Male | 1.00 (Reference) | 2.72 (2.19-3.36) | 4.56E-20 | 2.73 (1.79-4.16) | 3.28E-06 | 8.65 (6.37-11.73) | 1.41E-43 | 7.77E-01 |
| Female | 1.00 (Reference) | 2.21 (1.72-2.84) | 7.19E-10 | 2.81 (1.62-4.88) | 2.27E-04 | 6.60 (4.71-9.24) | 5.05E-28 |  |
| Body mass index |  |  |  |  |  |  |  |  |
| Normal or underweight | 1.00 (Reference) | 2.29 (1.65-3.18) | 6.37E-07 | 4.06 (1.63-10.09) | 2.59E-03 | 7.19 (4.46-11.59) | 6.06E-16 | 1.04E-01 |
| Overweight | 1.00 (Reference) | 2.15 (1.65-2.80) | 1.33E-08 | 4.40 (2.46-7.89) | 6.25E-07 | 5.87 (3.84-8.99) | 3.45E-16 |  |
| Obese | 1.00 (Reference) | 2.61 (1.97-3.46) | 1.93E-11 | 1.56 (0.90-2.69) | 1.14E-01 | 6.27 (4.30-9.15) | 1.54E-21 |  |
| Race/Ethnicity |  |  |  |  |  |  |  |  |
| White/Caucasian | 1.00 (Reference) | 2.25 (1.82-2.77) | 3.21E-14 | 2.38 (1.48-3.81) | 3.30E-04 | 8.32 (6.01-11.51) | 1.84E-37 | 1.08E-01 |
| Black/African American | 1.00 (Reference) | 2.89 (1.95-4.30) | 1.51E-07 | 1.84 (0.82-4.13) | 1.37E-01 | 7.97 (5.05-12.58) | 4.97E-19 |  |
| Others | 1.00 (Reference) | 2.89 (2.05-4.09) | 1.61E-09 | 3.66 (1.93-6.94) | 6.95E-05 | 5.61 (3.52-8.96) | 4.81E-13 |  |
| Smoking status |  |  |  |  |  |  |  |  |
| Ever | 1.00 (Reference) | 2.48 (2.00-3.08) | 1.80E-16 | 2.85 (1.88-4.33) | 7.84E-07 | 8.49 (6.29-11.44) | 1.22E-44 | 2.90E-01 |
| Never | 1.00 (Reference) | 2.39 (1.87-3.05) | 3.57E-12 | 2.66 (1.48-4.78) | 1.02E-03 | 6.84 (4.88-9.60) | 9.56E-29 |  |
| Vigorous physical activity |  |  |  |  |  |  |  |  |
| Yes | 1.00 (Reference) | 2.83 (1.81-4.40) | 4.37E-06 | 0.47 (0.06-3.66) | 4.71E-01 | 21.28 (5.51-82.23) | 9.23E-06 | 5.49E-02 |
| No | 1.00 (Reference) | 2.37 (1.99-2.82) | 2.47E-22 | 3.12 (2.22-4.38) | 5.48E-11 | 7.53 (5.99-9.47) | 1.02E-66 |  |
| Marital status |  |  |  |  |  |  |  |  |
| Married/Living with partner | 1.00 (Reference) | 2.34 (1.90-2.88) | 1.08E-15 | 2.27 (1.36-3.79) | 1.78E-03 | 6.34 (4.61-8.72) | 6.25E-30 | 9.44E-01 |
| Others | 1.00 (Reference) | 2.17 (1.67-2.83) | 1.02E-08 | 2.54 (1.54-4.18) | 2.50E-04 | 7.52 (5.39-10.49) | 1.41E-32 |  |
| Educational status |  |  |  |  |  |  |  |  |
| Less than high school | 1.00 (Reference) | 2.61 (2.03-3.34) | 3.94E-14 | 3.37 (2.21-5.13) | 1.59E-08 | 6.30 (4.61-8.59) | 3.89E-31 | 1.68E-02 |
| High school | 1.00 (Reference) | 2.14 (1.54-2.98) | 5.38E-06 | 1.49 (0.53-4.23) | 4.51E-01 | 12.50 (7.63-20.46) | 1.01E-23 |  |
| College or above | 1.00 (Reference) | 2.33 (1.75-3.11) | 7.24E-09 | 2.04 (0.95-4.38) | 6.68E-02 | 6.84 (4.32-10.82) | 2.25E-16 |  |

| **eTable 17.** Association of frailty and heart stress re-defined by a fixed cutoff with risk of cancer mortality in subgroups in NHANES. | | | | | | | | |
| --- | --- | --- | --- | --- | --- | --- | --- | --- |
| **A.** FI-Self-report | | | | | | | | |
| **Subgroups** | **No Heart Stress or Frailty** | **Heart Stress, no Frailty** | | **Frailty, no Heart Stress** | | **Frailty, and Heart Stress** | |  |
|  |  | **HR (95% CI)** | **P-value** | **HR (95% CI)** | **P-value** | **HR (95% CI)** | **P-value** | **P-int** |
| Age group |  |  |  |  |  |  |  |  |
| 20-64 | 1.00 (Reference) | 2.07 (1.38-3.10) | 4.61E-04 | 2.63 (1.74-3.98) | 4.06E-06 | 4.69 (2.75-7.99) | 1.33E-08 | 5.95E-01 |
| 65 years or older | 1.00 (Reference) | 1.46 (1.16-1.83) | 1.19E-03 | 1.59 (1.09-2.31) | 1.60E-02 | 2.57 (1.80-3.69) | 2.42E-07 |  |
| Gender |  |  |  |  |  |  |  |  |
| Male | 1.00 (Reference) | 1.84 (1.43-2.38) | 3.06E-06 | 2.13 (1.46-3.11) | 8.21E-05 | 3.03 (1.95-4.71) | 7.59E-07 | 8.84E-01 |
| Female | 1.00 (Reference) | 1.37 (1.00-1.88) | 4.73E-02 | 2.20 (1.46-3.31) | 1.49E-04 | 3.36 (2.28-4.93) | 7.11E-10 |  |
| Body mass index |  |  |  |  |  |  |  |  |
| Normal or underweight | 1.00 (Reference) | 1.78 (1.20-2.65) | 4.31E-03 | 2.44 (1.27-4.67) | 7.31E-03 | 4.44 (2.60-7.60) | 5.21E-08 | 6.51E-01 |
| Overweight | 1.00 (Reference) | 1.28 (0.94-1.74) | 1.21E-01 | 1.25 (0.74-2.11) | 3.95E-01 | 2.75 (1.66-4.56) | 8.95E-05 |  |
| Obese | 1.00 (Reference) | 2.08 (1.43-3.01) | 1.15E-04 | 2.24 (1.45-3.45) | 2.69E-04 | 2.44 (1.43-4.17) | 1.12E-03 |  |
| Race/Ethnicity |  |  |  |  |  |  |  |  |
| White/Caucasian | 1.00 (Reference) | 1.68 (1.30-2.16) | 7.25E-05 | 2.36 (1.57-3.54) | 3.32E-05 | 4.75 (3.27-6.90) | 3.14E-16 | 4.01E-04 |
| Black/African American | 1.00 (Reference) | 1.99 (1.18-3.37) | 1.03E-02 | 2.18 (1.23-3.86) | 7.40E-03 | 2.73 (1.18-6.34) | 1.90E-02 |  |
| Others | 1.00 (Reference) | 1.38 (0.90-2.10) | 1.35E-01 | 1.66 (0.99-2.80) | 5.57E-02 | 2.15 (1.24-3.71) | 6.19E-03 |  |
| Smoking status |  |  |  |  |  |  |  |  |
| Ever | 1.00 (Reference) | 1.56 (1.22-1.98) | 3.85E-04 | 1.92 (1.39-2.65) | 8.12E-05 | 2.66 (1.86-3.81) | 8.10E-08 | 1.63E-09 |
| Never | 1.00 (Reference) | 1.90 (1.34-2.68) | 2.74E-04 | 2.22 (1.31-3.76) | 3.10E-03 | 5.15 (3.18-8.35) | 3.01E-11 |  |
| Vigorous physical activity |  |  |  |  |  |  |  |  |
| Yes | 1.00 (Reference) | 1.38 (0.84-2.26) | 2.03E-01 | 1.66 (0.64-4.29) | 2.99E-01 | 0.00 (0.00-Inf) | 9.95E-01 | 1.83E-02 |
| No | 1.00 (Reference) | 1.71 (1.38-2.13) | 1.49E-06 | 2.10 (1.57-2.80) | 5.33E-07 | 3.46 (2.59-4.63) | 5.66E-17 |  |
| Marital status |  |  |  |  |  |  |  |  |
| Married/Living with partner | 1.00 (Reference) | 1.83 (1.43-2.35) | 1.83E-06 | 1.93 (1.33-2.78) | 4.58E-04 | 2.93 (1.94-4.43) | 3.71E-07 | 9.84E-02 |
| Others | 1.00 (Reference) | 1.24 (0.88-1.74) | 2.25E-01 | 1.90 (1.22-2.97) | 4.72E-03 | 3.35 (2.21-5.10) | 1.48E-08 |  |
| Educational status |  |  |  |  |  |  |  |  |
| Less than high school | 1.00 (Reference) | 1.41 (1.01-1.96) | 4.20E-02 | 1.70 (1.15-2.51) | 7.80E-03 | 2.66 (1.76-4.00) | 2.99E-06 | 1.77E-01 |
| High school | 1.00 (Reference) | 1.87 (1.25-2.81) | 2.38E-03 | 1.68 (0.91-3.12) | 9.74E-02 | 5.81 (3.36-10.05) | 3.13E-10 |  |
| College or above | 1.00 (Reference) | 1.83 (1.32-2.53) | 2.74E-04 | 2.98 (1.78-4.97) | 3.14E-05 | 2.57 (1.43-4.64) | 1.65E-03 |  |
| **B.** FI-Lab | | | | | | | | |
| **Subgroups** | **No Heart Stress or Frailty** | **Heart Stress, no Frailty** | | **Frailty, no Heart Stress** | | **Frailty, and Heart Stress** | |  |
|  |  | **HR (95% CI)** | **P-value** | **HR (95% CI)** | **P-value** | **HR (95% CI)** | **P-value** | **P-int** |
| Age group |  |  |  |  |  |  |  |  |
| 20-64 | 1.00 (Reference) | 1.81 (1.32-2.47) | 2.13E-04 | 1.69 (1.09-2.64) | 2.00E-02 | 5.18 (2.99-8.96) | 4.13E-09 | 3.30E-01 |
| 65 years or older | 1.00 (Reference) | 1.39 (1.12-1.73) | 3.14E-03 | 2.57 (1.62-4.06) | 5.90E-05 | 3.23 (2.24-4.66) | 3.74E-10 |  |
| Gender |  |  |  |  |  |  |  |  |
| Male | 1.00 (Reference) | 1.69 (1.32-2.16) | 3.67E-05 | 2.34 (1.60-3.42) | 1.33E-05 | 3.46 (2.45-4.87) | 1.52E-12 | 8.98E-01 |
| Female | 1.00 (Reference) | 1.44 (1.11-1.86) | 5.42E-03 | 1.63 (0.92-2.91) | 9.65E-02 | 5.96 (3.33-10.66) | 1.79E-09 |  |
| Body mass index |  |  |  |  |  |  |  |  |
| Normal or underweight | 1.00 (Reference) | 1.53 (1.07-2.20) | 2.06E-02 | 2.05 (1.06-3.98) | 3.27E-02 | 3.03 (1.81-5.06) | 2.33E-05 | 4.53E-02 |
| Overweight | 1.00 (Reference) | 1.35 (1.01-1.81) | 4.33E-02 | 2.74 (1.65-4.56) | 9.72E-05 | 3.72 (2.21-6.26) | 7.47E-07 |  |
| Obese | 1.00 (Reference) | 1.90 (1.39-2.60) | 5.57E-05 | 1.61 (0.95-2.72) | 7.40E-02 | 5.06 (2.76-9.28) | 1.67E-07 |  |
| Race/Ethnicity |  |  |  |  |  |  |  |  |
| White/Caucasian | 1.00 (Reference) | 1.63 (1.30-2.05) | 2.34E-05 | 1.97 (1.20-3.24) | 7.48E-03 | 4.08 (2.73-6.09) | 5.88E-12 | 5.45E-02 |
| Black/African American | 1.00 (Reference) | 2.04 (1.26-3.28) | 3.50E-03 | 1.46 (0.75-2.85) | 2.62E-01 | 3.80 (2.07-6.98) | 1.75E-05 |  |
| Others | 1.00 (Reference) | 1.23 (0.85-1.80) | 2.74E-01 | 2.64 (1.55-4.51) | 3.77E-04 | 3.07 (1.65-5.71) | 4.16E-04 |  |
| Smoking status |  |  |  |  |  |  |  |  |
| Ever | 1.00 (Reference) | 1.46 (1.17-1.82) | 6.82E-04 | 1.96 (1.34-2.87) | 4.91E-04 | 3.17 (2.20-4.57) | 5.17E-10 | 2.93E-05 |
| Never | 1.00 (Reference) | 1.86 (1.37-2.53) | 6.83E-05 | 2.14 (1.20-3.82) | 1.02E-02 | 7.41 (4.55-12.05) | 7.63E-16 |  |
| Vigorous physical activity |  |  |  |  |  |  |  |  |
| Yes | 1.00 (Reference) | 1.20 (0.75-1.92) | 4.51E-01 | 1.33 (0.57-3.06) | 5.08E-01 | 1.89 (0.72-4.96) | 1.95E-01 | 8.61E-01 |
| No | 1.00 (Reference) | 1.60 (1.32-1.94) | 1.86E-06 | 2.18 (1.54-3.09) | 1.05E-05 | 4.25 (3.12-5.79) | 5.32E-20 |  |
| Marital status |  |  |  |  |  |  |  |  |
| Married/Living with partner | 1.00 (Reference) | 1.65 (1.32-2.07) | 1.23E-05 | 1.88 (1.22-2.89) | 4.39E-03 | 3.52 (2.31-5.39) | 5.92E-09 | 4.04E-01 |
| Others | 1.00 (Reference) | 1.37 (1.02-1.86) | 3.86E-02 | 1.86 (1.15-3.03) | 1.21E-02 | 3.66 (2.37-5.65) | 4.44E-09 |  |
| Educational status |  |  |  |  |  |  |  |  |
| Less than high school | 1.00 (Reference) | 1.39 (1.04-1.85) | 2.46E-02 | 2.47 (1.58-3.86) | 7.52E-05 | 3.48 (2.19-5.53) | 1.28E-07 | 1.07E-02 |
| High school | 1.00 (Reference) | 1.89 (1.32-2.71) | 5.54E-04 | 2.01 (0.97-4.20) | 6.19E-02 | 4.24 (2.31-7.80) | 3.18E-06 |  |
| College or above | 1.00 (Reference) | 1.54 (1.15-2.08) | 4.14E-03 | 1.58 (0.87-2.86) | 1.30E-01 | 4.59 (2.81-7.48) | 9.92E-10 |  |
| **C.** FI-Combined | | | | | | | |  |
| **Subgroups** | **No Heart Stress or Frailty** | **Heart Stress, no Frailty** | | **Frailty, no Heart Stress** | | **Frailty, and Heart Stress** | |  |
|  |  | **HR (95% CI)** | **P-value** | **HR (95% CI)** | **P-value** | **HR (95% CI)** | **P-value** |  |
| Age group |  |  |  |  |  |  |  |  |
| 20-64 | 1.00 (Reference) | 1.92 (1.32-2.80) | 7.07E-04 | 2.63 (1.60-4.32) | 1.39E-04 | 5.04 (2.82-8.99) | 4.49E-08 | 5.84E-01 |
| 65 years or older | 1.00 (Reference) | 1.44 (1.16-1.80) | 1.11E-03 | 2.55 (1.67-3.91) | 1.60E-05 | 3.24 (2.20-4.75) | 2.25E-09 |  |
| Gender |  |  |  |  |  |  |  |  |
| Male | 1.00 (Reference) | 1.72 (1.34-2.22) | 2.83E-05 | 2.92 (1.91-4.46) | 8.30E-07 | 5.33 (3.50-8.11) | 5.91E-15 | 1.79E-01 |
| Female | 1.00 (Reference) | 1.46 (1.10-1.94) | 8.69E-03 | 2.59 (1.59-4.23) | 1.41E-04 | 3.14 (1.97-4.99) | 1.46E-06 |  |
| Body mass index |  |  |  |  |  |  |  |  |
| Normal or underweight | 1.00 (Reference) | 1.87 (1.28-2.73) | 1.29E-03 | 4.85 (2.31-10.20) | 3.09E-05 | 4.78 (2.61-8.75) | 3.97E-07 | 1.45E-01 |
| Overweight | 1.00 (Reference) | 1.25 (0.92-1.68) | 1.52E-01 | 1.92 (0.96-3.85) | 6.48E-02 | 3.95 (2.32-6.72) | 3.93E-07 |  |
| Obese | 1.00 (Reference) | 2.00 (1.41-2.84) | 1.04E-04 | 2.41 (1.49-3.90) | 3.44E-04 | 3.39 (1.86-6.18) | 6.55E-05 |  |
| Race/Ethnicity |  |  |  |  |  |  |  |  |
| White/Caucasian | 1.00 (Reference) | 1.69 (1.33-2.16) | 2.26E-05 | 2.65 (1.61-4.35) | 1.24E-04 | 7.76 (5.14-11.71) | 1.70E-22 | 2.39E-04 |
| Black/African American | 1.00 (Reference) | 1.82 (1.09-3.03) | 2.21E-02 | 2.85 (1.49-5.45) | 1.50E-03 | 3.93 (1.82-8.51) | 4.99E-04 |  |
| Others | 1.00 (Reference) | 1.37 (0.92-2.05) | 1.26E-01 | 2.69 (1.48-4.89) | 1.19E-03 | 2.38 (1.32-4.30) | 4.08E-03 |  |
| Smoking status |  |  |  |  |  |  |  |  |
| Ever | 1.00 (Reference) | 1.55 (1.23-1.95) | 2.29E-04 | 2.83 (1.93-4.15) | 9.96E-08 | 3.70 (2.46-5.56) | 2.83E-10 | 6.38E-10 |
| Never | 1.00 (Reference) | 1.83 (1.31-2.54) | 3.48E-04 | 2.63 (1.43-4.84) | 1.94E-03 | 5.92 (3.67-9.55) | 3.37E-13 |  |
| Vigorous physical activity |  |  |  |  |  |  |  |  |
| Yes | 1.00 (Reference) | 1.20 (0.73-1.97) | 4.82E-01 | 1.85 (0.55-6.21) | 3.20E-01 | 4.96 (0.46-52.87) | 1.85E-01 | 5.31E-02 |
| No | 1.00 (Reference) | 1.71 (1.39-2.11) | 3.90E-07 | 2.74 (1.95-3.83) | 4.72E-09 | 4.40 (3.21-6.02) | 2.18E-20 |  |
| Marital status |  |  |  |  |  |  |  |  |
| Married/Living with partner | 1.00 (Reference) | 1.75 (1.38-2.23) | 4.76E-06 | 2.47 (1.60-3.83) | 5.08E-05 | 4.34 (2.74-6.87) | 3.63E-10 | 5.87E-01 |
| Others | 1.00 (Reference) | 1.35 (0.98-1.86) | 6.42E-02 | 2.38 (1.44-3.91) | 6.52E-04 | 3.54 (2.28-5.49) | 1.79E-08 |  |
| Educational status |  |  |  |  |  |  |  |  |
| Less than high school | 1.00 (Reference) | 1.40 (1.03-1.90) | 3.14E-02 | 2.21 (1.39-3.52) | 8.72E-04 | 3.06 (1.97-4.75) | 6.11E-07 | 1.47E-01 |
| High school | 1.00 (Reference) | 1.98 (1.35-2.90) | 4.62E-04 | 2.89 (1.44-5.82) | 2.91E-03 | 9.76 (4.67-20.40) | 1.35E-09 |  |
| College or above | 1.00 (Reference) | 1.69 (1.23-2.33) | 1.28E-03 | 3.61 (1.99-6.54) | 2.42E-05 | 5.42 (3.03-9.67) | 1.14E-08 |  |

| **eTable 18.** Association of frailty and heart stress with risk of all-cause mortality in NHANES and HRS after excluding individuals who died within 2 years post-baseline. | | | | | | | | |
| --- | --- | --- | --- | --- | --- | --- | --- | --- |
| **A.** FI-Self-report | | | | | | | | |
|  |  |  | **Model 1** | | **Model 2** | | **Model 3** | |
|  |  | **Event/N (%)** | **HR (95%CI)** | **P-value** | **HR 95%CI** | **P-value** | **HR (95%CI)** | **P-value** |
| **NHANES** |  |  |  |  |  |  |  |  |
| No Frailty | No Heart Stress | 1263/3081 (40.99) | Ref |  | Ref |  | Ref |  |
|  | Heart Stress | 848/1275 (66.51) | 2.21 (2.03-2.41) | 5.82E-71 | 1.83 (1.68-2.00) | 1.09E-40 | 1.83 (1.67-2.00) | 2.16E-40 |
| Frailty | No Heart Stress | 369/650 (56.77) | 1.69 (1.50-1.89) | 1.05E-18 | 1.96 (1.74-2.21) | 2.18E-29 | 2.00 (1.78-2.26) | 2.92E-30 |
|  | Heart Stress | 382/468 (81.62) | 3.91 (3.48-4.39) | 6.72E-118 | 3.28 (2.90-3.70) | 5.09E-82 | 3.29 (2.91-3.71) | 1.96E-81 |
| **HRS** |  |  |  |  |  |  |  |  |
| No Frailty | No Heart Stress | 421/5485 (7.68) | Ref |  | Ref |  | Ref |  |
|  | Heart Stress | 383/1818 (21.07) | 2.92 (2.55-3.36) | 4.30E-52 | 2.03 (1.76-2.34) | 1.77E-22 | 2.00 (1.74-2.31) | 1.02E-21 |
| Frailty | No Heart Stress | 209/1043 (20.04) | 2.80 (2.37-3.31) | 4.71E-34 | 2.53 (2.14-2.99) | 2.70E-27 | 2.33 (1.95-2.78) | 4.92E-21 |
|  | Heart Stress | 313/769 (40.70) | 6.61 (5.71-7.65) | 9.69E-141 | 4.48 (3.83-5.22) | 3.49E-80 | 4.22 (3.60-4.94) | 7.56E-71 |
| **B.** FI-Lab | | | | | | | | |
|  |  |  | **Model 1** | | **Model 2** | | **Model 3** | |
|  |  | **Event/N (%)** | **HR (95%CI)** | **P-value** | **HR 95%CI** | **P-value** | **HR (95%CI)** | **P-value** |
| **NHANES** |  |  |  |  |  |  |  |  |
| No Frailty | No Heart Stress | 1748/8542 (20.46) | Ref |  | Ref |  | Ref |  |
|  | Heart Stress | 1014/2255 (44.97) | 2.72 (2.52-2.94) | 2.28E-141 | 1.76 (1.62-1.91) | 2.76E-43 | 1.76 (1.63-1.91) | 2.32E-43 |
| Frailty | No Heart Stress | 211/746 (28.28) | 1.44 (1.25-1.66) | 6.37E-07 | 1.82 (1.58-2.10) | 2.35E-16 | 1.82 (1.57-2.10) | 4.68E-16 |
|  | Heart Stress | 285/448 (63.62) | 4.92 (4.34-5.58) | 1.16E-136 | 3.01 (2.64-3.42) | 2.17E-62 | 3.05 (2.67-3.47) | 2.87E-63 |
| **HRS** |  |  |  |  |  |  |  |  |
| No Frailty | No Heart Stress | 454/5656 (8.03) | Ref |  | Ref |  | Ref |  |
|  | Heart Stress | 372/1831 (20.32) | 2.68 (2.33-3.07) | 5.50E-45 | 1.80 (1.56-2.07) | 3.46E-16 | 1.80 (1.56-2.07) | 4.81E-16 |
| Frailty | No Heart Stress | 159/705 (22.55) | 3.04 (2.54-3.64) | 1.74E-33 | 2.35 (1.96-2.83) | 8.10E-20 | 2.22 (1.84-2.67) | 3.55E-17 |
|  | Heart Stress | 297/695 (42.73) | 6.69 (5.78-7.75) | 1.19E-142 | 4.39 (3.77-5.11) | 2.04E-80 | 4.20 (3.60-4.90) | 4.80E-75 |
| **C.** FI-Combined | | | | | | | | |
|  |  |  | **Model 1** | | **Model 2** | | **Model 3** | |
|  |  | **Event/N (%)** | **HR (95%CI)** | **P-value** | **HR 95%CI** | **P-value** | **HR (95%CI)** | **P-value** |
| **NHANES** |  |  |  |  |  |  |  |  |
| No Frailty | No Heart Stress | 1387/3340 (41.53) | Ref |  | Ref |  | Ref |  |
|  | Heart Stress | 902/1365 (66.08) | 2.16 (1.98-2.35) | 1.33E-71 | 1.77 (1.63-1.93) | 5.02E-39 | 1.77 (1.63-1.93) | 7.84E-39 |
| Frailty | No Heart Stress | 244/383 (63.71) | 2.04 (1.78-2.34) | 7.45E-25 | 2.40 (2.10-2.76) | 4.92E-36 | 2.48 (2.15-2.85) | 5.42E-37 |
|  | Heart Stress | 332/384 (86.46) | 4.63 (4.10-5.23) | 3.26E-135 | 3.68 (3.24-4.17) | 3.68E-91 | 3.71 (3.27-4.22) | 7.58E-91 |
| **HRS** |  |  |  |  |  |  |  |  |
| No Frailty | No Heart Stress | 428/5601 (7.64) | Ref |  | Ref |  | Ref |  |
|  | Heart Stress | 361/1812 (19.92) | 2.75 (2.39-3.16) | 1.72E-45 | 1.89 (1.64-2.18) | 3.47E-18 | 1.87 (1.62-2.16) | 1.10E-17 |
| Frailty | No Heart Stress | 195/826 (23.61) | 3.36 (2.84-3.98) | 1.13E-44 | 2.87 (2.42-3.40) | 1.32E-33 | 2.65 (2.22-3.17) | 1.82E-26 |
|  | Heart Stress | 318/739 (43.03) | 7.16 (6.19-8.28) | 4.16E-155 | 4.92 (4.23-5.74) | 3.35E-93 | 4.71 (4.03-5.51) | 3.83E-84 |

| **eTable 19.** Association of frailty and heart stress with risk of cause-specific mortality in NHANES after excluding individuals who died within 2 years post-baseline. | | | | | | | | |
| --- | --- | --- | --- | --- | --- | --- | --- | --- |
| **A.** FI-Self-report | | | | | | | | |
|  |  |  | **Model 1** | | **Model 2** | | **Model 3** | |
|  |  | **Event/N (%)** | **HR (95%CI)** | **P-value** | **HR 95%CI** | **P-value** | **HR (95%CI)** | **P-value** |
| **Heart disease** |  |  |  |  |  |  |  |  |
| No Frailty | No Heart Stress | 268/2086 (12.85) | Ref |  | Ref |  | Ref |  |
|  | Heart Stress | 289/716 (40.36) | 3.93 (3.33-4.65) | 1.81E-58 | 2.89 (2.44-3.43) | 4.03E-34 | 2.90 (2.44-3.44) | 4.36E-34 |
| Frailty | No Heart Stress | 96/377 (25.46) | 2.23 (1.76-2.81) | 1.73E-11 | 2.86 (2.26-3.62) | 2.79E-18 | 2.67 (2.09-3.42) | 3.64E-15 |
|  | Heart Stress | 130/216 (60.19) | 7.89 (6.39-9.74) | 4.49E-82 | 6.52 (5.17-8.23) | 4.79E-56 | 6.50 (5.15-8.21) | 7.99E-56 |
| **Cerebrovascular disease** | |  |  |  |  |  |  |  |
| No Frailty | No Heart Stress | 72/1890 (3.81) |  |  |  |  |  |  |
|  | Heart Stress | 42/469 (8.96) | 2.45 (1.68-3.59) | 3.90E-06 | 1.94 (1.32-2.84) | 7.37E-04 | 1.86 (1.26-2.74) | 1.74E-03 |
| Frailty | No Heart Stress | 23/304 (7.57) | 2.05 (1.28-3.28) | 2.69E-03 | 2.49 (1.55-4.00) | 1.53E-04 | 2.24 (1.37-3.68) | 1.38E-03 |
|  | Heart Stress | 23/109 (21.10) | 6.30 (3.94-10.07) | 1.61E-14 | 3.97 (2.34-6.74) | 3.20E-07 | 3.50 (2.04-6.01) | 5.66E-06 |
| **CVD** |  |  |  |  |  |  |  |  |
| No Frailty | No Heart Stress | 340/2158 (15.76) |  |  |  |  |  |  |
|  | Heart Stress | 331/758 (43.67) | 3.51 (3.01-4.08) | 3.66E-59 | 2.57 (2.20-3.00) | 1.30E-32 | 2.57 (2.20-3.01) | 1.45E-32 |
| Frailty | No Heart Stress | 119/400 (29.75) | 2.15 (1.74-2.64) | 7.66E-13 | 2.64 (2.14-3.26) | 1.89E-19 | 2.62 (2.11-3.26) | 2.46E-18 |
|  | Heart Stress | 153/239 (64.02) | 7.00 (5.77-8.48) | 1.42E-87 | 5.48 (4.44-6.77) | 3.51E-56 | 5.44 (4.40-6.72) | 2.18E-55 |
| **Cancer** |  |  |  |  |  |  |  |  |
| No Frailty | No Heart Stress | 290/2108 (13.76) |  |  |  |  |  |  |
|  | Heart Stress | 137/564 (24.29) | 1.93 (1.58-2.37) | 1.95E-10 | 1.78 (1.45-2.18) | 3.57E-08 | 1.73 (1.41-2.13) | 1.82E-07 |
| Frailty | No Heart Stress | 83/364 (22.80) | 1.75 (1.37-2.24) | 6.32E-06 | 2.10 (1.64-2.69) | 3.46E-09 | 2.18 (1.69-2.80) | 1.32E-09 |
|  | Heart Stress | 49/135 (36.30) | 3.32 (2.45-4.50) | 8.11E-15 | 3.31 (2.40-4.55) | 1.91E-13 | 3.12 (2.26-4.30) | 4.23E-12 |
| **Others** | |  |  |  |  |  |  |  |
| No Frailty | No Heart Stress | 633/2451 (25.83) |  |  |  |  |  |  |
|  | Heart Stress | 380/807 (47.09) | 2.23 (1.96-2.53) | 4.98E-35 | 1.81 (1.59-2.05) | 3.22E-19 | 1.79 (1.57-2.04) | 1.31E-18 |
| Frailty | No Heart Stress | 167/448 (37.28) | 1.66 (1.40-1.96) | 7.02E-09 | 1.98 (1.67-2.36) | 6.25E-15 | 1.96 (1.64-2.34) | 7.72E-14 |
|  | Heart Stress | 180/266 (67.67) | 4.34 (3.68-5.13) | 6.05E-67 | 3.31 (2.78-3.94) | 6.73E-41 | 3.12 (2.61-3.73) | 8.78E-36 |
| **B.** FI-Lab | | | | | | | | |
|  |  |  | **Model 1** | | **Model 2** | | **Model 3** | |
|  |  | **Event/N (%)** | **HR (95%CI)** | **P-value** | **HR 95%CI** | **P-value** | **HR (95%CI)** | **P-value** |
| **Heart disease** |  |  |  |  |  |  |  |  |
| No Frailty | No Heart Stress | 379/7173 (5.28) |  |  |  |  |  |  |
|  | Heart Stress | 342/1583 (21.60) | 4.59 (3.97-5.32) | 9.23E-93 | 2.65 (2.28-3.09) | 5.07E-36 | 2.67 (2.29-3.11) | 4.06E-36 |
| Frailty | No Heart Stress | 37/572 (6.47) | 1.19 (0.85-1.67) | 3.12E-01 | 1.79 (1.27-2.50) | 7.83E-04 | 1.86 (1.33-2.62) | 3.35E-04 |
|  | Heart Stress | 90/253 (35.57) | 8.45 (6.71-10.63) | 7.14E-74 | 4.41 (3.45-5.64) | 1.68E-32 | 4.55 (3.56-5.81) | 6.72E-34 |
| **Cerebrovascular disease** | |  |  |  |  |  |  |  |
| No Frailty | No Heart Stress | 104/6898 (1.51) |  |  |  |  |  |  |
|  | Heart Stress | 53/1294 (4.10) | 2.78 (2.00-3.87) | 1.40E-09 | 1.88 (1.34-2.63) | 2.34E-04 | 1.89 (1.35-2.65) | 2.08E-04 |
| Frailty | No Heart Stress | 8/543 (1.47) | 0.94 (0.46-1.93) | 8.72E-01 | 1.90 (0.92-3.91) | 8.12E-02 | 1.68 (0.78-3.62) | 1.87E-01 |
|  | Heart Stress | 16/179 (8.94) | 6.02 (3.56-10.19) | 2.34E-11 | 3.00 (1.72-5.25) | 1.15E-04 | 3.28 (1.89-5.69) | 2.48E-05 |
| **CVD** |  |  |  |  |  |  |  |  |
| No Frailty | No Heart Stress | 483/7277 (6.64) |  |  |  |  |  |  |
|  | Heart Stress | 395/1636 (24.14) | 4.13 (3.61-4.72) | 7.16E-97 | 2.38 (2.07-2.74) | 1.27E-34 | 2.39 (2.08-2.74) | 1.24E-34 |
| Frailty | No Heart Stress | 45/580 (7.76) | 1.14 (0.84-1.54) | 4.16E-01 | 1.70 (1.25-2.31) | 6.76E-04 | 1.67 (1.23-2.28) | 1.15E-03 |
|  | Heart Stress | 106/269 (39.41) | 7.65 (6.20-9.45) | 4.06E-80 | 3.84 (3.07-4.80) | 2.21E-32 | 3.93 (3.15-4.91) | 1.46E-33 |
| **Cancer** |  |  |  |  |  |  |  |  |
| No Frailty | No Heart Stress | 430/7224 (5.95) |  |  |  |  |  |  |
|  | Heart Stress | 168/1409 (11.92) | 2.11 (1.76-2.52) | 2.57E-16 | 1.68 (1.40-2.02) | 2.31E-08 | 1.68 (1.40-2.02) | 3.11E-08 |
| Frailty | No Heart Stress | 52/587 (8.86) | 1.49 (1.12-1.99) | 6.64E-03 | 2.25 (1.68-3.00) | 4.06E-08 | 2.24 (1.67-2.99) | 5.45E-08 |
|  | Heart Stress | 32/195 (16.41) | 2.95 (2.06-4.23) | 3.54E-09 | 2.56 (1.77-3.70) | 5.42E-07 | 2.63 (1.81-3.83) | 4.13E-07 |
| **Others** | |  |  |  |  |  |  |  |
| No Frailty | No Heart Stress | 835/7629 (10.95) |  |  |  |  |  |  |
|  | Heart Stress | 451/1692 (26.65) | 2.74 (2.44-3.07) | 1.62E-66 | 1.79 (1.59-2.02) | 3.95E-22 | 1.78 (1.58-2.01) | 1.09E-21 |
| Frailty | No Heart Stress | 114/649 (17.57) | 1.64 (1.35-1.99) | 7.97E-07 | 2.28 (1.87-2.77) | 2.17E-16 | 2.22 (1.82-2.70) | 2.18E-15 |
|  | Heart Stress | 147/310 (47.42) | 5.88 (4.93-7.00) | 5.07E-87 | 3.41 (2.84-4.10) | 1.26E-39 | 3.46 (2.88-4.16) | 5.06E-40 |
| **C.** FI-Combined | | | | | | | | |
|  |  |  | **Model 1** | | **Model 2** | | **Model 3** | |
|  |  | **Event/N (%)** | **HR (95%CI)** | **P-value** | **HR 95%CI** | **P-value** | **HR (95%CI)** | **P-value** |
| **Heart disease** |  |  |  |  |  |  |  |  |
| No Frailty | No Heart Stress | 312/2265 (13.77) |  |  |  |  |  |  |
|  | Heart Stress | 311/774 (40.18) | 3.66 (3.12-4.28) | 1.22E-58 | 2.70 (2.30-3.18) | 2.25E-33 | 2.72 (2.31-3.20) | 1.54E-33 |
| Frailty | No Heart Stress | 54/193 (27.98) | 2.35 (1.76-3.14) | 6.82E-09 | 3.05 (2.28-4.08) | 5.74E-14 | 2.88 (2.13-3.91) | 9.39E-12 |
|  | Heart Stress | 109/161 (67.70) | 9.04 (7.25-11.26) | 7.17E-86 | 6.42 (5.04-8.16) | 1.36E-51 | 6.52 (5.12-8.31) | 6.36E-52 |
| **Cerebrovascular disease** | |  |  |  |  |  |  |  |
| No Frailty | No Heart Stress | 82/2035 (4.03) |  |  |  |  |  |  |
|  | Heart Stress | 45/508 (8.86) | 2.29 (1.59-3.30) | 7.76E-06 | 1.79 (1.24-2.58) | 1.96E-03 | 1.70 (1.17-2.47) | 5.16E-03 |
| Frailty | No Heart Stress | 13/152 (8.55) | 2.18 (1.21-3.91) | 9.21E-03 | 2.95 (1.64-5.30) | 3.05E-04 | 2.51 (1.36-4.63) | 3.14E-03 |
|  | Heart Stress | 20/72 (27.78) | 8.04 (4.93-13.12) | 6.44E-17 | 4.33 (2.49-7.53) | 2.14E-07 | 4.38 (2.53-7.60) | 1.37E-07 |
| **CVD** |  |  |  |  |  |  |  |  |
| No Frailty | No Heart Stress | 394/2347 (16.79) |  |  |  |  |  |  |
|  | Heart Stress | 356/819 (43.47) | 3.28 (2.84-3.78) | 5.05E-59 | 2.42 (2.08-2.80) | 1.17E-31 | 2.42 (2.09-2.81) | 9.15E-32 |
| Frailty | No Heart Stress | 67/206 (32.52) | 2.28 (1.76-2.95) | 4.75E-10 | 2.85 (2.19-3.70) | 3.64E-15 | 2.81 (2.15-3.67) | 5.46E-14 |
|  | Heart Stress | 129/181 (71.27) | 7.97 (6.52-9.73) | 1.46E-91 | 5.39 (4.34-6.70) | 5.23E-52 | 5.48 (4.40-6.82) | 2.65E-52 |
| **Cancer** |  |  |  |  |  |  |  |  |
| No Frailty | No Heart Stress | 319/2272 (14.04) |  |  |  |  |  |  |
|  | Heart Stress | 152/615 (24.72) | 1.94 (1.60-2.36) | 1.69E-11 | 1.77 (1.46-2.15) | 9.32E-09 | 1.73 (1.42-2.10) | 4.89E-08 |
| Frailty | No Heart Stress | 53/192 (27.60) | 2.13 (1.59-2.84) | 3.72E-07 | 2.68 (2.00-3.58) | 4.14E-11 | 2.83 (2.09-3.84) | 1.79E-11 |
|  | Heart Stress | 34/86 (39.53) | 3.62 (2.54-5.16) | 9.95E-13 | 3.29 (2.27-4.76) | 3.18E-10 | 3.20 (2.21-4.64) | 7.83E-10 |
| **Others** | |  |  |  |  |  |  |  |
| No Frailty | No Heart Stress | 674/2627 (25.66) |  |  |  |  |  |  |
|  | Heart Stress | 394/857 (45.97) | 2.16 (1.91-2.45) | 7.28E-34 | 1.72 (1.51-1.95) | 5.91E-17 | 1.70 (1.50-1.93) | 2.63E-16 |
| Frailty | No Heart Stress | 124/263 (47.15) | 2.28 (1.88-2.77) | 3.09E-17 | 2.77 (2.28-3.36) | 5.04E-25 | 2.79 (2.29-3.39) | 3.04E-24 |
|  | Heart Stress | 169/221 (76.47) | 5.88 (4.96-6.97) | 1.18E-92 | 4.29 (3.58-5.13) | 1.04E-56 | 4.16 (3.46-4.99) | 9.28E-53 |

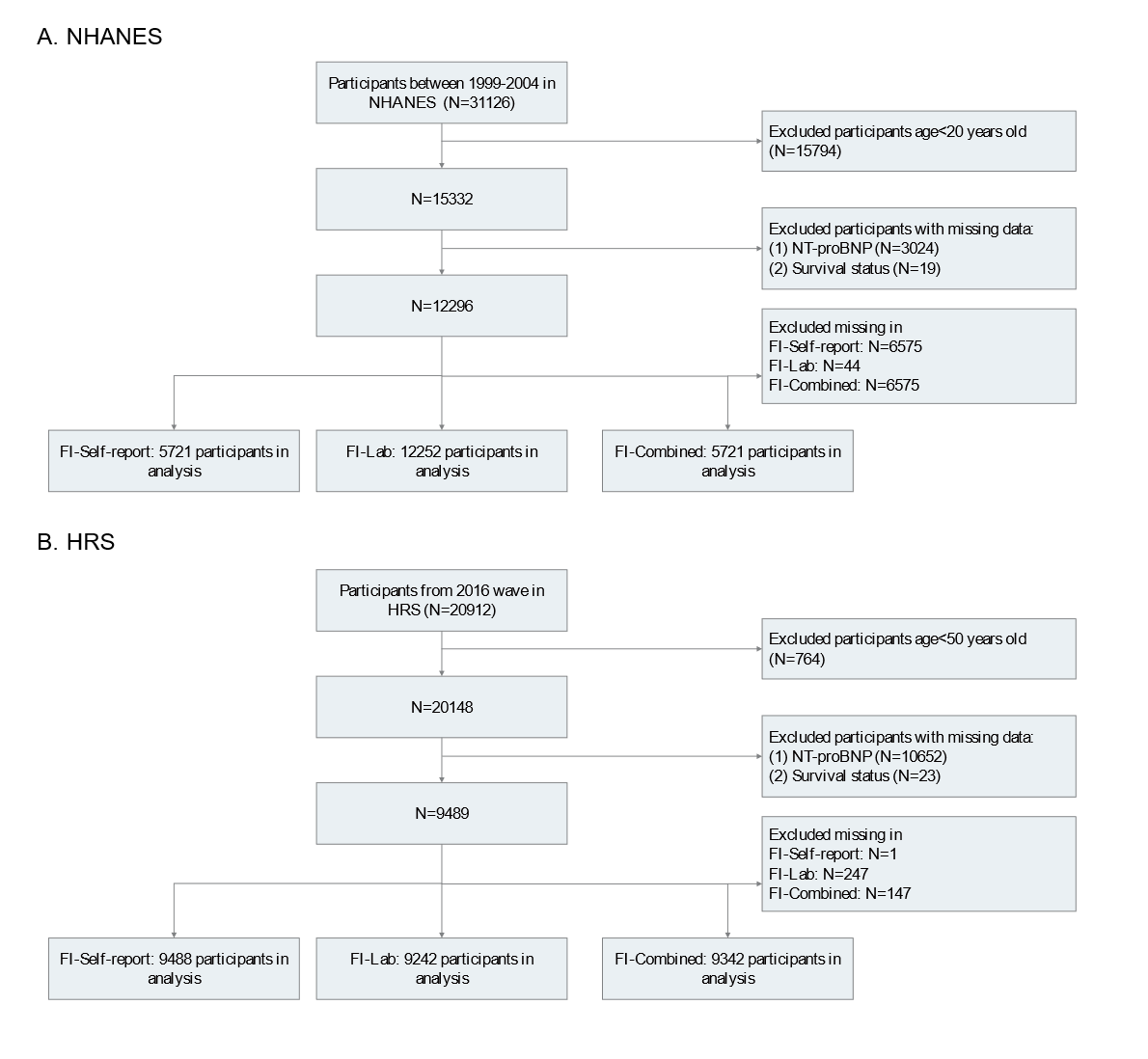
**eFigure 1.** Flowchart of the study.

NHANES indicates US National Health and Nutrition Examination Survey; HRS, Health and Retirement Study; NT-proBNP, N-terminal pro-B-type natriuretic peptide.

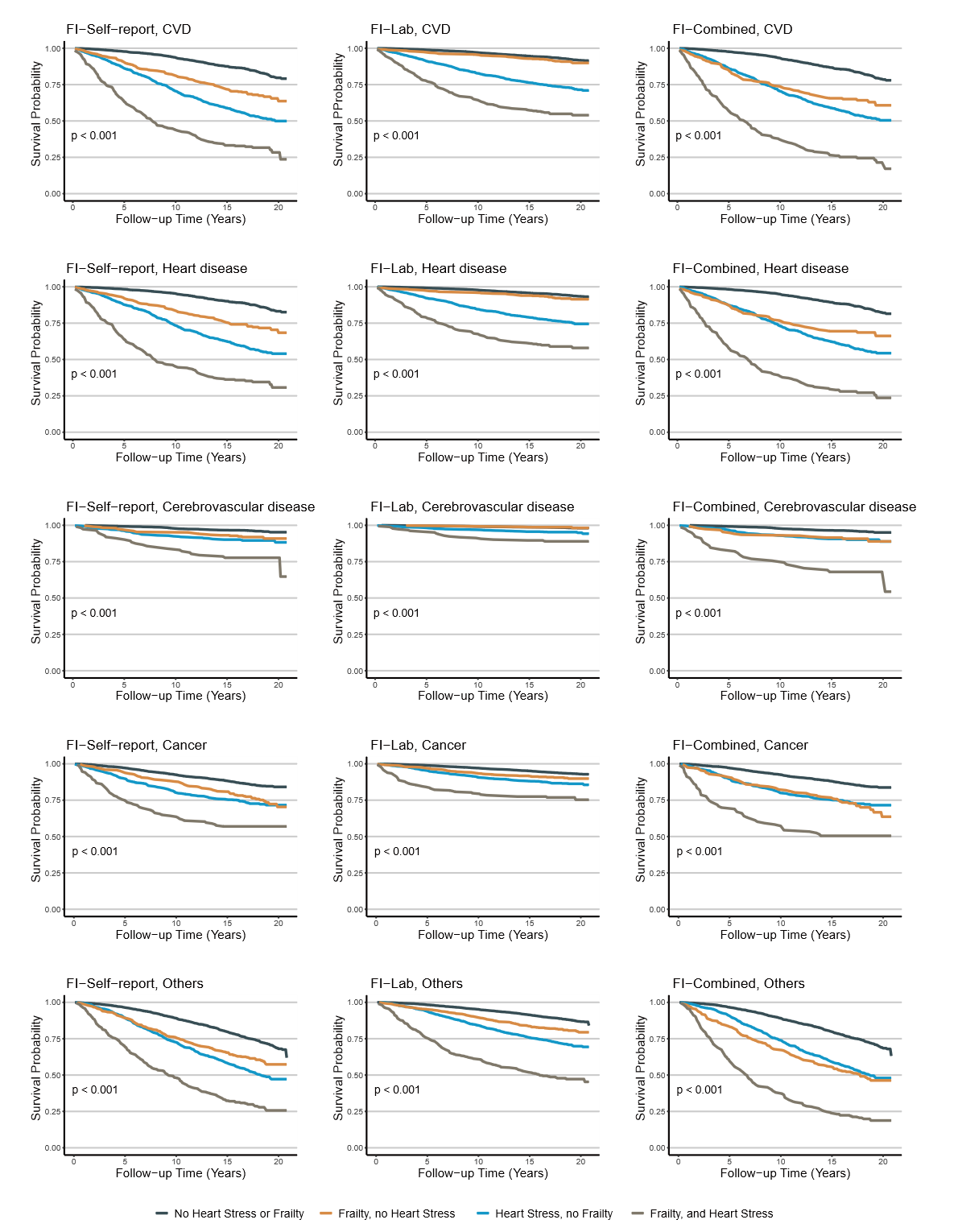
**eFigure 2.** Cause-specific mortality stratified by combined frailty and HS.

Kaplan-Meier plots demonstrate survival probabilities according to baseline frailty and HS status across 4 groups in the NHANES. Cause-specific mortality outcomes includes CVD, heart disease, cerebrovascular disease, cancer and other causes. HS: heart stress. NHANES: National Health and Nutrition Examination Survey. CVD: cardiovascular disease.

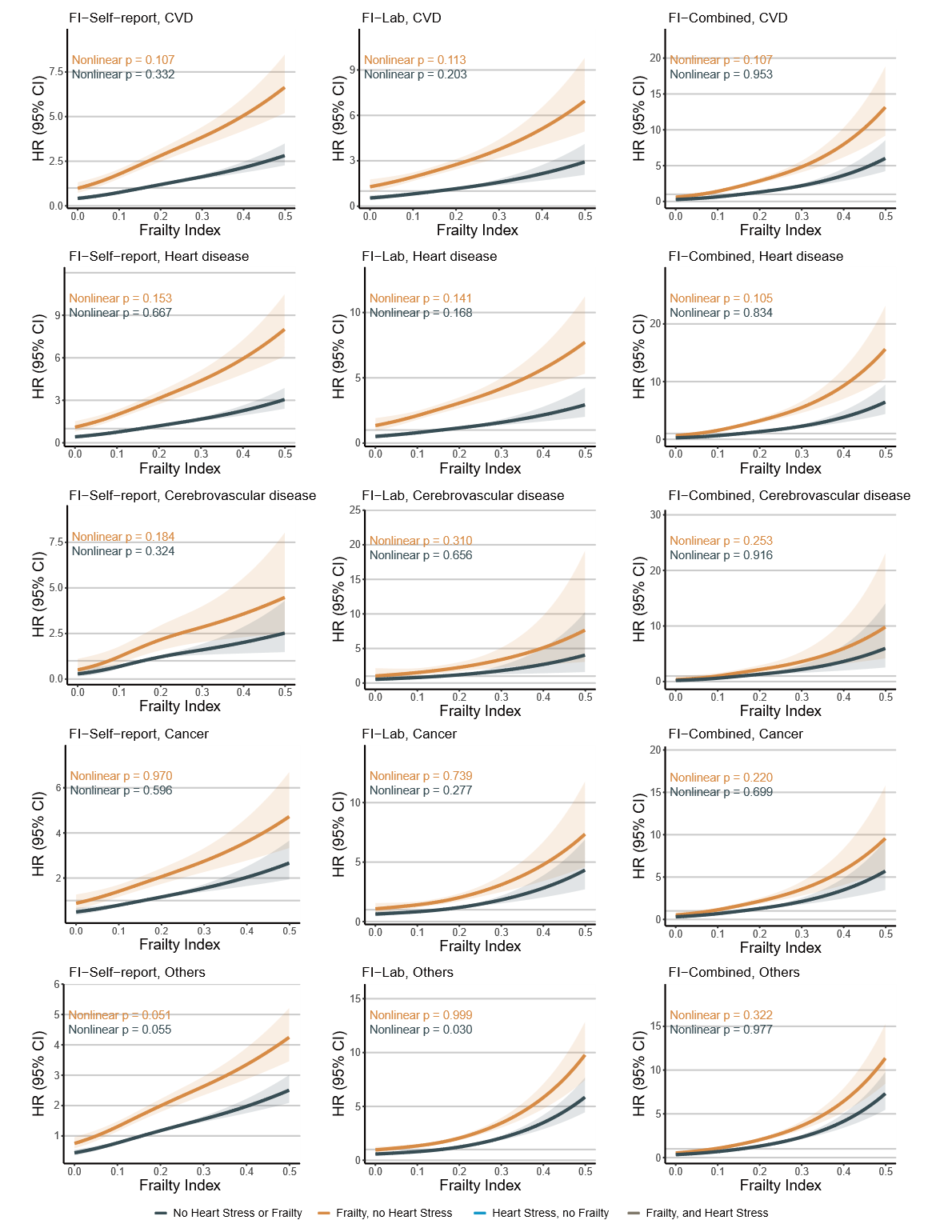
**eFigure 3.** Association between baseline FI and cause-specific mortality stratified by HS status.

Restricted cubic spline analyses show estimated hazard ratios for cause-specific mortality across continuous levels of FI in the NHANES. Cause-specific mortality outcomes includes CVD, heart disease, cerebrovascular disease, cancer and other causes. Models were adjusted for age, gender, race/ethnicity, education, body mass index, smoking status, physical activity, and marital status. FI: frailty index. HS: heart stress. NHANES: National Health and Nutrition Examination Survey. CVD: cardiovascular disease.

**eFigure 4.** All-cause mortality stratified by combined frailty and HS re-defined by a
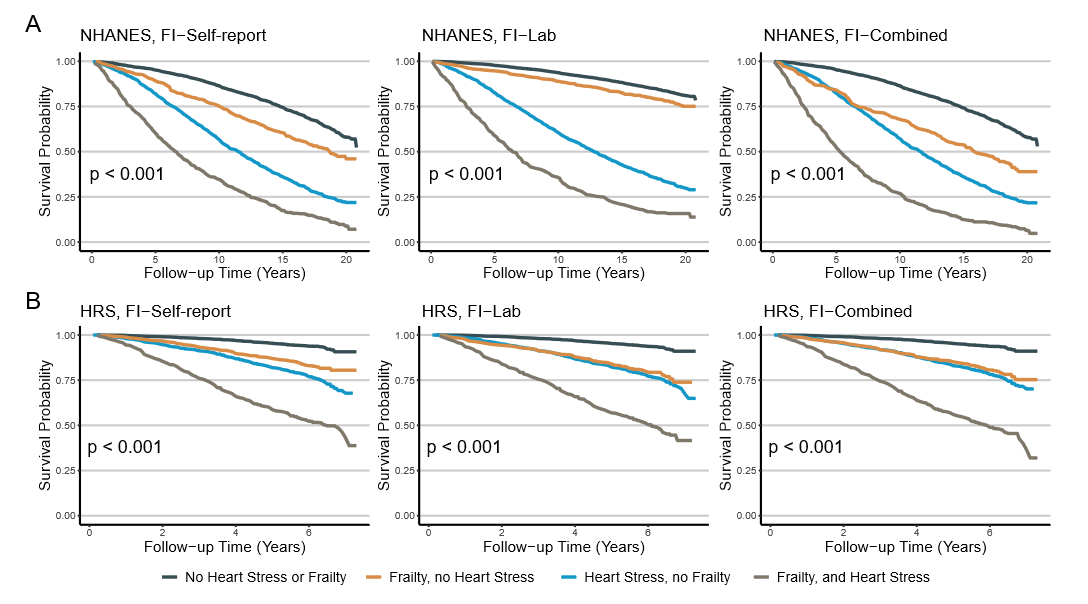
fixed cutoff in sensitivity analyses.

HS was re-defined by applying a fixed cutoff of NT-proBNP. Kaplan-Meier plots demonstrate survival probabilities according to baseline frailty and HS status across 4 groups in the NHANES (A) and the HRS (B) cohorts. HS: heart stress. NT-proBNP: N-terminal pro-B-type natriuretic peptide. NHANES: National Health and Nutrition Examination Survey. HRS, Health and Retirement Study.

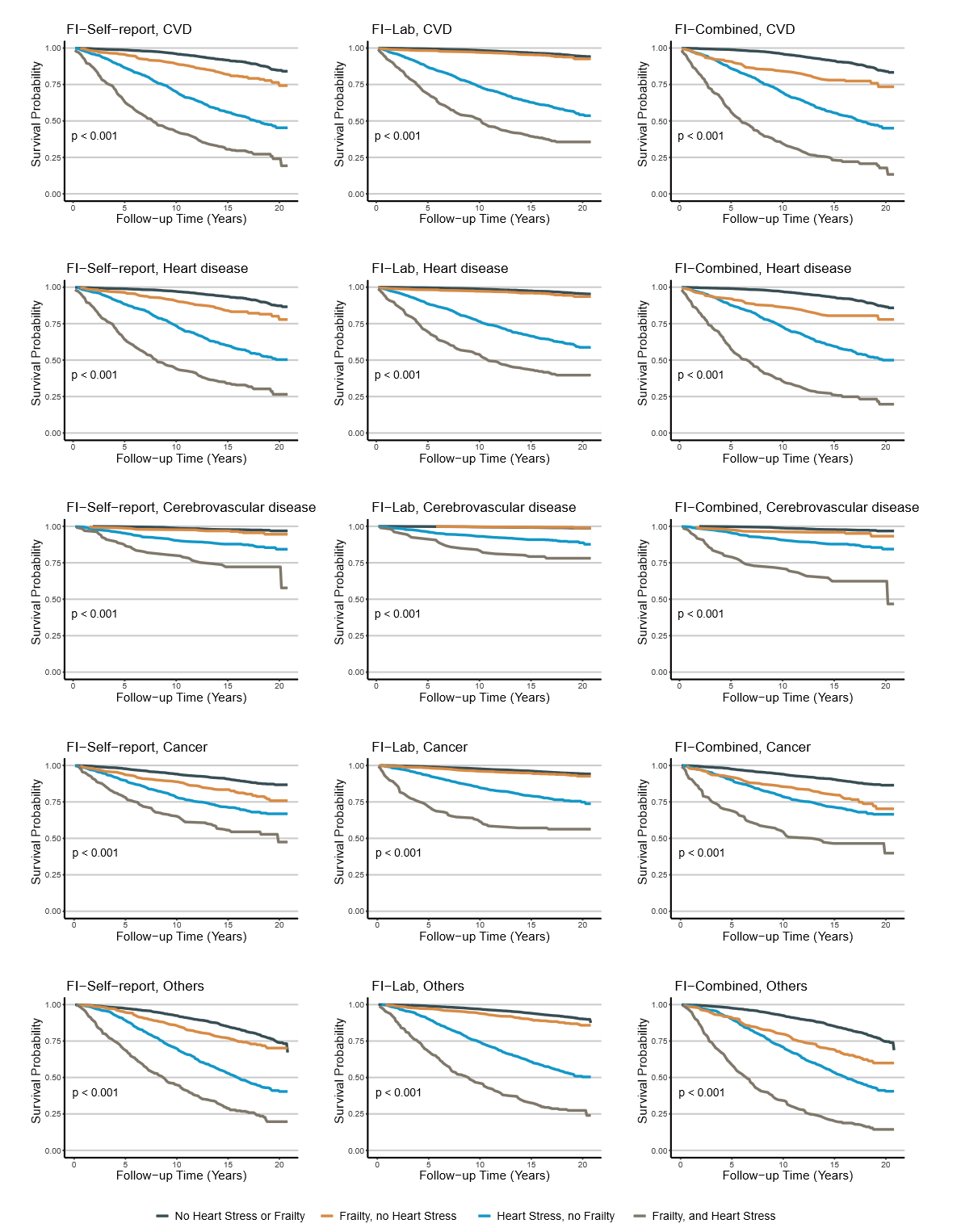
**eFigure 5.** Cause-specific mortality stratified by combined frailty and HS re-defined by a fixed cutoff in sensitivity analyses.

HS was re-defined by applying a fixed cutoff of NT-proBNP. Kaplan-Meier plots demonstrate survival probabilities according to baseline frailty and HS status across 4 groups in the NHANES. Cause-specific mortality outcomes includes CVD, heart disease, cerebrovascular disease, cancer and other causes. HS: heart stress. NT-proBNP: N-terminal pro-B-type natriuretic peptide. NHANES: National Health and Nutrition Examination Survey. CVD, cardiovascular disease.

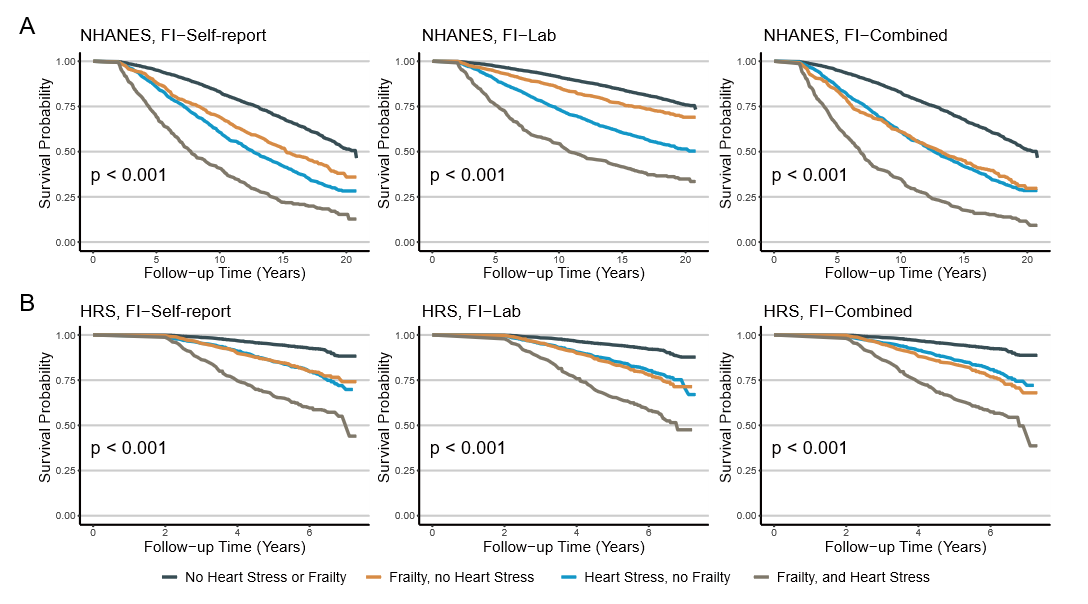
**eFigure 6.** All-cause mortality stratified by combined frailty and HS after excluding early deaths in sensitivity analyses.

Individuals who died within 2 years after baseline were excluded from analyses. Kaplan-Meier plots demonstrate survival probabilities according to baseline frailty and HS status across 4 groups in the NHANES (A) and HRS (B) cohorts. HS: heart stress. NHANES: National Health and Nutrition Examination Survey. HRS, Health and Retirement Study.

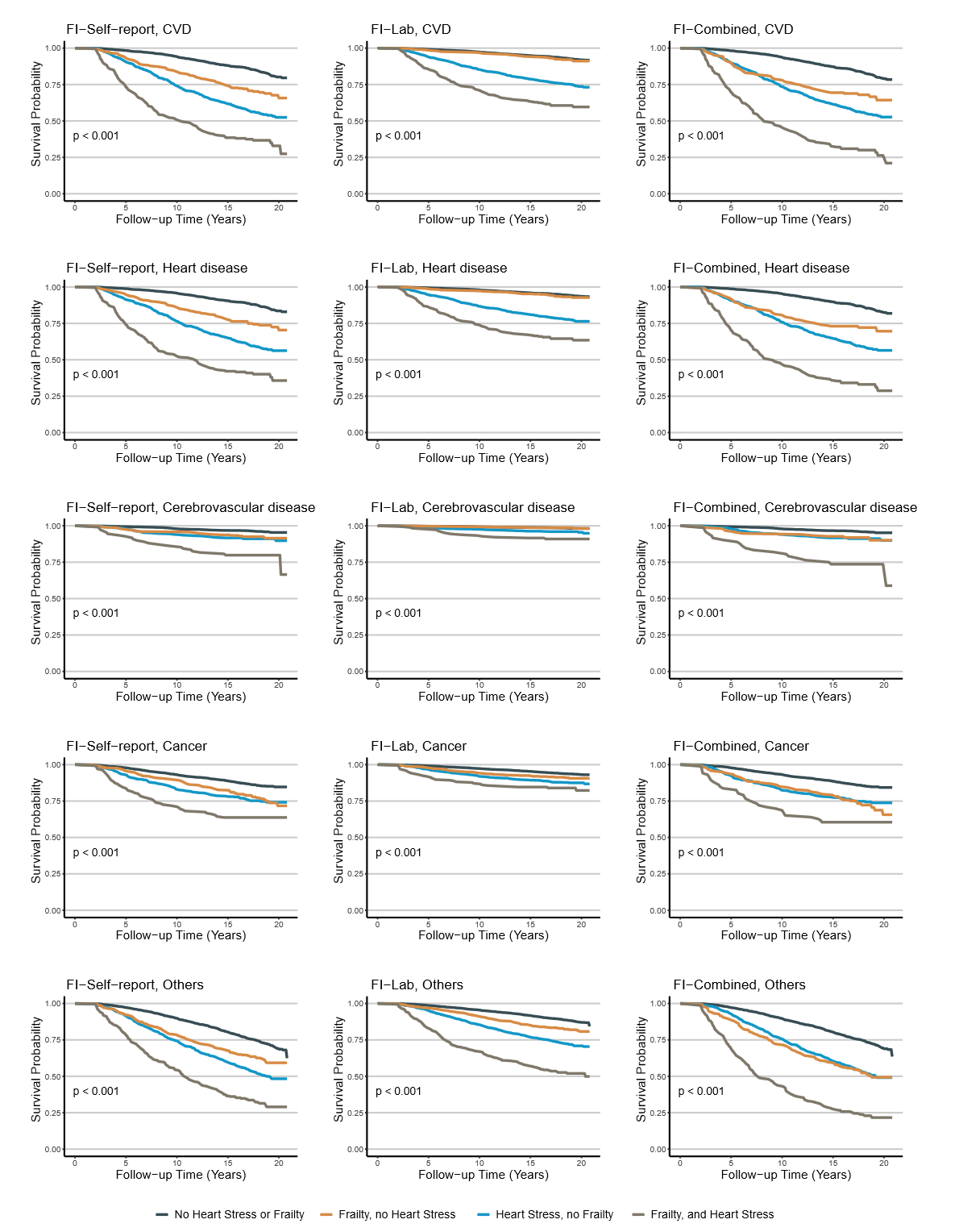
**eFigure 7.** Cause-specific mortality stratified by combined frailty and HS after excluding early deaths in sensitivity analyses.

Individuals who died within 2 years after baseline were excluded from analyses. Kaplan-Meier plots demonstrate survival probabilities according to baseline frailty and HS status across 4 groups in the NHANES. Cause-specific mortality outcomes includes CVD, heart disease, cerebrovascular disease, cancer and other causes. HS, heart stress. NHANES, US National Health and Nutrition Examination Survey. CVD, cardiovascular disease.

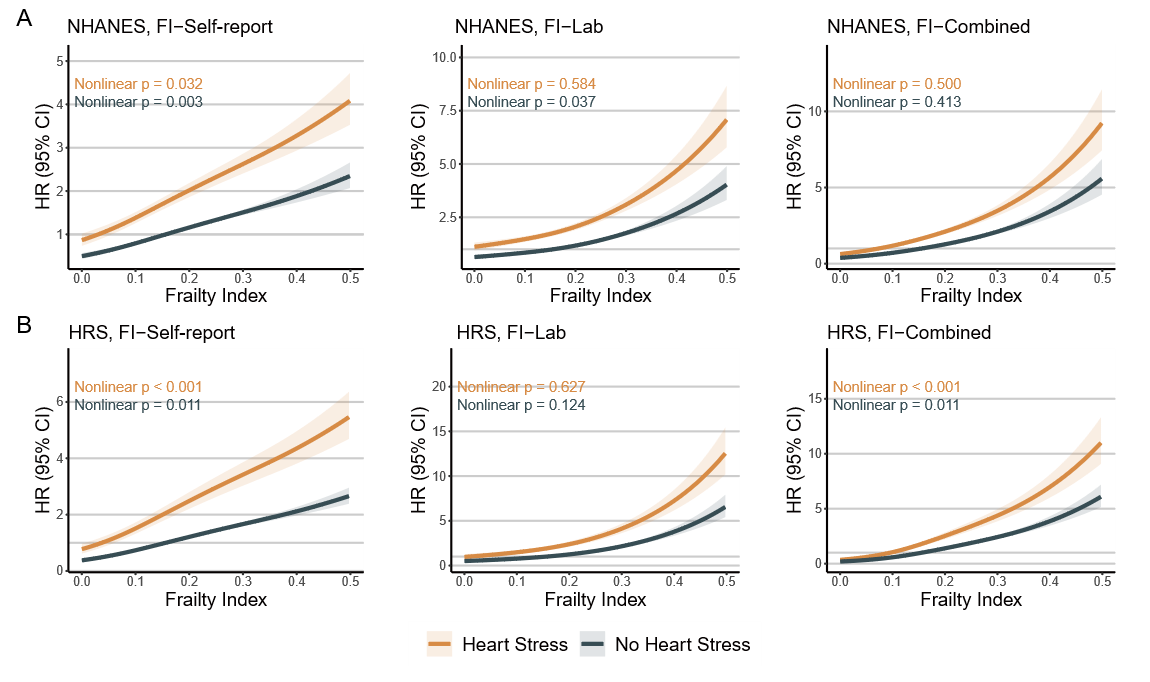
**eFigure 8.** Association between baseline FI and all-cause mortality stratified by HS status re-defined by a fixed cutoff in sensitivity analyses.

HS was re-defined by applying a fixed cutoff of NT-proBNP. Restricted cubic spline analyses show estimated hazard ratios for all-cause mortality across continuous levels of FI in the NHANES (A) and the HRS (B) cohorts. Models were adjusted for age, gender, race/ethnicity, education, BMI, smoking status, physical activity, and marital status. FI, frailty index**.** HS, heart stress. NT-proBNP, N-terminal pro-B-type natriuretic peptide. NHANES, US National Health and Nutrition Examination Survey. HRS, Health and Retirement Study.

**eFigure 9.** Association between baseline FI and cause-specific mortality stratified by HS status re-defined by a fixed cutoff in sensitivity analyses.

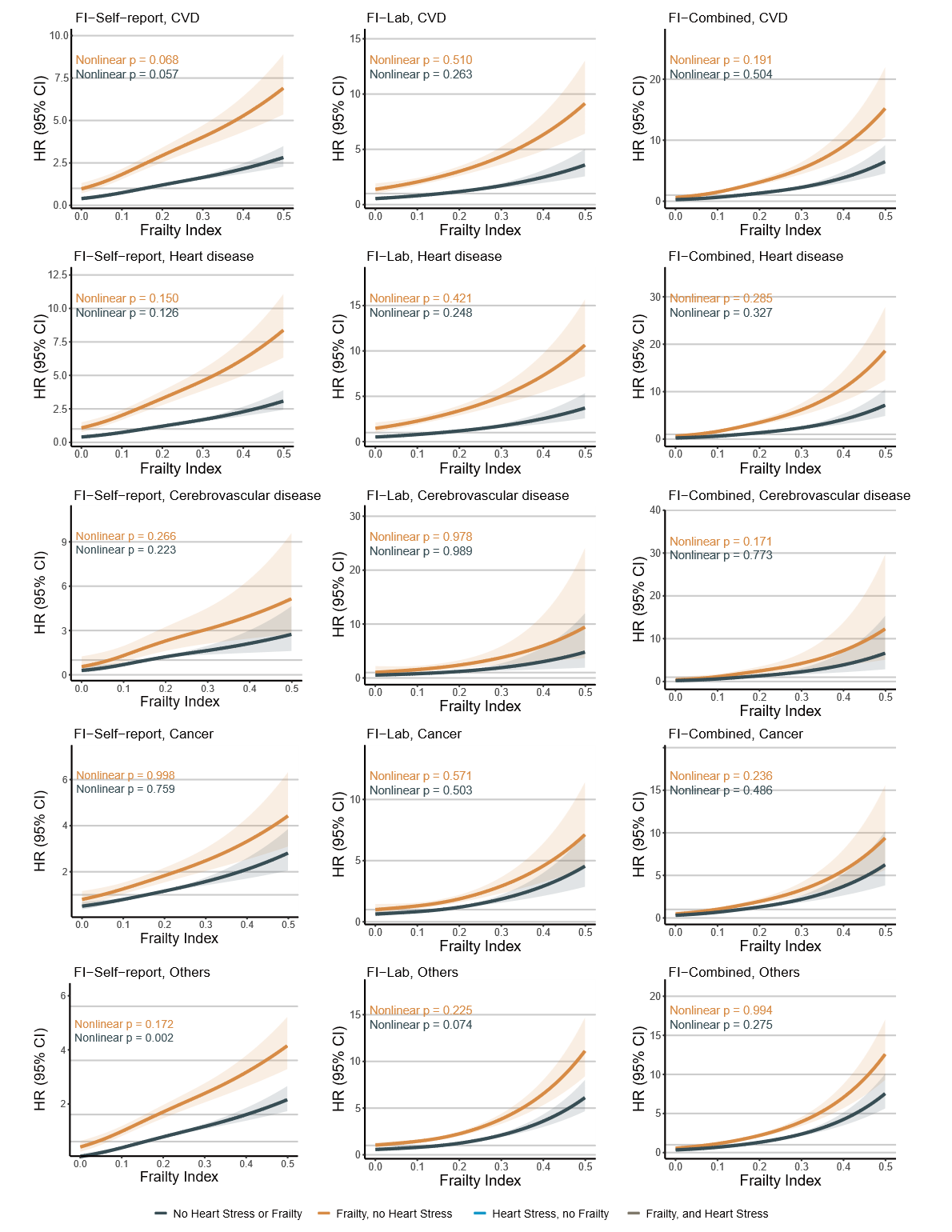
HS was re-defined by applying a fixed cutoff of NT-proBNP. Restricted cubic spline analyses show estimated hazard ratios for cause-specific mortality across continuous levels of FI in the NHANES. Cause-specific mortality outcomes includes CVD, heart disease, cerebrovascular disease, cancer and other causes. Models were adjusted for age, gender, race/ethnicity, education, body mass index, smoking status, physical activity, and marital status. FI, frailty index**.** HS, heart stress. NT-proBNP, N-terminal pro-B-type natriuretic peptide. NHANES, US National Health and Nutrition Examination Survey. CVD, cardiovascular disease.

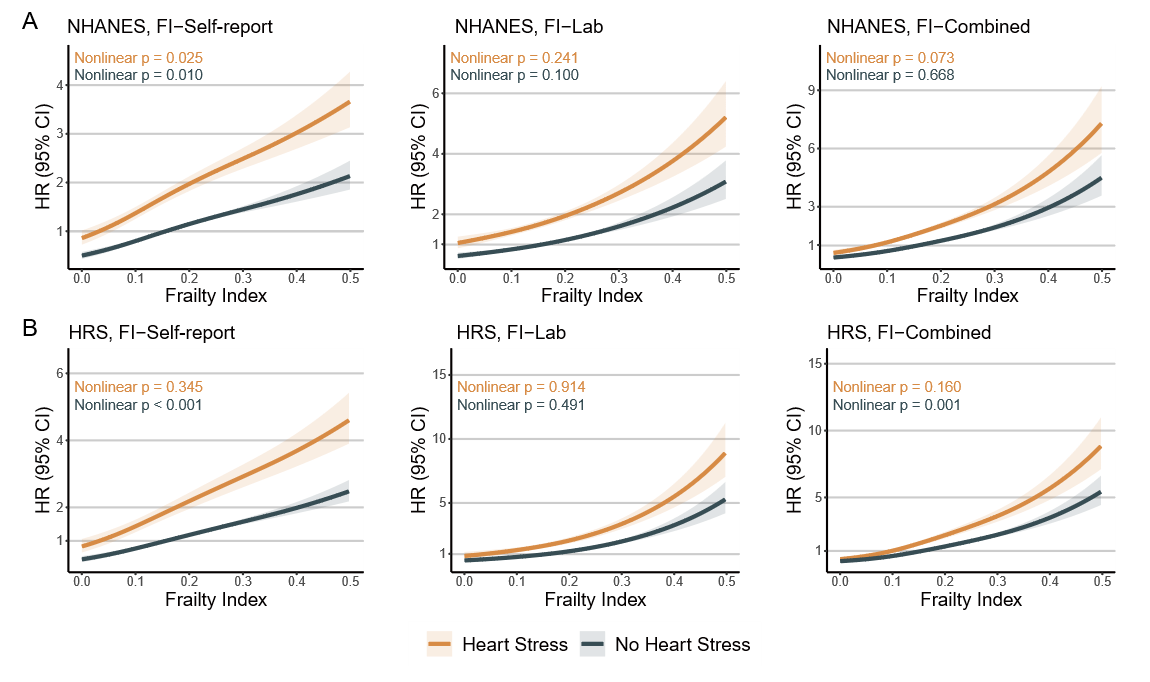
**eFigure 10.** Association between baseline FI and all-cause mortality stratified by HS status after excluding early deaths in sensitivity analyses.

Individuals who died within 2 years after baseline were excluded from analyses. Restricted cubic spline analyses show estimated hazard ratios for all-cause mortality across continuous levels of FI in the NHANES (A) and the HRS (B) cohorts. Models were adjusted for age, gender, race/ethnicity, education, body mass index, smoking status, physical activity, and marital status. FI, frailty index**.** HS, heart stress. NHANES, US National Health and Nutrition Examination Survey. HRS, Health and Retirement Study.

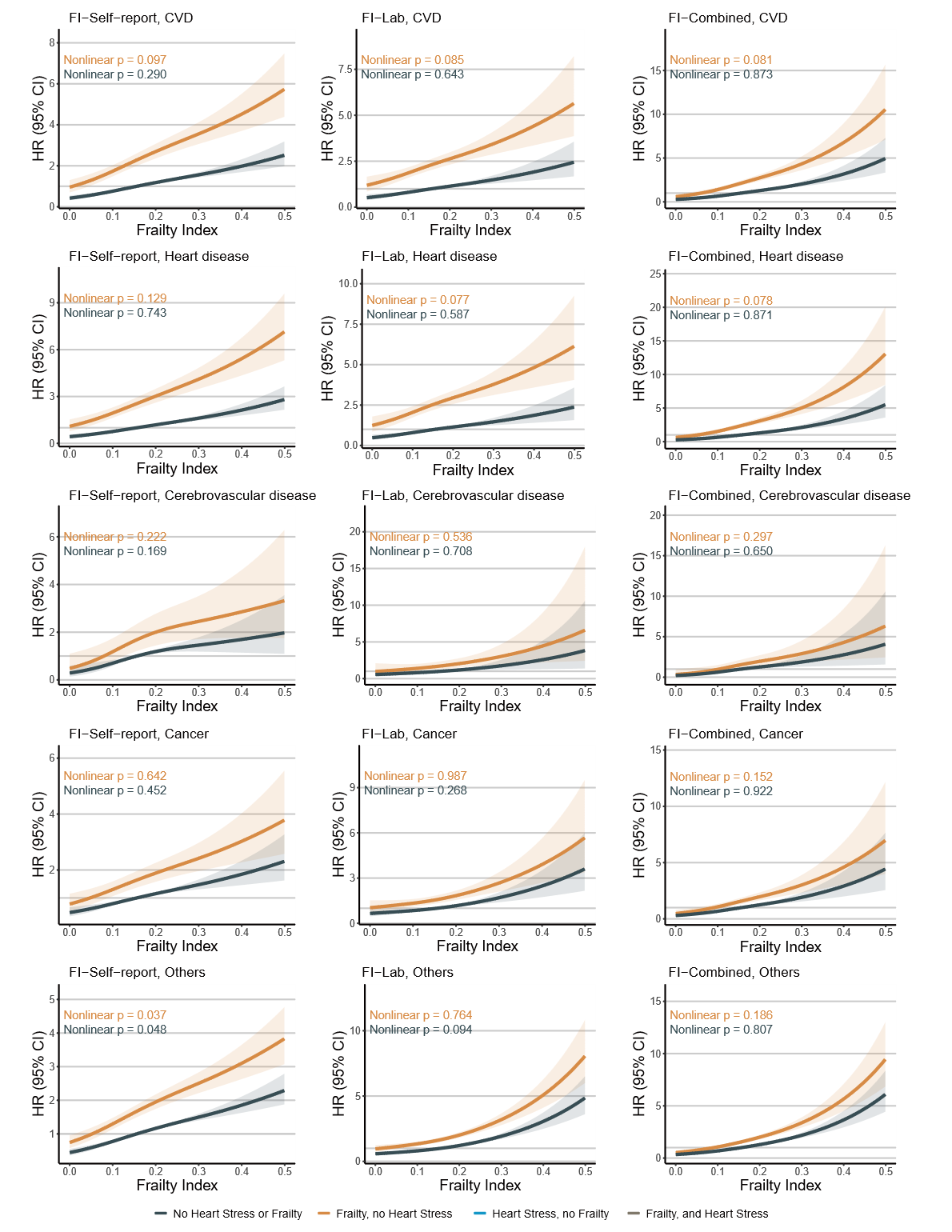
**eFigure 11.** Association between baseline FI and cause-specific mortality stratified by HS status after excluding early deaths in sensitivity analyses.

Individuals who died within 2 years after baseline were excluded from analyses. Restricted cubic spline analyses show estimated hazard ratios for cause-specific mortality across continuous levels of FI in the NHANES. Cause-specific mortality outcomes includes CVD, heart disease, cerebrovascular disease, cancer and other causes. Models were adjusted for age, gender, race/ethnicity, education, body mass index, smoking status, physical activity, and marital status. FI, frailty index**.** HS, heart stress. NHANES, US National Health and Nutrition Examination Survey. CVD, cardiovascular disease.
